# Cost-effectiveness of broadly neutralizing antibodies for infant HIV prophylaxis in settings with high HIV burdens: a simulation modeling study

**DOI:** 10.1101/2023.11.06.23298184

**Authors:** Christopher Alba, Shelly Malhotra, Stephanie Horsfall, Matthew E. Barnhart, Adrie Bekker, Katerina Chapman, Coleen K. Cunningham, Patricia E. Fast, Genevieve G. Fouda, Kenneth A. Freedberg, Ameena Goga, Lusine R. Ghazaryan, Valériane Leroy, Carlyn Mann, Margaret M. McCluskey, Elizabeth J. McFarland, Vincent Muturi-Kioi, Sallie R. Permar, Roger Shapiro, Devin Sok, Lynda Stranix-Chibanda, Milton C. Weinstein, Andrea L. Ciaranello, Caitlin M. Dugdale

## Abstract

**Introduction:** Approximately 130 000 infants acquire HIV annually despite global maternal antiretroviral therapy scale-up. We evaluated the potential clinical impact and cost-effectiveness of offering long-acting, anti-HIV broadly neutralizing antibody (bNAb) prophylaxis to infants in three distinct settings.

**Methods:** We simulated infants in Côte d’Ivoire, South Africa, and Zimbabwe using the Cost-Effectiveness of Preventing AIDS Complications-Pediatric (CEPAC-P) model. We modeled strategies offering a three-bNAb combination in addition to WHO-recommended standard-of-care oral prophylaxis to infants: a) with known, WHO-defined high-risk HIV exposure at birth (*HR-HIVE*); b) with known HIV exposure at birth (*HIVE*); or c) with or without known HIV exposure (*ALL*). Modeled infants received *1-dose*, *2-doses*, or *Extended* (every 3 months through 18 months) bNAb dosing. Base case model inputs included 70% bNAb efficacy (sensitivity analysis range: 10-100%), 3-month efficacy duration/dosing interval (1-6 months), and $20/dose cost ($5-$100/dose). Outcomes included pediatric HIV infections, life expectancy, lifetime HIV-related costs, and incremental cost-effectiveness ratios (ICERs, in US$/year-of-life-saved [YLS], assuming a ≤50% GDP per capita cost-effectiveness threshold).

**Results:** The base case model projects that bNAb strategies targeting *HIVE* and *ALL* infants would prevent 7-26% and 10-42% additional pediatric HIV infections, respectively, compared to standard-of-care alone, ranging by dosing approach. *HIVE-Extended* would be cost-effective (cost-saving compared to standard-of-care) in Côte d’Ivoire and Zimbabwe; *ALL-Extended* would be cost-effective in South Africa (ICER: $882/YLS). BNAb strategies targeting *HR-HIVE* infants would result in greater lifetime costs and smaller life expectancy gains than *HIVE-Extended*. Throughout most bNAb efficacies and costs evaluated in sensitivity analyses, targeting *HIVE* infants would be cost-effective in Côte d’Ivoire and Zimbabwe, and targeting *ALL* infants would be cost-effective in South Africa.

**Discussion:** Adding long-acting bNAbs to current standard-of-care prophylaxis would be cost-effective, assuming plausible efficacies and costs. The cost-effective target population would vary by setting, largely driven by maternal antenatal HIV prevalence and postpartum incidence.

## INTRODUCTION

In 2022, an estimated 130 000 children acquired HIV worldwide, of whom approximately half acquired HIV while breastfeeding.^1^ An estimated two-thirds of children who acquired HIV had mothers living with undiagnosed HIV during pregnancy or who acquired HIV during pregnancy or breastfeeding.^2^ Therefore, in addition to improving maternal HIV detection and rapid antiretroviral therapy (ART) initiation, novel biomedical strategies offering protection against vertical HIV transmission from unrecognized and/or acute maternal HIV infection may be needed to eliminate vertical HIV transmission.

Given that vertical HIV transmission risk is time-bound by the end of breastfeeding, infant prophylaxis may be an ideal application of the passive immunity offered by anti-HIV broadly neutralizing antibodies (bNAbs). The bNAb VRC01 has demonstrated efficacy in preventing HIV acquisition of VRC01-sensitive virus in adults, and VRC01, VRC01-LS, and VRC07-523-LS have demonstrated safety when given to both adults and newborn infants.^3–7^ Pharmacokinetic studies suggest that some bNAbs, such as VRC07-523LS, can maintain potentially protective levels for up to three months after subcutaneous administration in infants.^7^ While there are no published studies of bNAb efficacy in human infants, bNAbs have demonstrated efficacy against both perinatal and postnatal simian-HIV acquisition in non-human primate models.^8,9^ At least ten bNAbs are under clinical investigation for HIV-1 treatment or prophylaxis, including combination bNAb products, such as PGDM1400+PGT121+VRC07-523LS, which may have lower production costs and higher efficacy due to increased breadth and potency (NCT03205917, NCT03928821, NCT04212091, NCT03721510).^10^ If proven clinically effective, universally administering bNAbs for infant prophylaxis in settings with high HIV burdens could provide protection against vertical HIV transmission during the breastfeeding period, including for infants with mothers with unrecognized and/or acute HIV infection during pregnancy and breastfeeding for whom there are limited prevention strategies. Among infants with recognized HIV exposure, bNAb prophylaxis potentially offers a way to overcome adherence, tolerability, and resistance challenges with current standard-of-care oral infant antiretroviral prophylaxis. Linking bNAb infant prophylaxis into routine immunization or maternal and child health care infrastructure could also potentially facilitate broad delivery, streamline supply chain and storage, and mitigate against issues of stigma by delinking from HIV programmatic services.

Our previous modeling work found that offering a bNAb to infants with known HIV exposure at birth would be cost-effective in Côte d’Ivoire, South Africa, and Zimbabwe.^11^ However, the potential long-term, population-level clinical benefits and cost-effectiveness of a universal infant bNAb prophylaxis program in high HIV burden settings have not yet been studied. Here, we extend our prior modeling work to include infants born to mothers with unrecognized HIV infection or mothers at risk for acute HIV acquisition postpartum. We aimed to estimate the clinical impact, costs, and cost-effectiveness of offering combination bNAb prophylaxis—in addition to existing standard-of-care HIV prevention services—to infants in Côte d’Ivoire, South Africa, and Zimbabwe.

## METHODS

### Study design

We projected the clinical and economic impacts of bNAb strategies using the validated Cost-Effectiveness of Preventing AIDS Complications–Pediatrics (CEPAC-P) microsimulation model.^12^ We modeled infant cohorts from birth to death in Côte d’Ivoire, South Africa, and Zimbabwe—countries with high HIV burdens chosen to represent settings with varied maternal HIV prevalence, postpartum HIV incidence, knowledge of HIV status, ART uptake, breastfeeding practices, and economic constraints (Table 1). This study adheres to the Consolidated Health Economic Evaluation Reporting Standards (Supplementary Table 1) and was approved by the Mass General Brigham Human Research Committee.

**Table 1.**
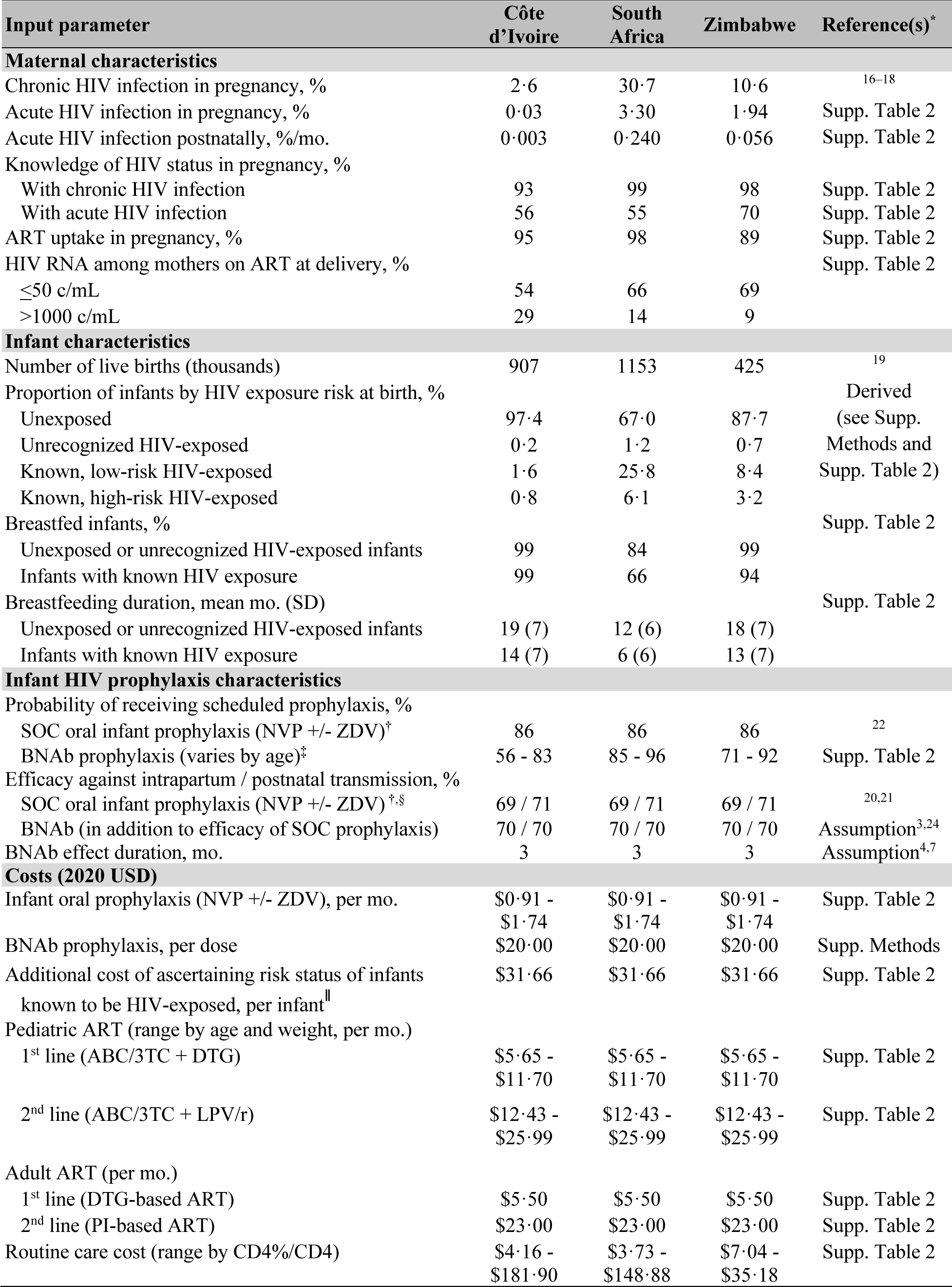
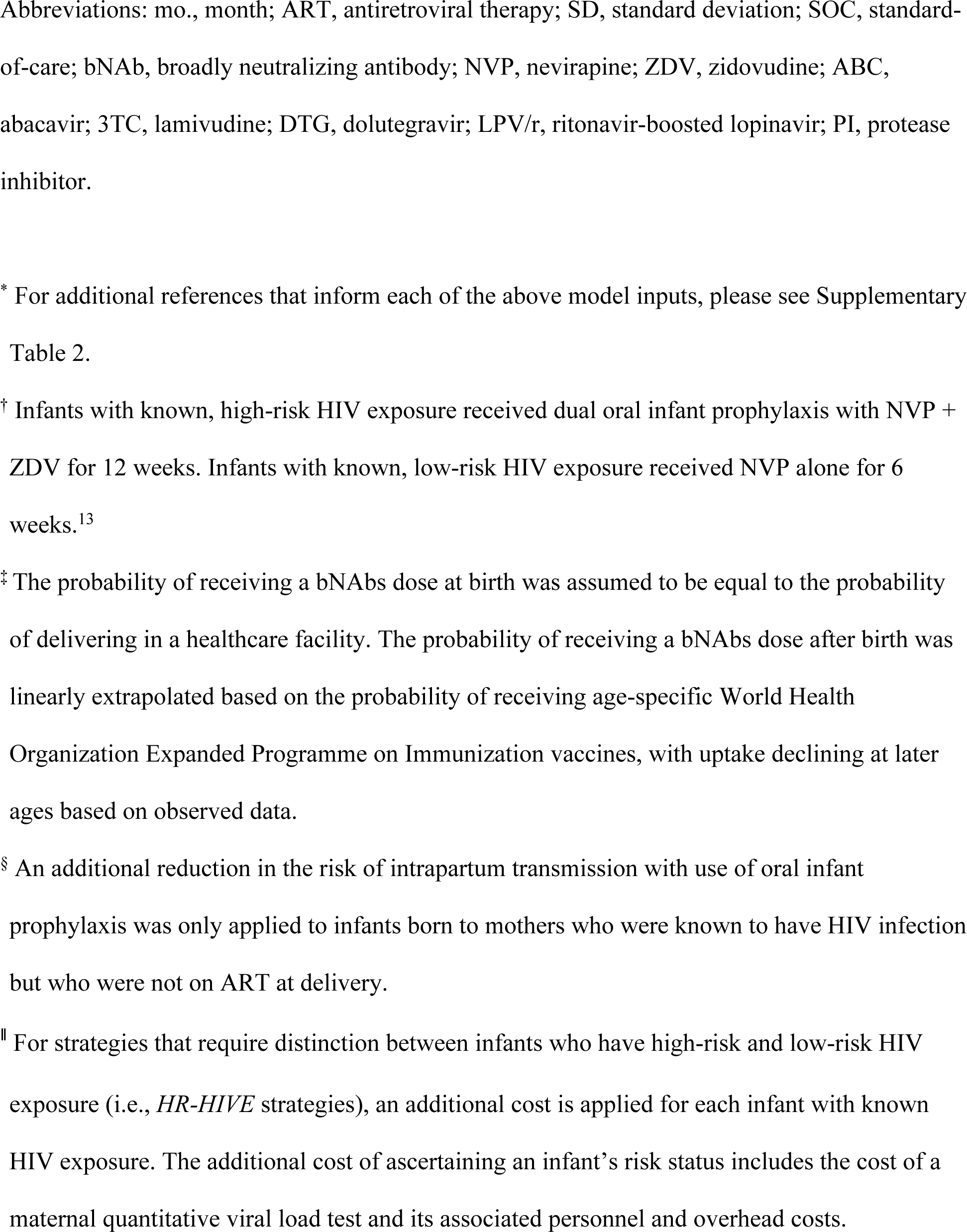
Selected base case model input parameters.

### Model overview

Infants enter the CEPAC-P model at birth and experience a probability of acquiring HIV perinatally (i.e., intrauterine/intrapartum transmission) and a monthly probability of acquiring HIV postnatally, while breastfeeding, depending on their mother’s HIV status, ART engagement, and virologic suppression. Modeled infants known to be HIV-exposed at birth are eligible to receive 6-12 weeks of World Health Organization (WHO)-recommended oral antiretroviral postnatal prophylaxis,^13^ reducing their HIV acquisition risk. Infants may also be eligible to receive bNAbs at birth and during breastfeeding (see “Modeled strategies” below) and, if received, the infants can experience an additional reduction in intrapartum and/or postnatal HIV acquisition. Children who acquire HIV face monthly probabilities of incident opportunistic infections (OIs) and AIDS-related death. Children with HIV can be detected by presenting with an OI or via routine early infant diagnosis (EID). Once detected, children experience a probability of ART initiation. While on effective ART, children experience reduced monthly OI and AIDS-related mortality probabilities (Supplementary Table 2).

### Modeled populations

For each country, we modeled four mutually exclusive infant sub-cohorts based on WHO-defined HIV exposure risk. These sub-cohorts comprised infants who, at birth, were: a) known to be HIV-exposed and high-risk (i.e., infant’s mother was known to have acquired HIV during pregnancy, received less than four weeks of ART before delivery, or had a viral load ≥1000 copies/mL); b) known to be HIV-exposed and low-risk (i.e., infant’s mother was on ART with a viral load <1000 copies/mL at delivery); c) unrecognized as HIV-exposed; or d) HIV-unexposed (although potentially at risk for HIV exposure postnatally through breastfeeding) (Supplementary Figure 2).^13^ Together, these four sub-cohorts represent all infants born in a country annually. We calculated the proportion of infants within each sub-cohort using country-specific estimates of maternal HIV prevalence/incidence, knowledge of HIV status, ART uptake, and virologic suppression (Table 1; Supplementary Methods and Supplementary Table 2).

### Modeled strategies

First, we modeled a current standard-of-care strategy in which infants with known HIV exposure (low- or high-risk) were eligible to receive WHO-recommended antiretroviral prophylaxis without bNAbs. Then, we modeled several bNAb strategies assessing different target populations and bNAb dosing approaches (Supplementary Figure 2). The three target populations included: a) infants with known, high-risk HIV exposure at birth (*HR-HIVE* strategies); b) infants with known HIV exposure at birth irrespective of high- or low-risk (*HIVE* strategies); or c) all infants with or without known HIV exposure (*ALL* strategies). For each target population, we modeled three bNAb dosing approaches in which eligible infants could receive: a) one dose of bNAbs at birth (*1-dose*); b) two doses of bNAbs, once at birth and once at three months (*2-doses*); or c) extended dosing, starting at birth and every three months thereafter for up to 18 months while breastfeeding (to align with existing programs that administer childhood vaccines through 18 months of age and mean breastfeeding duration across these settings) (*Extended*).^4,14,15^ Additionally, we modeled hybrid strategies in which different target populations were offered different bNAb dosing approaches, for example, a strategy in which infants with known HIV exposure were offered extended doses of bNAbs (i.e., every 3 months) while infants without known HIV exposure were offered only one dose of bNAbs at birth. For simplicity, in the primary results, we present only the one hybrid strategy that would not be both less beneficial and more costly than at least one of the non-hybrid strategies (i.e., *ALL–1-dose* plus *HIVE– Extended*); projected clinical outcomes and costs of all other hybrid strategies can be found in Supplementary Table 5. In all bNAb strategies in the base case, infants with known HIV exposure continued to receive standard-of-care antiretroviral prophylaxis. However, we explored the potential impact of bNAb prophylaxis alone without antiretroviral prophylaxis in a sensitivity analysis.

### Model inputs

#### Cohort characteristics

We informed cohort characteristics using UNAIDS estimates, country-specific survey data, and published literature (Table 1; Supplementary Table 2). We modeled recent country-specific antenatal HIV prevalence (Côte d’Ivoire: 2·6%, South Africa: 30·7%, and Zimbabwe: 10·6%)^16–18^ and postpartum maternal HIV incidence (Côte d’Ivoire: 0·003%/month, South Africa: 0·240%/month, and Zimbabwe: 0·056%/month; Supplementary Table 2). Knowledge of maternal status, probability of acute infection in pregnancy, maternal ART uptake, proportion of women with virologic suppression, and breastfeeding duration also varied by country (Table 1).

Using the cohort characteristics, we estimated the proportion of infants in each of the four risk exposure groups at the time of delivery (i.e., high-risk HIV-exposed, low-risk HIV-exposed unrecognized HIV-exposed, and HIV-unexposed) (Supplementary Methods) and scaled cohort sizes to the number of annual live births estimated by UNICEF.^19^ Model inputs regarding HIV natural history, background mortality, ART efficacy, EID cascade, and HIV-related costs were also informed by published literature (Table 1; Supplementary Table 2).

#### Infant HIV prophylaxis

Based on clinical trial data, standard-of-care oral antiretroviral infant prophylaxis was modeled to have 69% efficacy against intrapartum transmission and 71% efficacy against postnatal transmission and was ranged widely in sensitivity analyses.^20,21^ Adherence to oral antiretroviral prophylaxis was modeled to be 86%.^22^

IMPAACT P1112 is the only completed Phase I study investigating single bNAbs as infant HIV prophylaxis and has demonstrated the safety, tolerability, and favorable pharmacokinetics of VRC01, VRC01LS, and VRC07-523LS.^4,15^ Additional Phase I/II studies in HIV-exposed and unexposed newborns are in progress or development, including PedMAB, SAMBULELO, and EDCTP Neo bnAb.^23^ While bNAb prophylaxis remains investigational, Phase IIb trials found that acquisition of VRC01-sensitive isolates was 75% lower in adults who received VRC01, compared to those who received placebo.^3^ VRC07-523LS, a more potent bNAb, has demonstrated greater neutralizing breadth than VRC01 (91% versus 66% at IC_80_ <10 µg/mL) against primary African isolates in multiclade panels and is well-tolerated in infants.^7,24^ Similar to combination ART, studies have found that bNAb combinations targeting multiple different HIV-1 epitopes have greater potency and neutralizing breadth than individual bNAb products alone.^24,25^ Long-acting bNAb combinations currently under investigation, such as combinations of PGT121, PGDM1400, and VRC07-523LS, will likely have even greater breadth and/or potency.^24^ As such, in the base case, we assumed a three-bNAb combination would have 70% efficacy against intrapartum and postnatal HIV transmission, in addition to the vertical transmission reduction conferred by oral infant prophylaxis for eligible infants, with no impact on in utero transmission (Supplementary Methods). This 70% efficacy is slightly higher than would be estimated by multiplying the efficacy of VRC01 against sensitive isolates and breadth of VRC07-523LS alone, with a base case three-month effect duration per administration based on pharmacokinetic data of subcutaneous VRC07-523LS administration in infants.^4,7^ The uptake of a birth bNAb dose among eligible infants was bounded by the percent of births performed in a healthcare facility (Supplementary Table 2). The uptake of subsequent bNAb doses in the postpartum period were informed by setting-specific uptake of other routine age-specific immunizations at comparable timepoints, assuming a strategy of integration into Extended Programme on Immunization programs (Supplementary Table 2).

#### Costs

Antiretroviral prophylaxis and ART costs were informed by Global Fund price lists and WHO-recommended weight-based formulations (Supplementary Table 2). The base case average cost for a 3-bNAb combination was modeled as $20 per dose, including the estimated production costs ($11, assuming a 100mg bNAb dose), as well as routine delivery (including training, personal protective equipment, cold-chain, and social mobilization, $2), personnel ($1), and facility/overhead ($3) costs based on an analysis of vaccine delivery costing data from 92 countries, conservatively rounded up to the nearest $10 (Supplementary Methods).^26–29^ We also applied a cost of identifying high-risk infants in the *HR-HIVE* strategies by adding the cost of a maternal quantitative viral load test, associated clinic visit (including personnel and overhead costs), and result return with counseling ($31·66) per infant known to be HIV-exposed but in a secondary analysis also examined the cost-effectiveness of *HR-HIVE* strategies when withholding this cost (Supplementary Table 2). All costs are reported in 2020 USD.

### Modeled outcomes

We projected cumulative vertical HIV transmission by 36 months of age, undiscounted and discounted (3% per year) life expectancy, and average lifetime HIV-related costs from the healthcare system perspective.^30^ A lifetime time horizon was chosen to capture all clinical and economic impacts of averting HIV infections. We calculated incremental cost-effectiveness ratios (ICERs; USD/year of life saved [YLS]) by ordering strategies by increasing life expectancy and dividing the difference in discounted costs by the difference in discounted life expectancy of consecutive, non-dominated strategies. A strategy was considered dominated if it resulted in lower life expectancy gains than a lower cost strategy (i.e., strong dominance) or if it resulted in lower life expectancy gains per dollar spent than a higher cost strategy (i.e., extended dominance). For each country, the strategy that resulted in the greatest clinical benefit with an ICER ≤50% of a country’s 2020 GDP per capita (Côte d’Ivoire: $1163/YLS, South Africa: $2828/YLS, Zimbabwe: $607/YLS) was considered cost-effective based on current literature-based cost-effectiveness thresholds in resource-limited settings (Supplementary Methods). We also assessed the impact of using a more conservative cost-effectiveness threshold (≤20% of a country’s 2020 GDP per capita); we present these results only in the Supplement since this approach did not change the cost-effective strategy under base case assumptions (Supplementary Figure 8). Non-dominated strategies that resulted in higher life expectancy and lower costs than the standard-of-care were also marked as being cost-saving.

### Sensitivity analyses

We conducted univariate sensitivity analyses to test the robustness of our findings to changes in key input parameters. Given uncertainty in product characteristics, we independently varied bNAb protective efficacy (10-100%), effect duration (1-6 months), and costs ($5-$100 per dose, or 0.25x-5x base case cost, to capture scenarios in which bNAbs cost less due to more efficient production at scale or cost more due to implementation challenges related to scale-up). We also independently varied bNAb uptake, pediatric ART costs (0·5x-2·0x), cost of ascertaining high-risk status ($0-$31·66), mean breastfeeding duration (setting- and risk-group-specific ranges), maternal ART coverage during pregnancy (60-100%), maternal knowledge of acute HIV infection (25-95% during pregnancy, 0-15% per month postpartum), infant antiretroviral prophylaxis efficacy (40-90% intrapartum and 58-90% postnatally), postpartum maternal HIV incidence (0·5x-2·0x), and postnatal transmission risk (0·5x-2·0x) (see Supplementary Table 3 for range justifications). These sensitivity analyses were meant to reflect an evolving HIV epidemic and care landscape to account for potential advances in other prevention and detection strategies (e.g., lower maternal HIV prevalence or increased maternal testing and retesting). We also separately examined the effects of modeling: a) bNAbs having no protective efficacy against intrapartum transmission to capture scenarios in which women may deliver at home and not arrive to postpartum care early enough to prevent intrapartum transmission, b) offering bNAbs to infants without also providing oral antiretroviral prophylaxis, and c) reducing the cost-effectiveness threshold to 20% GDP per capita. In multivariate sensitivity analyses, we simultaneously varied bNAb efficacy, effect duration, and cost, and we separately varied bNAb efficacy, cost, and maternal HIV prevalence throughout sub-nationally observed ranges (1-5% in Côte d’Ivoire, 15-45% in South Africa, and 5-25% in Zimbabwe).

## RESULTS

### Clinical outcomes

With standard-of-care maternal ART and infant antiretroviral prophylaxis, our model projects a total vertical HIV transmission rate of 8·4%, 4·4%, and 9·5% in Côte d’Ivoire, South Africa, and Zimbabwe, respectively, similar to UNAIDS projections (Table 2, Supplementary Table 4). Depending on the bNAb dosing approach, bNAb strategies that target infants with known HIV exposure (*HIVE* strategies) would prevent 7-26% additional pediatric HIV infections compared to the standard-of-care alone, reducing the cumulative vertical HIV transmission rate to between 6·3-7·8%, 3·3-3·8%, and 6·9-8·4% in Côte d’Ivoire, South Africa, and Zimbabwe, respectively (Table 2, Figures 1-2). Expanding the target population for bNAbs to all infants (*ALL* strategies) was projected to further improve clinical outcomes, preventing 10-42% additional annual pediatric HIV infections compared to the standard-of-care alone, reducing the cumulative vertical HIV transmission rate to between 5·5-7·5%, 2·5-3·5%, and 5·7-7·9% in Côte d’Ivoire, South Africa, and Zimbabwe, respectively.

**Table 2.**
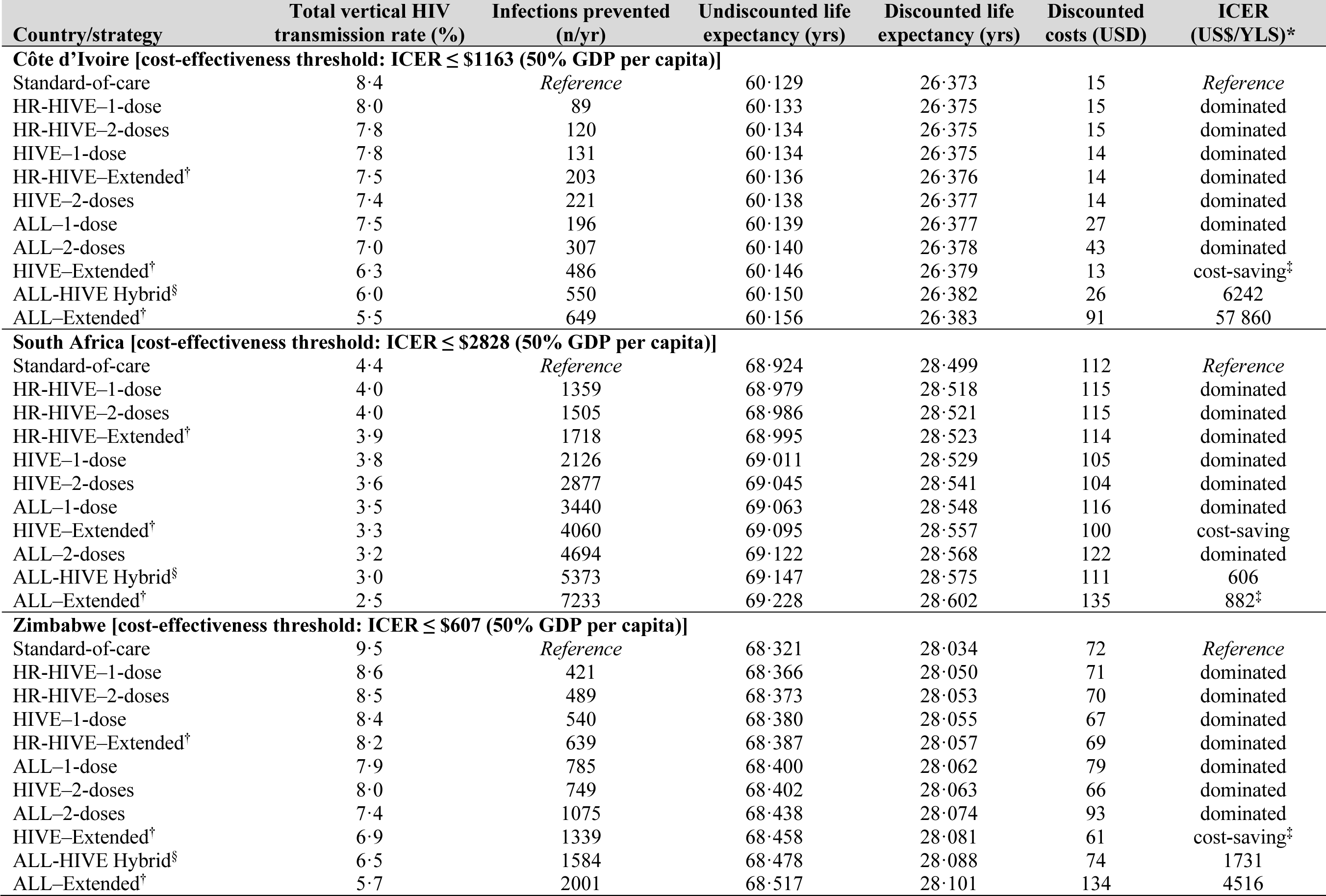
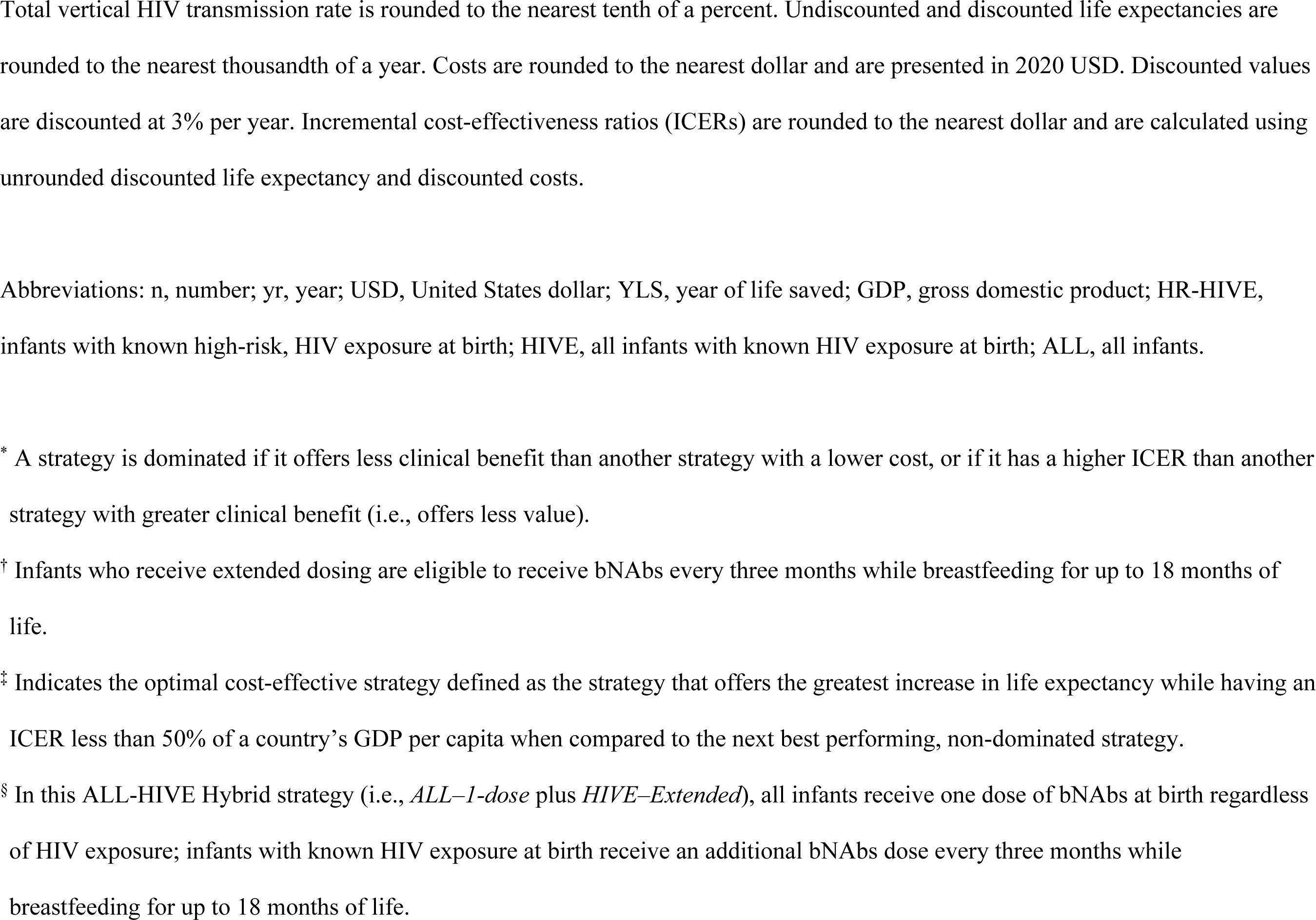
Base case clinical and economic outcomes of bNAb infant prophylaxis programs, by country.

| Country/strategy | Total vertical HIV transmission rate (%) | Infections prevented (n/yr) | Undiscounted life expectancy (yrs) | Discounted life expectancy (yrs) | Discounted costs (USD) | ICER (US\$/YLS)* |
| --- | --- | --- | --- | --- | --- | --- |
| **Côte d'Ivoire [cost-effectiveness threshold: ICER ≤ \$1163 (50% GDP per capita)]** | | | | | | |
| Standard-of-care | 8.4 | *Reference* | 60.129 | 26.373 | 15 | *Reference* |
| HR-HIVE-1-dose | 8.0 | 89 | 60.133 | 26.375 | 15 | dominated |
| HR-HIVE-2-doses | 7.8 | 120 | 60.134 | 26.375 | 15 | dominated |
| HIVE-1-dose | 7.8 | 131 | 60.134 | 26.375 | 14 | dominated |
| HR-HIVE-Extended<sup>†</sup> | 7.5 | 203 | 60.136 | 26.376 | 14 | dominated |
| HIVE-2-doses | 7.4 | 221 | 60.138 | 26.377 | 14 | dominated |
| ALL-1-dose | 7.5 | 196 | 60.139 | 26.377 | 27 | dominated |
| ALL-2-doses | 7.0 | 307 | 60.140 | 26.378 | 43 | dominated |
| HIVE-Extended<sup>†</sup> | 6.3 | 486 | 60.146 | 26.379 | 13 | cost-saving<sup>‡</sup> |
| ALL-HIVE Hybrid<sup>§</sup> | 6.0 | 550 | 60.150 | 26.382 | 26 | 6242 |
| ALL-Extended<sup>†</sup> | 5.5 | 649 | 60.156 | 26.383 | 91 | 57 860 |
| **South Africa [cost-effectiveness threshold: ICER ≤ \$2828 (50% GDP per capita)]** | | | | | | |
| Standard-of-care | 4.4 | *Reference* | 68.924 | 28.499 | 112 | *Reference* |
| HR-HIVE-1-dose | 4.0 | 1359 | 68.979 | 28.518 | 115 | dominated |
| HR-HIVE-2-doses | 4.0 | 1505 | 68.986 | 28.521 | 115 | dominated |
| HR-HIVE-Extended<sup>†</sup> | 3.9 | 1718 | 68.995 | 28.523 | 114 | dominated |
| HIVE-1-dose | 3.8 | 2126 | 69.011 | 28.529 | 105 | dominated |
| HIVE-2-doses | 3.6 | 2877 | 69.045 | 28.541 | 104 | dominated |
| ALL-1-dose | 3.5 | 3440 | 69.063 | 28.548 | 116 | dominated |
| HIVE-Extended<sup>†</sup> | 3.3 | 4060 | 69.095 | 28.557 | 100 | cost-saving |
| ALL-2-doses | 3.2 | 4694 | 69.122 | 28.568 | 122 | dominated |
| ALL-HIVE Hybrid<sup>§</sup> | 3.0 | 5373 | 69.147 | 28.575 | 111 | 606 |
| ALL-Extended<sup>†</sup> | 2.5 | 7233 | 69.228 | 28.602 | 135 | 882<sup>‡</sup> |
| **Zimbabwe [cost-effectiveness threshold: ICER ≤ \$607 (50% GDP per capita)]** | | | | | | |
| Standard-of-care | 9.5 | *Reference* | 68.321 | 28.034 | 72 | *Reference* |
| HR-HIVE-1-dose | 8.6 | 421 | 68.366 | 28.050 | 71 | dominated |
| HR-HIVE-2-doses | 8.5 | 489 | 68.373 | 28.053 | 70 | dominated |
| HIVE-1-dose | 8.4 | 540 | 68.380 | 28.055 | 67 | dominated |
| HR-HIVE-Extended<sup>†</sup> | 8.2 | 639 | 68.387 | 28.057 | 69 | dominated |
| ALL-1-dose | 7.9 | 785 | 68.400 | 28.062 | 79 | dominated |
| HIVE-2-doses | 8.0 | 749 | 68.402 | 28.063 | 66 | dominated |
| ALL-2-doses | 7.4 | 1075 | 68.438 | 28.074 | 93 | dominated |
| HIVE-Extended<sup>†</sup> | 6.9 | 1339 | 68.458 | 28.081 | 61 | cost-saving<sup>‡</sup> |
| ALL-HIVE Hybrid<sup>§</sup> | 6.5 | 1584 | 68.478 | 28.088 | 74 | 1731 |
| ALL-Extended<sup>†</sup> | 5.7 | 2001 | 68.517 | 28.101 | 134 | 4516 |

Total vertical HIV transmission rate is rounded to the nearest tenth of a percent. Undiscounted and discounted life expectancies are rounded to the nearest thousandth of a year. Costs are rounded to the nearest dollar and are presented in 2020 USD. Discounted values are discounted at 3% per year. Incremental cost-effectiveness ratios (ICERs) are rounded to the nearest dollar and are calculated using unrounded discounted life expectancy and discounted costs.
Abbreviations: n, number; yr, year; USD, United States dollar; YLS, year of life saved; GDP, gross domestic product; HR-HIVE, infants with known high-risk, HIV exposure at birth; HIVE, all infants with known HIV exposure at birth; ALL, all infants.
\* A strategy is dominated if it offers less clinical benefit than another strategy with a lower cost, or if it has a higher ICER than another strategy with greater clinical benefit (i.e., offers less value).
† Infants who receive extended dosing are eligible to receive bNAbs every three months while breastfeeding for up to 18 months of life.
‡ Indicates the optimal cost-effective strategy defined as the strategy that offers the greatest increase in life expectancy while having an ICER less than 50% of a country's GDP per capita when compared to the next best performing, non-dominated strategy.
§ In this ALL-HIVE Hybrid strategy (i.e., *ALL-1-dose* plus *HIVE-Extended*), all infants receive one dose of bNAbs at birth regardless of HIV exposure; infants with known HIV exposure at birth receive an additional bNAbs dose every three months while breastfeeding for up to 18 months of life.

**Figure 1.**
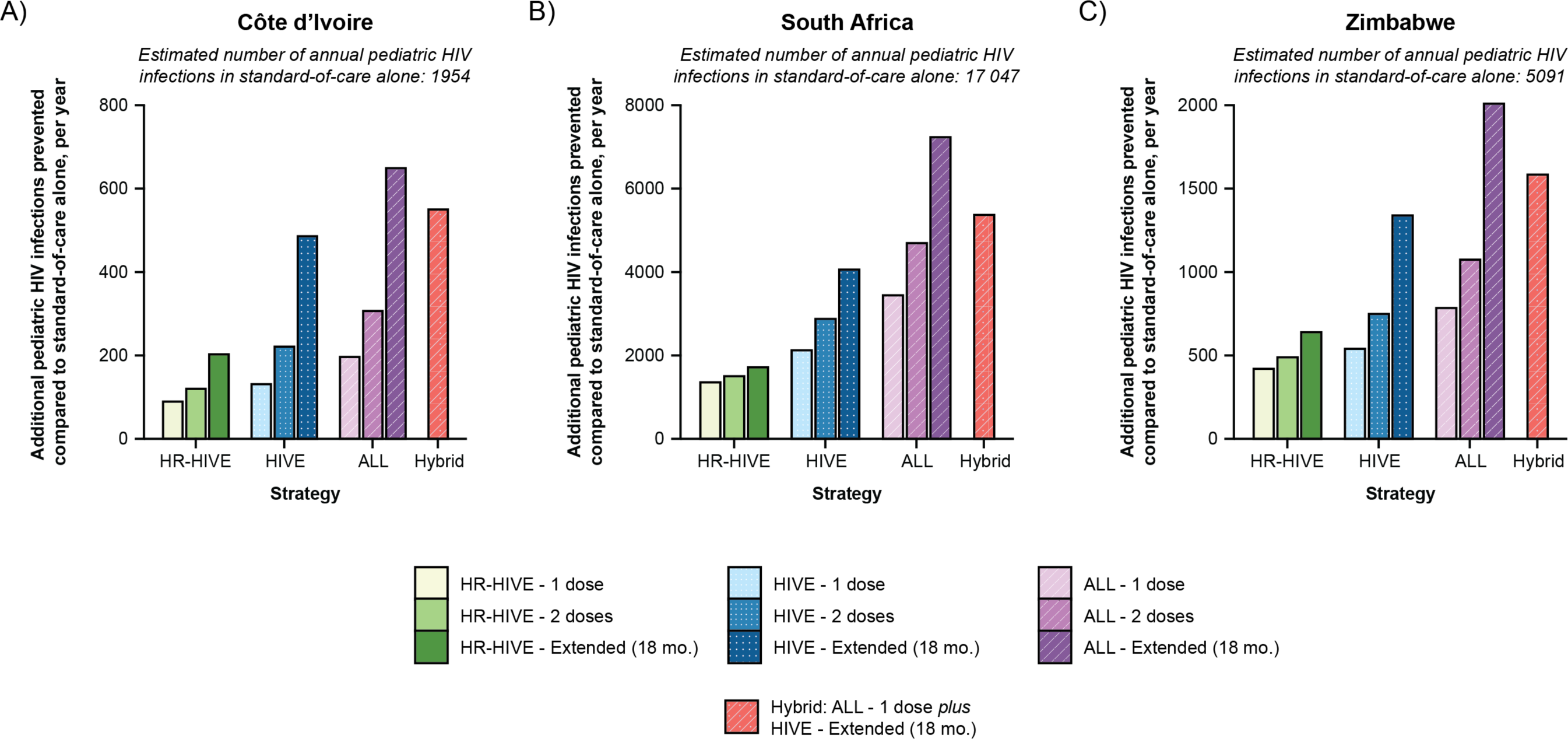
Base case number of model-projected HIV infections prevented by bNAb strategies, by country. The number of model-estimated pediatric HIV infections (i.e., before 36 months) prevented by each broadly neutralizing antibody (bNAb) strategy, compared to the standard-of-care alone, is presented for Côte d’Ivoire (Panel A), South Africa (Panel B), and Zimbabwe (Panel C). The axes on each panel are different due to the different number of projected infections in the standard-of-care alone. Abbreviations: HIVE, bNAb strategy targeting infants with known HIV exposure; ALL, bNAb strategy targeting all infants; mo., month.

**Figure 2.**
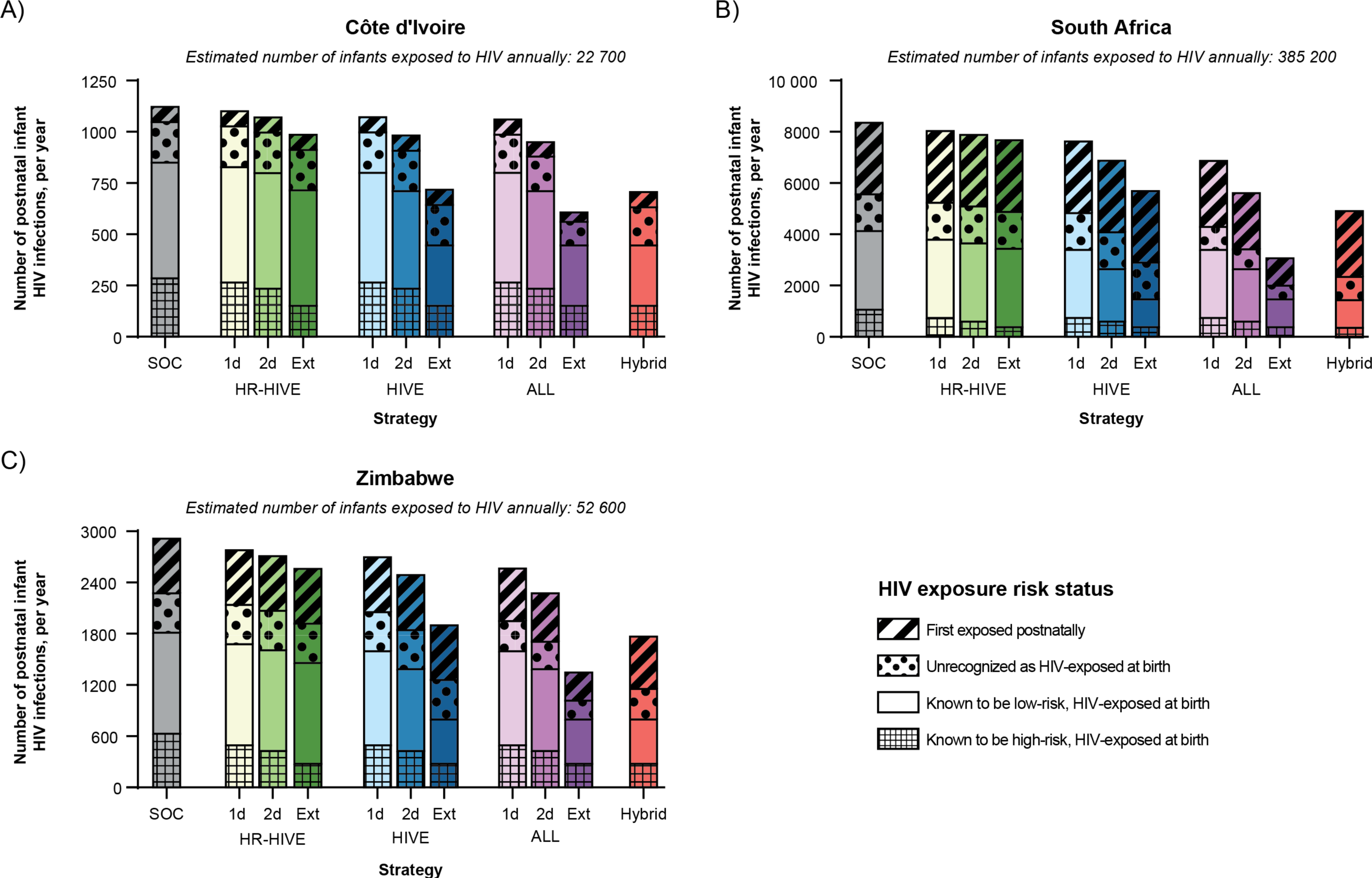
Breakdown of base case model-projected postnatal vertical HIV transmissions occurring in each exposure group, by country. The number of postnatal pediatric HIV infections (i.e., before 36 months) occurring in each of four exposure groups assessed at birth (i.e., among infants who are initially unexposed, infants who are unrecognized to be HIV-exposed, infants with known low-risk exposure, or infants with known high-risk exposure) is presented for Côte d’Ivoire (Panel A), South Africa (Panel B), and Zimbabwe (Panel C). The “Hybrid” strategy (i.e., *ALL–1-dose* plus *HIVE–Extended*) offers one dose of broadly neutralizing antibodies (bNAbs) to all infants with or without HIV exposure, and an additional bNAbs dose every three months to infants with known HIV exposure at birth for up to 18 months while breastfeeding. The estimated number of infants at risk of HIV acquisition in each exposure group varies by country. In Côte d’Ivoire, an estimated 6500 infants have known, high-risk HIV exposure at birth; 14 700 infants have known, low-risk HIV exposure at birth; 1100 infants have unrecognized HIV exposure at birth; and 400 infants are first exposed postnatally. In South Africa, an estimated 65 600 infants have known, high-risk HIV exposure at birth; 294 500 infants have known, low-risk HIV exposure at birth; 18 400 infants have unrecognized HIV exposure at birth; and 9700 infants are first exposed postnatally. In Zimbabwe, an estimated 11 500 infants have known, high-risk HIV exposure at birth; 35 400 infants have known, low-risk HIV exposure at birth; 3600 infants have unrecognized HIV exposure at birth; and 2100 infants are first exposed postnatally. Abbreviations: HR-HIVE, bNAb strategy targeting infants with known, high-risk HIV exposure; HIVE, bNAb strategy targeting infants with known HIV exposure; ALL, bNAb strategy targeting all infants; SOC, standard-of-care; 1d, one dose; 2d, two dose; Ext, extended; mo., month.

### Cost-effectiveness outcomes

In all three countries, *HIVE–Extended* was projected to be cost-saving compared to the standard-of-care alone: it would increase life expectancy while reducing lifetime HIV-related costs (Table 2). While *HR-HIVE* strategies would improve clinical outcomes at lower or similar costs compared to the standard-of-care, they would result in greater lifetime HIV-related costs and smaller life expectancy gains than *HIVE–Extended* and are, thus, dominated. Using a 50% GDP per capita cost-effectiveness threshold, of the three modeled countries, one of the *ALL* strategies would only be cost-effective in South Africa, where maternal HIV prevalence and incidence is currently the highest: *ALL–Extended* would be the cost-effective strategy in South Africa (ICER: $882/YLS) while *HIVE–Extended* would be the cost-effective strategy and cost-saving in Côte d’Ivoire and Zimbabwe (Table 2).

### Univariate sensitivity analyses

In all settings examined, at least one bNAb strategy remained cost-effective relative to the standard-of-care across all sensitivity analyses throughout plausible ranges (Supplementary Figures 6-7). However, which specific bNAb strategy would be cost-effective in each country was most sensitive to changes in bNAb product characteristics (i.e., efficacy, cost, and effect duration) and decreases in maternal knowledge of HIV status (Supplementary Figure 6, Supplementary Tables 9-11 and 20-21).

#### BNAb product characteristics (cost and efficacy)

A bNAb strategy targeting all infants with known HIV exposure at birth was cost-effective at all evaluated bNAb efficacies (10%-100%) and costs ($10-$100/dose). The upper cost range was intended to capture scenarios in which increased product costs, implementation challenges, and scale-up may lead to increased total program costs. In Côte d’Ivoire, *HIVE–Extended* would remain cost-effective even if bNAb efficacy were reduced to 20% or bNAb cost were increased to $100 (Figure 3). In South Africa, the cost-effective bNAb strategy would continue to target all infants (*ALL–Extended* or *ALL–1-dose plus HIVE–Extended*) unless bNAb efficacy were reduced to ≤20% or bNAb cost were increased to $100 per dose, in which case *HIVE–Extended* would become cost-effective. In Zimbabwe, the cost-effective strategy would remain *HIVE–Extended* unless bNAb efficacy were reduced to 10% or bNAb cost were increased to $100 per dose, in which case *HIVE–1-dose* would become cost-effective; the cost-effective strategy would change to *ALL–1-dose plus HIVE–Extended* if bNAb cost were reduced to $5 per dose.

**Figure 3.**
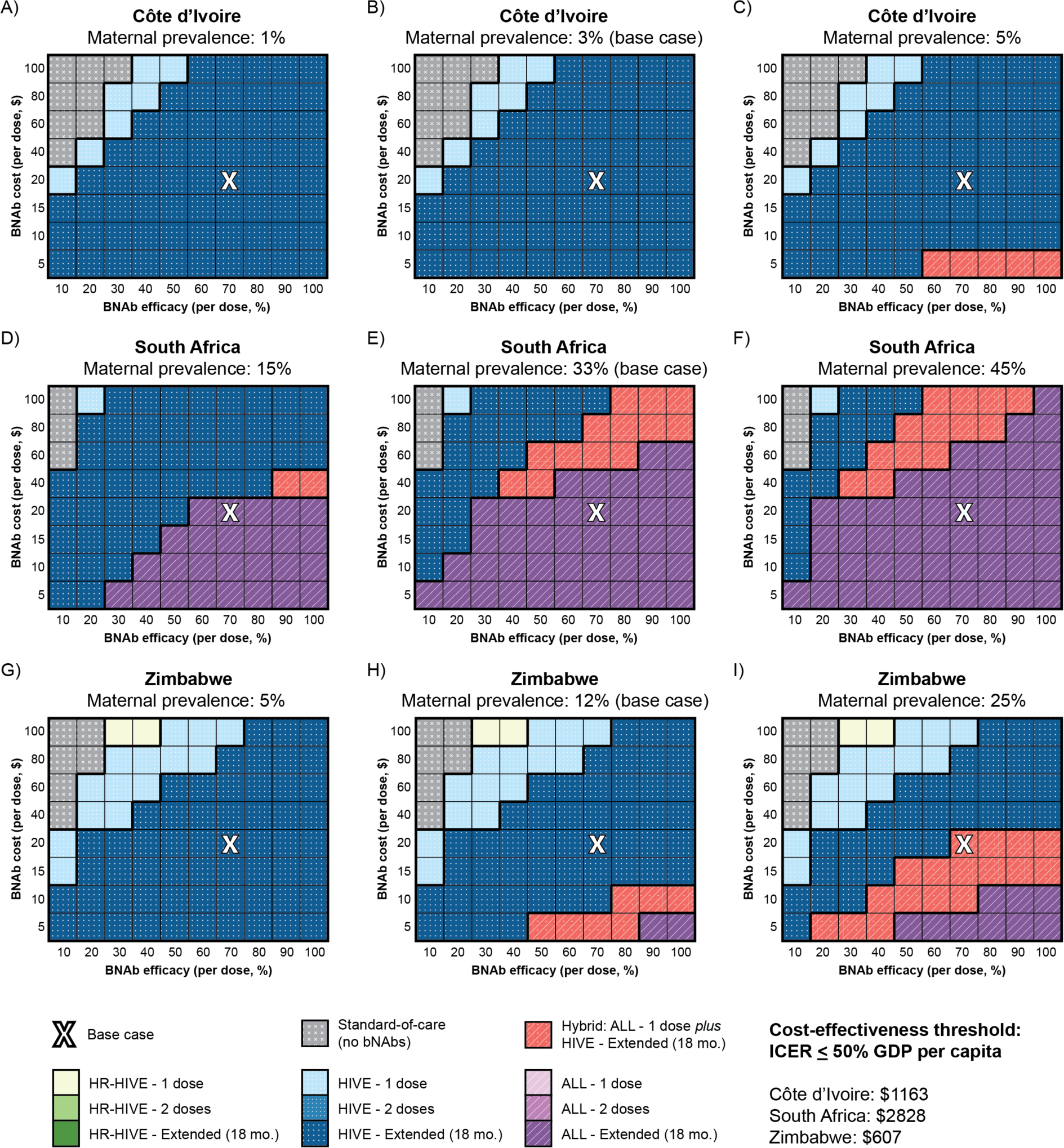
Cost-effectiveness results of a three-way sensitivity analysis assessing bNAb efficacy, cost, and maternal HIV prevalence/incidence, by country. We conducted a three-way sensitivity analysis on the impact of bNAb cost per dose (vertical axis), efficacy per dose (horizontal axis), and maternal HIV prevalence (left to right panels) on the cost-effective bNAb strategy in the three countries examined: Côte d’Ivoire (Panels A-C), South Africa (Panels D-F), and Zimbabwe (Panels G-I). To simulate low (left-most panels) and high (right-most panels) maternal prevalence scenarios, we used the lowest and highest subnational maternal prevalence point estimates reported in survey data as inputs.^16,17,42^ Maternal prevalence at the time of delivery is inclusive of chronic HIV infection and incident infection during pregnancy. Postpartum maternal HIV incidence was also decreased (Côte d’Ivoire: 0.001%/month, South Africa: 0.107%/month, Zimbabwe: 0.022%/month) or increased (Côte d’Ivoire: 0.005%/month, South Africa: 0.331%/month, Zimbabwe: 0.116%/month) proportional to the change in prevalence. BNAb strategies targeting only high-risk infants known to be HIV-exposed at birth (HR-HIVE) are depicted in shades of green. Strategies targeting all infants known to be HIV-exposed (HIVE) are depicted in shades of blue, and strategies targeting all infants irrespective of HIV exposure (ALL) are depicted in shades of purple. The hybrid strategy offering one dose of bNAbs at birth to all infants irrespective of HIV exposure and extended bNAb doses to infants known to be HIV-exposed (Hybrid) is depicted in red. Eligible infants would receive either one dose, two doses, or a dose of bNAbs every three months while breastfeeding for up to 18 months (i.e., extended dosing). The cost-effective bNAb strategy was defined as per the methods. The white “X” in each panel reflects the base case assumptions of 70% bNAb efficacy at a cost of $20/dose. Abbreviations: bNAb, broadly neutralizing antibody; mo., month.

#### Maternal knowledge of HIV status

If maternal knowledge of acute HIV acquisition during pregnancy were increased to 95% (up from base case of 56% in Côte d’Ivoire, 55% in South Africa, and 70% in Zimbabwe) and knowledge of acute HIV acquisition postpartum were increased to 15%/month (up from base case of 2%/month in Côte d’Ivoire, 9%/month in South Africa, and 5%/month in Zimbabwe)–a scenario reflecting scaled-up maternal HIV testing during pregnancy and retesting–the cost-effective strategy would not change in any of the three countries under base case assumptions about bNAb product characteristics (i.e., 70% efficacy, $20/dose cost, and 3-month effect duration) (Supplementary Table 20). Decreased maternal knowledge of acute HIV acquisition would change the cost-effective strategy to *ALL–1-dose* plus *HIVE–Extended* in Zimbabwe, but not Côte d’Ivoire, under base case assumptions about bNAb product characteristics (Supplementary Table 20).

#### Other univariate analyses

In all three countries, the cost-effective strategy was not sensitive to changes in ART costs, bNAb uptake, breastfeeding duration, cost of ascertaining high-risk status, maternal ART coverage during pregnancy, infant oral antiretroviral prophylaxis efficacy, postpartum maternal retention in care, and postnatal vertical HIV transmission risk over tested ranges (Supplementary Figure 7; Supplementary Tables 12-19 and 22). Evaluating strategies under the assumptions that bNAbs would have no efficacy against intrapartum transmission, that bNAbs prophylaxis would be implemented without associated oral antiretroviral prophylaxis, that no additional costs were associated with identifying high-risk infants in the *HR-HIVE* strategies, or that the cost-effectiveness threshold were reduced to 20% GDP per capita did not change the cost-effective strategy (Supplementary Figures 7-8; Supplementary Tables 6 and 19).

### Multivariate sensitivity analyses

If the maternal HIV prevalence and incidence were each reduced by over half (prevalence: 1·0% in Côte d’Ivoire, 15·0% in South Africa, and 5·0% in Zimbabwe; postpartum incidence: 0·001%/mo in Côte d’Ivoire, 0·107%/mo in South Africa, and 0·022% in Zimbabwe)–reflecting a scenario in which maternal HIV prevention were strengthened–the cost-effective strategy would not change in any of the three countries under base case assumptions about bNAb product characteristics (Supplementary Table 8).

BNAb strategies remained cost-effective in all three countries at most combinations of bNAb efficacy, effect duration, and cost (Supplementary Figure 9), and at most combinations of bNAb efficacy, bNAb cost, and maternal HIV prevalence/incidence (Figure 3). In Côte d’Ivoire, targeting bNAbs to infants with known HIV exposure at birth (*HIVE* strategies) would be cost-effective at most combinations of bNAb product characteristics and maternal prevalence/incidence. In South Africa, targeting bNAbs to all infants (*ALL* strategies) would be cost-effective at most tested combinations of bNAb product characteristics and maternal prevalence/incidence. In Zimbabwe, targeting bNAbs to infants with known HIV exposure at birth (*HIVE* strategies) would be cost-effective at most tested combinations of bNAb product characteristics and maternal prevalence/incidence, with *ALL–1-dose plus HIVE–Extended* and *ALL–Extended* becoming cost-effective at high efficacy and low cost combinations, particularly if maternal prevalence/incidence were higher than in the base case.

## DISCUSSION

We projected the clinical impact and cost-effectiveness of different strategies of bNAb HIV prophylaxis for infants in addition to the current WHO-recommended strategies to prevent vertical HIV transmission in three countries with high HIV burdens. We found that offering bNAb prophylaxis to all infants, irrespective of known HIV exposure, could prevent 10-42% more annual pediatric HIV infections than the standard-of-care in Côte d’Ivoire, South Africa, and Zimbabwe. However, cost-effectiveness varies by setting. While exact bNAb product characteristics are unknown, our projections suggest that offering bNAbs to all infants (i.e., *ALL* strategies) would be cost-effective in settings with high maternal HIV prevalence and incidence, as in South Africa, throughout a wide range of plausible bNAb costs and efficacies. Offering bNAbs to all infants in settings like Côte d’Ivoire and Zimbabwe, where maternal HIV prevalence and GDP per capita are lower, would not be cost-effective at 70% bNAb efficacy and $20/dose cost, using a 50% GDP per capita threshold for cost-effectiveness. However, we found that offering bNAbs specifically to infants with known HIV exposure in these settings, regardless of low- or high-risk, throughout the first 18 months of breastfeeding, would be cost-effective, even if efficacy were as low as 20% and costs were as high as $80/dose.

Acute maternal HIV infection during pregnancy and breastfeeding confers a high risk of vertical HIV transmission and may be more likely to go undiagnosed in the absence of repeated maternal HIV testing. In contrast to existing maternal ART and infant antiretroviral prophylaxis approaches, a program offering universal bNAb infant prophylaxis regardless of HIV exposure, akin to a vaccine, could protect infants who are unrecognized as HIV-exposed at birth or who become HIV-exposed postnatally, addressing a critical gap in the perinatal HIV prevention continuum. Accordingly, we found that bNAb strategies targeting all infants would offer the greatest value in settings where maternal HIV detection is suboptimal and/or where maternal HIV incidence during pregnancy or breastfeeding is high, such as in South Africa. Our results suggest that offering bNAbs to all infants may be cost-effective in national or subnational settings like those we examined when maternal HIV prevalence exceeds approximately 15%, bNAbs cost ≤$20/dose, and bNAbs have ≥60% efficacy.

BNAb infant prophylaxis may hold additional clinical advantages beyond having long-acting effectiveness against vertical HIV transmission. In contrast to antiretroviral prophylaxis, bNAbs are unlikely to contribute to pre-treatment drug resistance against first-line ART among infants who acquire HIV. Similarly, while their safety data in infants is limited,^4,15^ bNAbs may carry less risk of hematologic toxicities among neonates compared to antiretrovirals.^31,32^ Furthermore, subcutaneous bNAb prophylaxis would be less sensitive than infant antiretroviral prophylaxis to developmental changes in metabolism, making bNAbs ideal protective candidates for preterm infants who are more likely to experience challenges absorbing and metabolizing antiretroviral prophylaxis.^33^ To be conservative with respect to the potential clinical benefit of bNAb prophylaxis in this model-based analysis, we did not account for these factors, but they may make bNAbs even more cost-effective compared to existing infant HIV prevention approaches.

Our results suggest bNAbs would significantly reduce vertical HIV transmission, but they would not eliminate transmission entirely. Even if all infants were offered bNAbs throughout breastfeeding (i.e., *ALL*–*Extended*), total vertical transmission was modeled to be 5·5%, 2·5%, and 5·7% in Côte d’Ivoire, South Africa, and Zimbabwe, respectively. Most of these transmissions were due to intrauterine transmission, for which bNAbs would have no impact, or intrapartum transmission, for which bNAbs may have limited protective coverage. The modeled clinical benefit of bNAbs in our analysis was also limited by our assumption about their uptake. Efforts to increase bNAb uptake would result in additional clinical benefit and cost-effectiveness. Our results suggest that bNAbs would help make significant progress toward achieving elimination of vertical HIV transmission, but full elimination will likely require a multi-pronged approach including improved maternal HIV testing, ART coverage, support for ART adherence, and retention in care throughout pregnancy and breastfeeding.

This analysis may help directly inform targets for bNAb product characteristics during development. However, a national bNAb infant prophylaxis program would also need to factor implementation considerations. This analysis found that the cost-effective target population for bNAb implementation would largely depend on maternal HIV prevalence and postpartum incidence. As such, a single bNAb implementation strategy across national settings may not be recommended from a cost-effectiveness standpoint. However, global policy-making bodies could consider simplifying recommendations on bNAb implementation strategies based on key metrics, including national or subnational estimates of maternal HIV prevalence and incidence. Additionally, the modeled clinical impact of bNAb implementation strategies reflect introduction into existing maternal-child health care programs and HIV care cascades. For example, to avoid overestimating the clinical benefit of bNAbs, the birth bNAbs dose was modeled to be offered only to eligible infants born in a healthcare facility and the receipt of subsequent bNAb doses were based on setting-specific uptake of other routine infant immunizations. Further, the modeled clinical impact did not depend on perfect HIV risk ascertainment at birth but rather on existing early infant diagnosis programs. The cost-effective strategy in each of the three settings and across many sensitivity analyses would either offer bNAbs to infants with known HIV exposure at birth or all infants regardless of known HIV exposure at birth. These sorts of approaches could potentially simplify programmatic implementation of infant postnatal prophylaxis insofar as they would not necessitate distinction between high-risk and non-high-risk HIV exposure based on maternal ART status or maternal HIV viral load which has proven to be challenging to determine with respect to implementation of the current standard-of-care oral infant prophylaxis regimen.

As the eligible population is scaled-up, it is possible that implementation costs decrease initially due to economies of scale (i.e., more efficient production and delivery) and later increase as it may get more difficult to reach all infants. Additional potential implementation considerations with cost implications could include the challenges of administering bNAbs at timepoints not aligned with existing routine immunization schedules, requirements for multiple injections per visit if bNAbs are not co-formulated, and additional requirements for infant HIV testing (i.e., if there were a potential interaction of bNAbs on infant HIV RNA test sensitivity or antibody test specificity, requiring additional clinic visits and infant HIV testing). To account for the costs of these and other potential implementation challenges, we included sensitivity analyses on bNAb cost. Even in scenarios in which these implementation factors lead to substantially increased total bNAb program costs (up to $100/dose) bNAb strategies would continue to hold good value.

We found that providing bNAb infant HIV prophylaxis in addition to the standard-of-care offers good value relative to current HIV prevention and maternal-child health interventions.^34^ Our results suggest that in all three study settings, offering bNAb prophylaxis to all known HIV-exposed infants (*HIVE* strategies) would be cost-saving relative to standard-of-care maternal ART and infant prophylaxis as currently implemented, as the costs of bNAb prophylaxis would be offset by averting subsequent costs and health consequences from pediatric HIV infections. This finding is in contrast to most economic evaluations of pre-exposure prophylaxis (PrEP) in adults, which have found that adult PrEP is cost-effective for select populations but is unlikely to be cost-saving.^34^ In South Africa, we found that a program offering bNAbs to all infants at birth and throughout breastfeeding might not be cost-saving but would be cost-effective with an ICER of $882/YLS. This ICER compares favorably to published ICERs of other maternal-child health interventions in sub-Saharan Africa, including antenatal influenza vaccination (ICER: $4689/QALY gained),^35^ PrEP in pregnancy (ICER: $965/DALY averted),^36^ and birth EID testing (ICERs: $903-$2900/YLS).^37^ However, there is growing concern that the often-used 100% GDP per capita cost-effectiveness threshold may be inappropriate in many resource-limited settings.^38^ As such, we used a more conservative 50% GDP per capita cost-effectiveness threshold in the main analysis (Supplementary Methods). Using an even lower threshold of 20% GDP per capita did not change the cost-effective strategy under base case assumptions of bNAb product characteristics (Supplementary Figure 8).

It is important to note that cost-effectiveness is only one factor in health policy decision-making. Notably, while most hybrid strategies offering different bNAb dosing approaches to infants based on HIV exposure status would be dominated from a cost-effectiveness standpoint (i.e., provide a similar clinical benefit at only slightly greater cost than a non-hybrid strategy), they may be of interest to policy makers due to potentially improved feasibility, acceptability, or equity (Supplementary Table 5). As a complement to the current analysis, further work is currently underway to understand the feasibility and acceptability of bNAbs implementation strategies as part of the CELEBRATE and IMPAACT 2037 studies. While a cost-effectiveness analysis provides useful information on the value of bNAb infant prophylaxis relative to the current standard-of-care, a budget impact analysis to better assess affordability may be useful.

This analysis has several limitations. First, bNAb products are currently under clinical investigation, so the characteristics of a licensed bNAb combination product for infant HIV prophylaxis in clinical care settings remain unknown. Despite this uncertainty, our sensitivity analyses suggest that a bNAb combination product would offer good value in sub-Saharan African settings through a broad range of plausible efficacies and costs. Second, we did not evaluate the potential value of universal bNAb prophylaxis in the context of maternal or infant long-acting injectable antiretrovirals (LA-ARVs); however, any bNAb product coming to market is likely to have a more favorable toxicity profile than small molecular antiretrovirals, particularly among preterm infants with impaired metabolism and the timelines for widespread access to LA-ARVs across study settings remain unclear.^33^ Third, we did not explore how bNAb programs may best integrate with existing or scaled-up maternal or infant HIV testing results at the time of each potential bNAb dose administration, as these complexities were beyond the scope of the analysis. Fourth, we did not assess the impact of bNAb prophylaxis on HIV test results in early infant diagnosis programs which may influence the final identification of an infant’s HIV status. Finally, we did not explicitly consider health equity in this analysis. Conclusions regarding the cost-effective strategy for bNAb implementation in each setting may differ depending upon prioritization of social distribution of health, equity considerations, and affordability of HIV programs with different levels of donor funding and government investment.

If shown to be safe and effective, adding extended bNAb infant prophylaxis to the current standard-of-care would substantially reduce pediatric HIV infections and help make important progress toward eliminating vertical HIV transmission in settings like Côte d’Ivoire, South Africa, and Zimbabwe. Offering bNAbs to all infants with or without known HIV exposure could be cost-effective in countries or subnational regions with high maternal HIV prevalence and incidence. The projected clinical impact and cost-effectiveness of bNAb infant prophylaxis may serve as potential benchmarks for target bNAb characteristics and costs and should motivate further research into the clinical safety, pharmacokinetics, efficacy, and implementation considerations of bNAb combinations as a potentially cost- and life-saving HIV prevention intervention.

## Data Availability

All data produced in the present study are available upon reasonable request to the authors.

