## Supplementary Methods, Figures, and Tables for "Cost-effectiveness of broadly neutralizing antibodies for infant HIV prophylaxis in settings with high HIV burdens: a simulation modeling study"

**Appendix**

### Supplementary Methods

These supplementary methods provide additional details regarding modeling methodology, input specification, and derivation of key parameters. The Cost-Effectiveness of Preventing AIDS Complications – Pediatric (CEPAC-P) model has previously been validated and additional information is available in previously published works.[1–11] For further details regarding the mathematical formulas used in the model, model flowcharts, and opportunities for collaboration, we direct readers to the CEPAC website: [https://mpec.massgeneral.org/cepac-model/](https://mpec.massgeneral.org/cepac-model/).

#### Cohort weighting

For each country, we simulated four sub-cohorts of infants with varying risks of HIV exposure assessed at the time of birth:

1) infants with known, high-risk HIV exposure;
2) infants with known, low-risk HIV exposure;
3) infants with unrecognized HIV exposure; and
4) infants initially unexposed to HIV (but who may become exposed postnatally).

We used World Health Organization (WHO) criteria to define high-risk (i.e., mother was not on antiretroviral therapy [ART] in pregnancy, mother was on ART in pregnancy but had a viral load [VL] >1000 copies/mL near delivery, or mother was on ART for <4 weeks prior to delivery) and low-risk (i.e., mother was on ART and had VL ≤1000 copies/mL near delivery) exposure.[12]

Together, these four sub-cohorts include all infants born in a country. To estimate the proportion of infants within each sub-cohort, we used published data on country-specific estimates of maternal HIV prevalence/incidence during pregnancy, knowledge of HIV status, ART uptake, and probability of virologic suppression to populate the following equations:

$$\text{Known, high-risk proportion} = \left(\textit{\% of mothers known to have chronic HIV who were not on ART during pregnancy}\right) + \left(\textit{\% of mothers known to have chronic HIV who were on ART with a VL} \geq 1000 \textit{ copies/mL}\right) + \left(\textit{\% of mothers known to have been acutely infected with HIV during pregnancy}\right)$$

$$\text{Known, low-risk proportion} = \left(\textit{\% of mothers known to have chronic HIV who were on ART with a VL} < 1000 \textit{ copies/mL}\right)$$

$$\text{Unknown HIV-exposed proportion} = \left(\textit{\% of mothers with unknown chronic HIV during pregnancy}\right) + \left(\textit{\% of mothers with unknown acute HIV during pregnancy}\right)$$

$$\text{Unexposed proportion} = 1 - \left(\textit{known, high-risk proportion}\right) - \left(\textit{known, low-risk proportion}\right) - \left(\textit{unknown HIV-exposed proportion}\right)$$

We used the proportion of infants within each of the four sub-cohorts to weight model outcomes and scale results representative of all infants born in a country.

#### Additional modeled strategies

To determine the potential impact of broadly neutralizing antibody (bNAb) infant prophylaxis, we modeled the standard-of-care alone and various strategies offering bNAbs, in addition to the standard-of-care, to subsets of infants. A bNAb strategy targeted one of three nested populations of infants: 1) infants with known, high-risk

HIV exposure at birth (*HR-HIVE* strategies); 2) infants with any known HIV exposure at birth (*HIVE* strategies); or 3) all infants regardless of known HIV exposure (*ALL* strategies). Infants in the target population were offered one of three dosing approaches: 1) one bNAbs dose at birth (*1-dose*), 2) one bNAbs dose at birth and one dose at three months (*2-doses*), or 3) one dose of bNAbs at birth and one bNAbs dose every three months thereafter for up to 18 months while breastfeeding (*Extended*). The combination of a target population and a dosing approach comprised a bNAb strategy. For example, *ALL–1-dose* would be the bNAb strategy in which all infants, regardless of known HIV exposure, are offered one dose of bNAbs at birth.

In the primary analysis, we mainly examined strategies in which all infants in the target population received the same dosing approach. However, we also examined hybrid strategies in which subsets of the target population received different dosing approaches. For example, *ALL–1-dose plus HIVE–Extended* would offer one dose of bNAbs to all infants at birth and an additional dose of bNAbs to infants with known HIV exposure every three months for up to 18 months while breastfeeding. With the exception of *ALL–1-dose plus HIVE–Extended* all other hybrid strategies did not hold comparable value to non-hybrid strategies and are, thus, only presented in the appendix (Supplementary Table 5).

#### Infant HIV prophylaxis model structure

The infant HIV prophylaxis module (Supplemental Figure 2) is embedded within the CEPAC-P model and allows the user to simulate up to four concurrent and independent lines of HIV prophylaxis. The influence of infant HIV prophylaxis on reducing intrapartum (IP) transmission is reflected as a one-time multiplier on the IP component of the overall intrauterine (IU)/IP transmission risks at birth. For all months after birth, children who are breastfeeding, have not had a prior positive HIV test, and who do not have an active dose of prophylaxis from a prior month face optional user-specified prophylaxis eligibility criteria based on their age, maternal characteristics, and early infant diagnosis (EID) test results.

If the child meets eligibility criteria for that line of prophylaxis, they then face a probability of access and adherence to prophylaxis each month. If the child receives a dose of prophylaxis, an "efficacy multiplier" is applied to reduce postnatal transmission for a user-specified number of months during which the prophylaxis dose is active. Irrespective of efficacy, if a child receives prophylaxis in a given month, they also experience a probability of mild and/or severe drug toxicity in the month the dose was administered. Every toxicity event also carries a toxicity cost; when a severe toxicity is encountered, it also carries a probability of death in that month and triggers the prophylaxis regimen to be permanently stopped.

#### Oral infant prophylaxis regimens and efficacy

In the standard-of-care and in all bNAb strategies, infants with known HIV exposure were eligible for oral infant prophylaxis per WHO recommendations (i.e., six weeks of daily nevirapine [NVP] for infants who are low-risk, HIV-exposed and 12 weeks of daily zidovudine [ZDV] plus nevirapine for infants who are high-risk, HIV-exposed).[12] With conflicting data on the efficacy of one vs. multi-drug infant prophylaxis regimens, we modeled the same relative efficacy of both regimens.[13]

Modeled perinatal transmission risks among women on ART in pregnancy were largely derived from studies in which infants received short courses of antiretroviral prophylaxis after birth. Therefore, we did not model additional reductions in perinatal transmission among infants born to mothers on ART in pregnancy. However, among infants born to women who did not receive ART in pregnancy, we modeled a 69% relative reduction in the intrapartum component of perinatal transmission with oral infant prophylaxis based on data from the Post-Exposure Prophylaxis of Infants (PEPI) trial.[14] In the PEPI study, the cumulative HIV acquisition rate at six weeks among breastfeeding infants who received a single NVP dose plus one week of ZDV was 5·10% compared to 1·58% among infants who received control plus extended dual prophylaxis (NVP + ZDV) (relative risk [RR]: 0·31).[14] While transmission by six weeks may include some early postnatal transmission, we applied this risk reduction to the intrapartum component of vertical transmission risks for high-risk infants in the model, to be conservative with respect to the relative incremental benefits of bNAbs.

We also modeled a 71% decrease in postnatal transmission risk with oral infant prophylaxis based on an analysis of pooled individual data from five randomized trials of infant NVP prophylaxis demonstrating an adjusted hazard ratio (in time-varying analyses) of 0·29 (95% CI: 0·20-0·42) for vertical transmission while NVP was administered.[15]

#### BNAb efficacy and duration

Anti-HIV bNAbs, such as VRC01, VRC01-LS, and VRC07-523LS, have demonstrated safety, tolerability, and favorable pharmacokinetics in infants.[16–18] Current bNAbs under investigation for use as infant HIV prophylaxis include:

| Study | BNAb(s) investigated | Target site | Trial type | Included populations | Trial status |
|---|---|---|---|---|---|
| IMPAACT P1112 | VRC01<br><br>VRC01LS<br><br>VRC07-523LS | CD4-binding site<br><br>CD4-binding site<br><br>CD4-binding site | Phase 1 study on safety and pharamacokinetics | Infants with HIV exposure | Completed |
| PedMAB | VRC07-523LS *plus* CAP256V2LS | CD4-binding site<br><br>V1/V2 loop | Phase 1/2 study on safety, pharamacokinetics, and dose finding | Newborns with HIV exposure | Enrolling |
| SAMBULEL O | VRC07-523LS | CD4-binding site | Phase 2 study on safety and pharmacokinetics | Newborns with HIV exposure, without HIV exposure, and with HIV | In development |
| EDCTP Neo bNAb Trial | VRC07-523LS | CD4-binding site | Proof-of-concept on effectiveness, efficacy, and operational feasibility | Newborns with HIV exposure | In development |
| IMPAACT 2037 | PGT121.414.LS *with* or *without* VRC07523-LS | V3 glycan<br><br>CD4-binding site | Phase 1 study on safety and pharmacokinetics | Newborns with HIV exposure | In development |

Table adapted from [19].

Since there are currently no published human infant HIV prophylaxis efficacy studies, base case efficacy was estimated using data from human adult, non-human primate, and *in vitro* studies. In the Antibody Mediated Prevention (AMP) trials, VRC01 did not prevent overall sexual acquisition of HIV among populations of cisgender men and transgender adults in the Americas and Europe, or among women in sub-Saharan Africa.[20] However, acquisition of VRC01-sensitive isolates was 75·4% lower among individuals who received VRC01 compared to individuals who received placebo ($IC_{80}$ <1 μg/mL).[20] Other bNAbs under investigation, such as 3BNC117-LS, VRC07-523LS, 1-18, and 10-1074-LS are more potent and have greater breadth than VRC01.[21] For example, VRC07-523LS neutralized 91% of primary African isolates in a multiclade panel with an $IC_{80}$ of 4·4 μg/mL compared to VRC01 neutralizing 66% with an $IC_{80}$ of 18·4 μg/mL.[21]

By applying the 75% efficacy against acquisition of sensitive isolates found in the AMP trial to the 91% neutralization coverage of VRC07-523LS found in the multiclade panel, we estimate VRC07-523LS could potentially have 68% overall efficacy. Similar to combination ART, a final bNAb infant prophylaxis product will likely be a combination of bNAbs, such as the VRC07-523LS+PGT121+PGDM1400 combination currently being studied as adult prophylaxis (NCT03205917, NCT03928821, NCT04212091, and NCT03721510), to achieve high breadth and potency. As such, we assumed a three-bNAb combination would have 70% efficacy in the base case, only slightly higher than the implied efficacy of VRC07-523LS alone to be conservative of the clinical impact a bNAb combination could have as prophylaxis. Given the uncertainty in final product characteristics, bNAb efficacy was varied widely in sensitivity analyses.

While the AMP studies investigated bNAb efficacy when delivered as pre-exposure prophylaxis, non-human primate studies have also observed efficacy of bNAbs as post-exposure prophylaxis if given within 24-30 hours of exposure.[22,23] Based on these data, we assumed that a bNAb given at birth would reduce the risk of IP HIV transmission as post-exposure prophylaxis, in addition to acting as pre-exposure prophylaxis for postnatal transmission.

Pharmacokinetic data from HIV-exposed infants suggest that with an 80mg dose of VRC07-523LS delivered subcutaneously at birth, concentrations of the bNAb would remain sufficiently high to achieve protective efficacy through at least 12 weeks of life.[16] In the base case, we assumed a three-month effect duration following each bNAb dose, and we also varied this assumption in sensitivity analyses.

##### BNAb cost estimates

The base case average bNAb cost was modeled as $20 per dose, including the estimated costs of production (assuming a 100mg bNAb dose), as well as delivery (including training, personal protective equipment, and cold-chain), personnel, and facility/overhead needed to administer vaccines in low- and middle-income countries. The midpoint of the estimated cost range for all components was rounded to the nearest dollar and summed to reflect the total cost of bNAbs if they were produced, delivered, and administered at scale.

**Itemized costs included in modeled bNAb cost per dose**

| Cost component | Cost estimate or range* | Rounded midpoint cost* |
|---|---|---|
| BNAb production costs per 100mg dose[24] | $2·00 - $20·00 | $11 |
| Supply and delivery costs per dose, including: training, social mobilization, hand hygiene, personal protective equipment, waste management, and cold-chain costs[25] | $1·66 | $2 |
| Service personnel costs[26] | $0·68 - $1·45 | $1 |
| Facility overhead and capital costs[27] | $1·92 - $4·21 | $3 |
| **Total estimated cost** | **$6·26 - $27·32** | **$17** |

*All costs are reported in 2020 USD.

To conservatively estimate the cost-effectiveness of bNAb infant prophylaxis, the total estimated cost was rounded up to the nearest ten ($20/dose). The bNAb cost was varied widely in sensitivity analyses ($5/dose - $100/dose) to capture the total estimated cost using the lower bound of each cost component and to account for scenarios in which there are additional unforeseen costs or if bNAbs are not produced at scale.

##### Determining cost-effectiveness

We calculated incremental cost-effectiveness ratios (ICERs; USD/year of life saved [YLS]) by ordering strategies by increasing discounted life expectancy and dividing the difference in discounted costs by the difference in discounted life expectancy of consecutive, non-dominated strategies. Life expectancy and costs were discounted at 3%/year per standard cost-effectiveness practices.[28] BNAb strategies were dominated if they resulted in shorter life expectancy than a less costly strategy or resulted in higher costs per YLS (i.e., higher ICER) than the next best-performing, non-dominated strategy. We considered a bNAb strategy cost-saving if it was non-dominated and resulted in the same or greater life expectancy than the standard-of-care at lower lifetime HIV-related costs.

While the WHO-CHOICE 100% GDP per capita-based cost-effectiveness threshold has been widely cited, there is growing concern that spending to this threshold may not offer good value, particularly in resource-limited settings.[29–35] Due to budget constraints, investing in interventions at this GDP-based cost-effectiveness threshold may result in substantial opportunity costs (e.g., forgone health benefits) relative to other investments in health care that offer better value.[29–35] Cost-effectiveness thresholds accounting for these additional considerations have been estimated in the published literature:

**Cost-effectiveness thresholds for LMICs proposed in published literature**

| | **Cost-effectiveness threshold (in % GDP per capita)** | | |
|---|---|---|---|
| **Source** | **Côte d'Ivoire** | **South Africa** | **Zimbabwe** |
| Woods B, Revill P, Sculpher M, Claxton K, 2016[29] | 4-52% | 17-69% | 1-32% |
| Ochalek JM, Lomas J, Claxton KP, 2018[33] | 15-19% | 43-58% | 22-30% |
| Jit M, 2021[34] | 30-40% | 30-40% | 30-40% |
| Edoka EP, Stacey NK, 2020[35] | - | 53% | - |
| Meyer-Rath G, Rensburg C, Larson B, Jamieson L, Rosen S, 2017[31] | - | 10-17% | - |

Table adapted from [36].

As such, we used a more conservative cost-effectiveness threshold of 50% of GDP per capita in our base case analysis. Strategies that resulted in the greatest clinical benefit with an ICER $\leq$50% of a country's GDP per capita (Côte d'Ivoire: $1163/YLS, South Africa: $2828/YLS, Zimbabwe: $607/YLS)[37] were considered cost-effective. Given that a 50% GDP per capita cost-effectiveness threshold may still require donor-support, we also assessed the impact of using a 20% GDP per capita cost-effectiveness threshold in a sensitivity analysis. Since the cost-effective bNAb strategy did not change using the 20% GDP per capita cost-effectiveness threshold in any of the three settings modeled under base case bNAb cost and efficacy assumptions, we chose to only present the 50% GDP per capita cost-effectiveness threshold in the main manuscript.

**Supplementary Figure 1.** CEPAC-P infant prophylaxis module flowchart

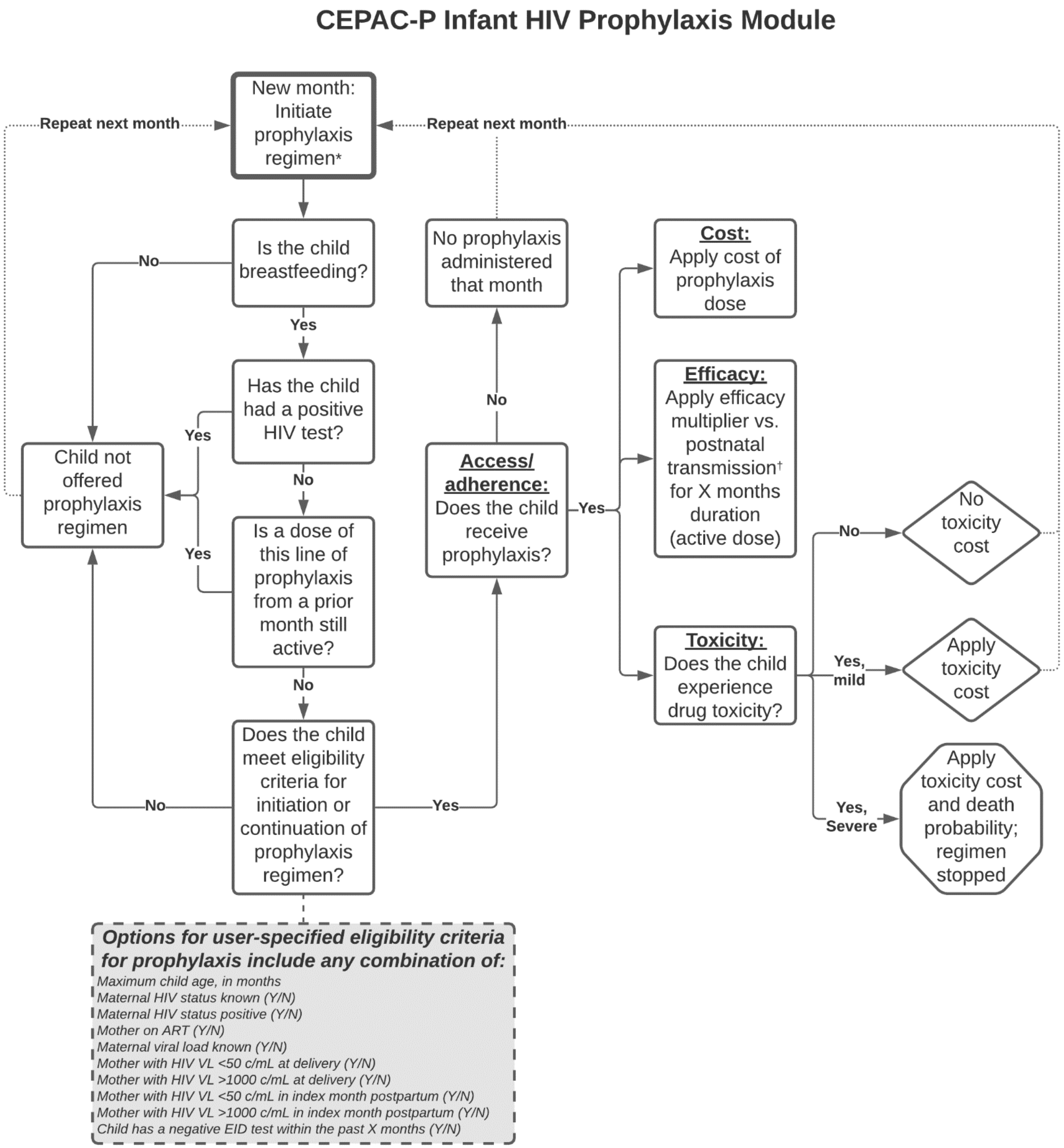

CEPAC-P, Cost-Effectiveness of Preventing AIDS Complications–Pediatrics Model; Y, yes; N, no; ART, antiretroviral therapy; VL, viral load; EID, early infant diagnosis.

* This process is repeated separately for each line of prophylaxis each month.

† Efficacy of prophylaxis against intrapartum transmission is applied directly as a one-time multiplier against the intrapartum component of the intrauterine/intrapartum transmission risk.

**Supplementary Figure 2.** Schematic of modeled cohorts and bNAb strategies

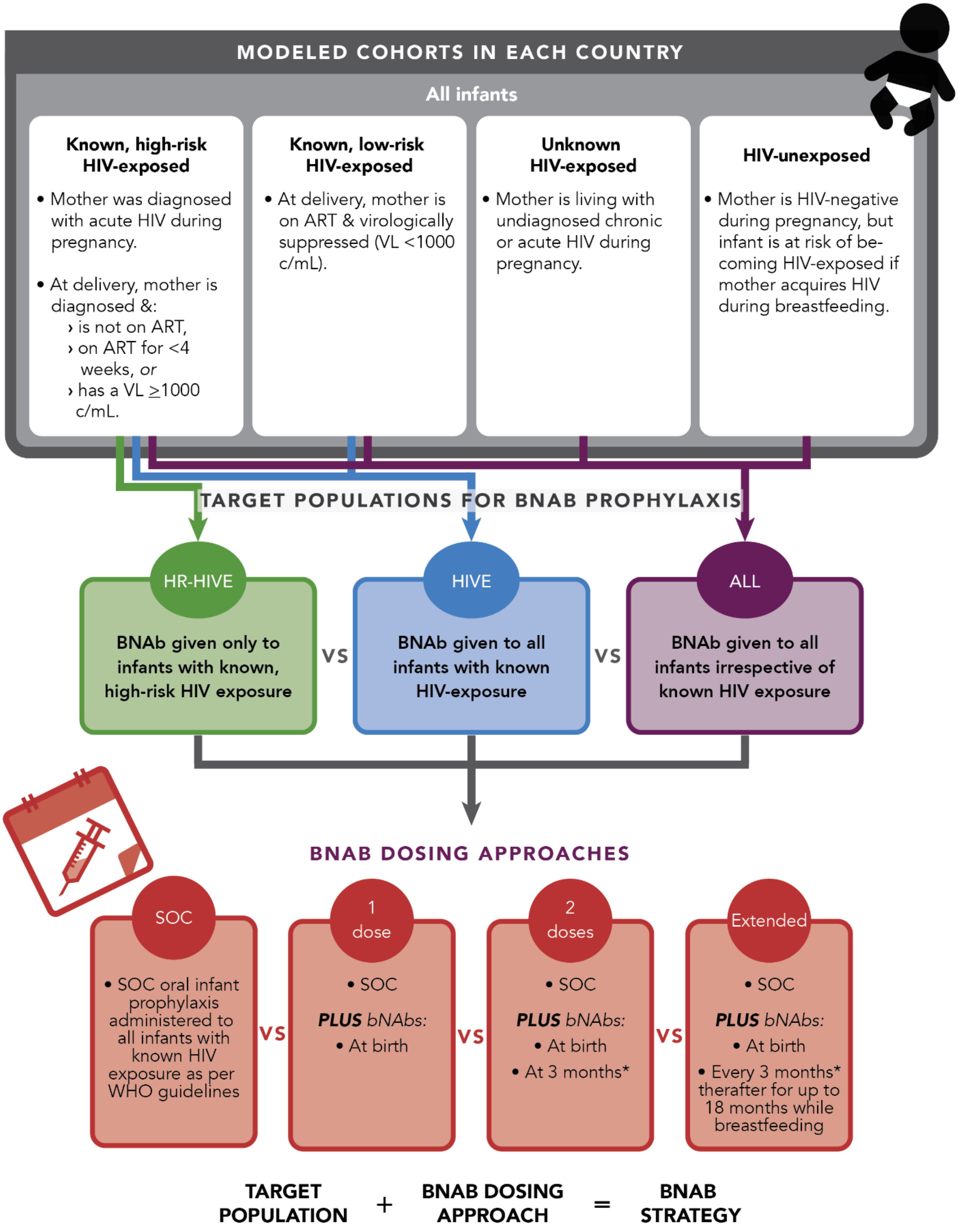

ART, antiretroviral therapy; VL, viral load; SOC, standard-of-care.

* The bNAb effect duration was varied during sensitivity analysis, and the frequency of administration was adjusted accordingly.

**Supplementary Figure 3.** Efficiency frontier of all bNAb strategies assessed, by country

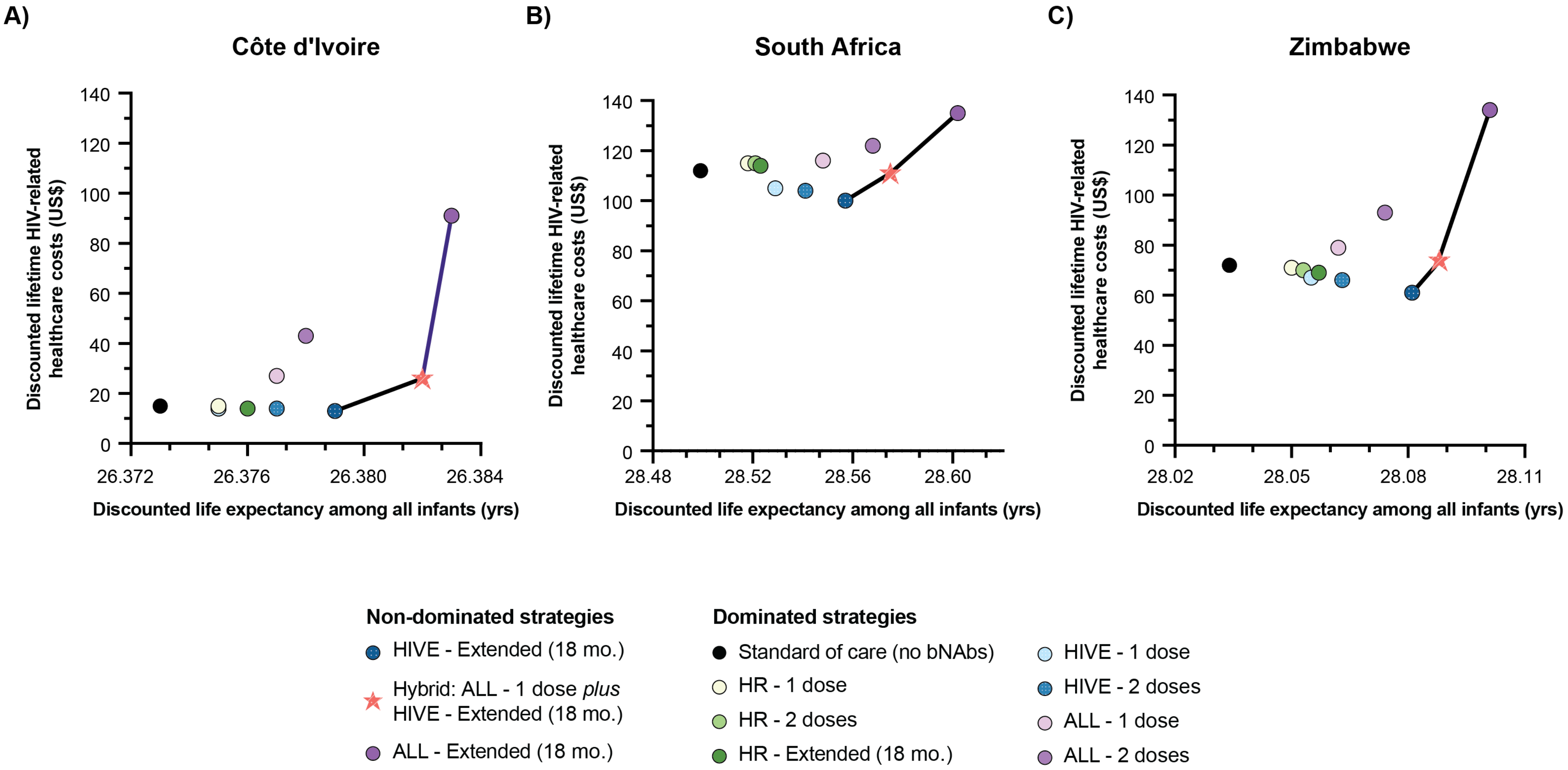

US$, United States dollar; yrs, years; HIVE, bNAb strategy offering bNAbs to infants with known HIV exposure at birth; ALL, bNAb strategy offering bNAbs to all infants regardless of HIV exposure; mo., month.

**Supplementary Figure 4.** Cumulative vertical transmission in standard-of-care and bNAb strategies, by country

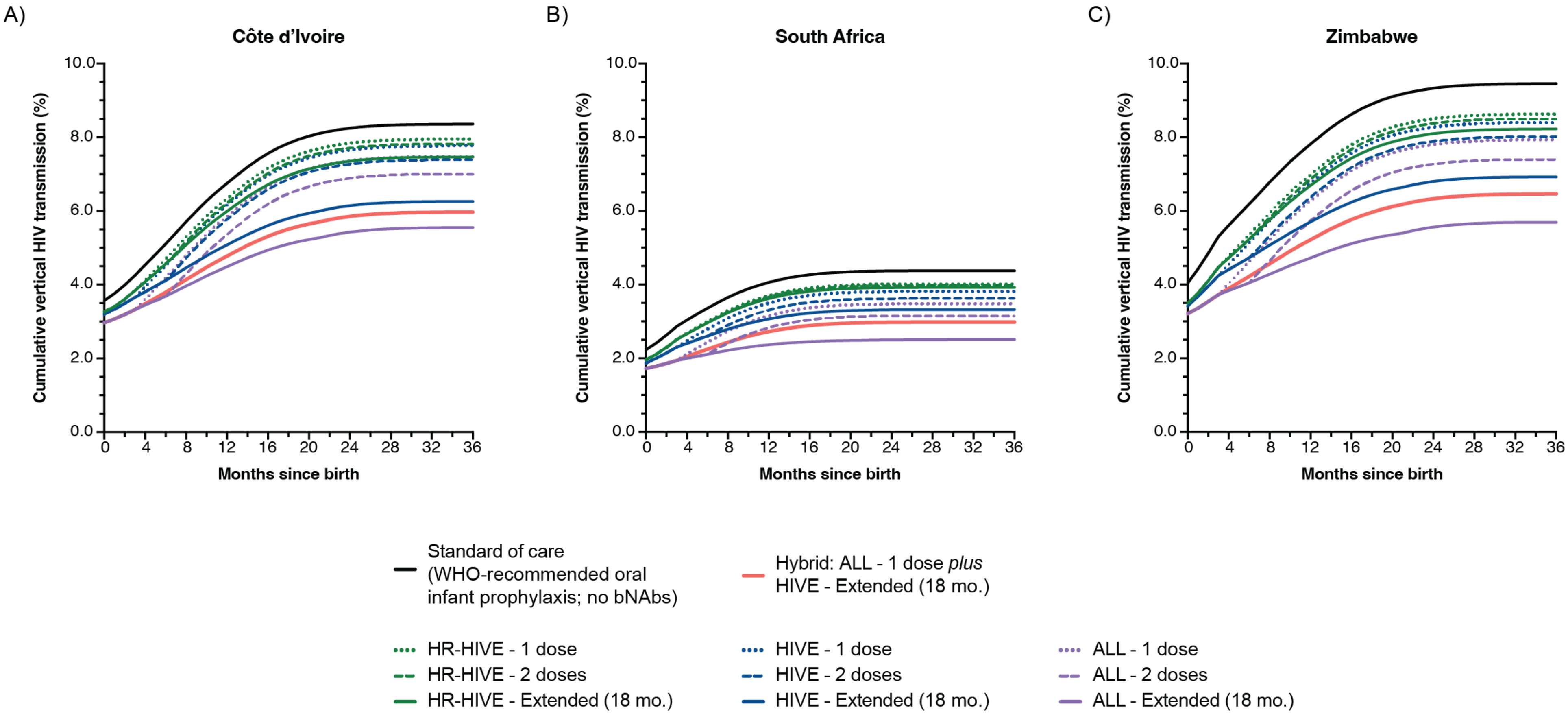

BNAb, broadly neutralizing antibody; WHO, World Health Organization; HR-HIVE, bNAb strategy targeting infants with known, high-risk HIV exposure at birth; HIVE, bNAb strategy targeting infants with known HIV exposure at birth; ALL, bNAb strategy targeting all infants regardless of known HIV exposure; mo., month.

**Supplementary Figure 5.** Breakdown of intrauterine/intrapartum infections occurring in each exposure group

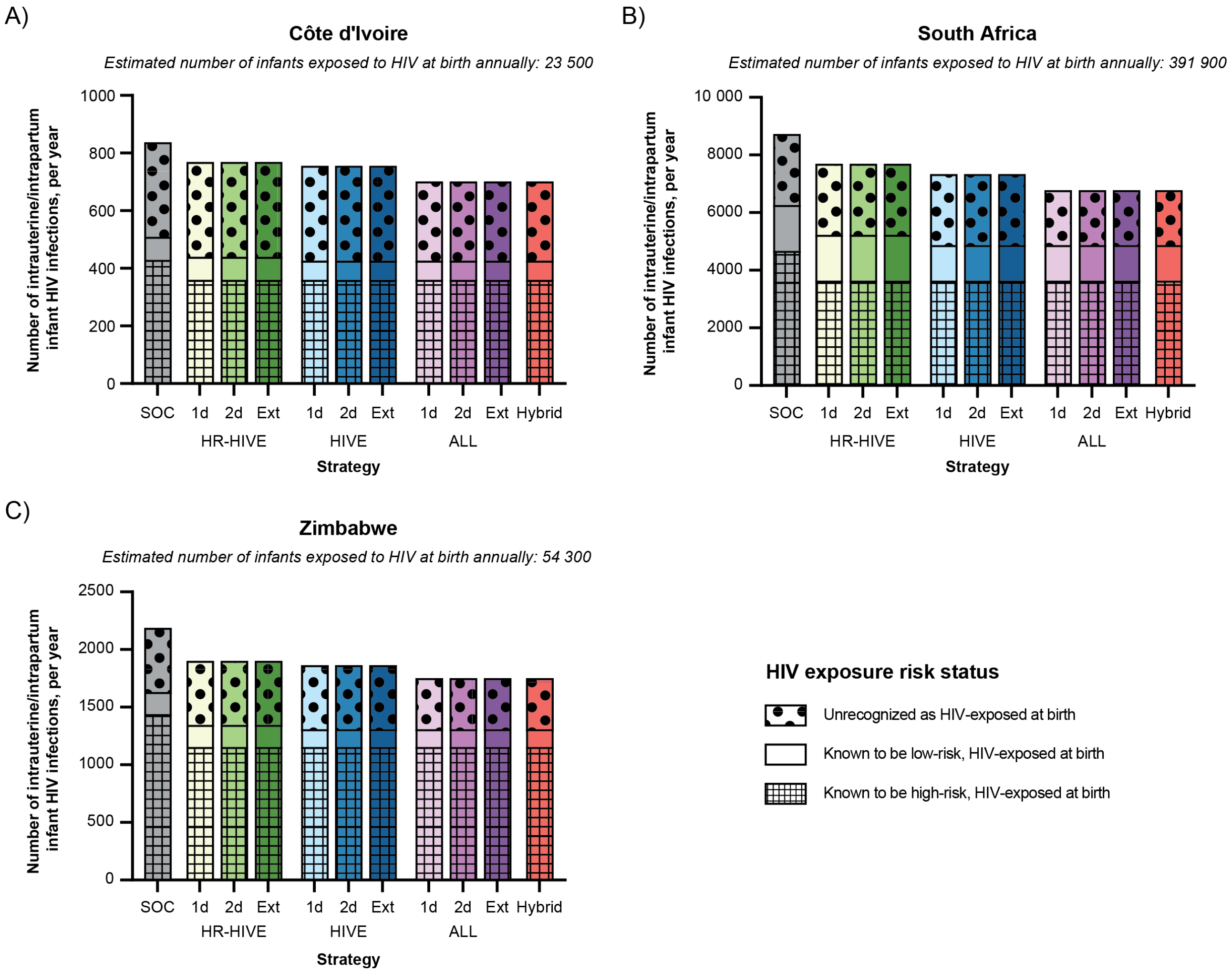

SOC, standard-of-care; 1d, one bNAb dose; 2d, two bNAb doses; Ext, extended bNAb dosing (one dose every three months through 18 months of life while breastfeeding); HR-HIVE, bNAb strategy targeting infants with known, high-risk HIV exposure at birth; HIVE, bNAb strategy targeting infants with known HIV exposure at birth; ALL, bNAb strategy targeting all infants regardless of known HIV exposure.

The estimated number of infants at risk of intrauterine/intrapartum HIV acquisition in each exposure group varies by country. In Côte d'Ivoire, an estimated 6900 infants have known, high-risk HIV exposure at birth; 14 800 infants have known, low-risk HIV exposure at birth; and 1400 infants have unrecognized HIV exposure at birth. In South Africa, an estimated 66 200 infants have known, high-risk HIV exposure at birth; 295 700 have known, low-risk HIV exposure at birth; and 11 600 infants have unrecognized HIV exposure at birth. In Zimbabwe, an estimated 12 600 infants have known, high-risk HIV exposure at birth; 35 500 infants have known, low-risk HIV exposure at birth; and 2600 infants have unrecognized HIV exposure at birth.

**Supplementary Figure 6.** Cost-effective bNAb strategy across influential univariate sensitivity analyses, by country

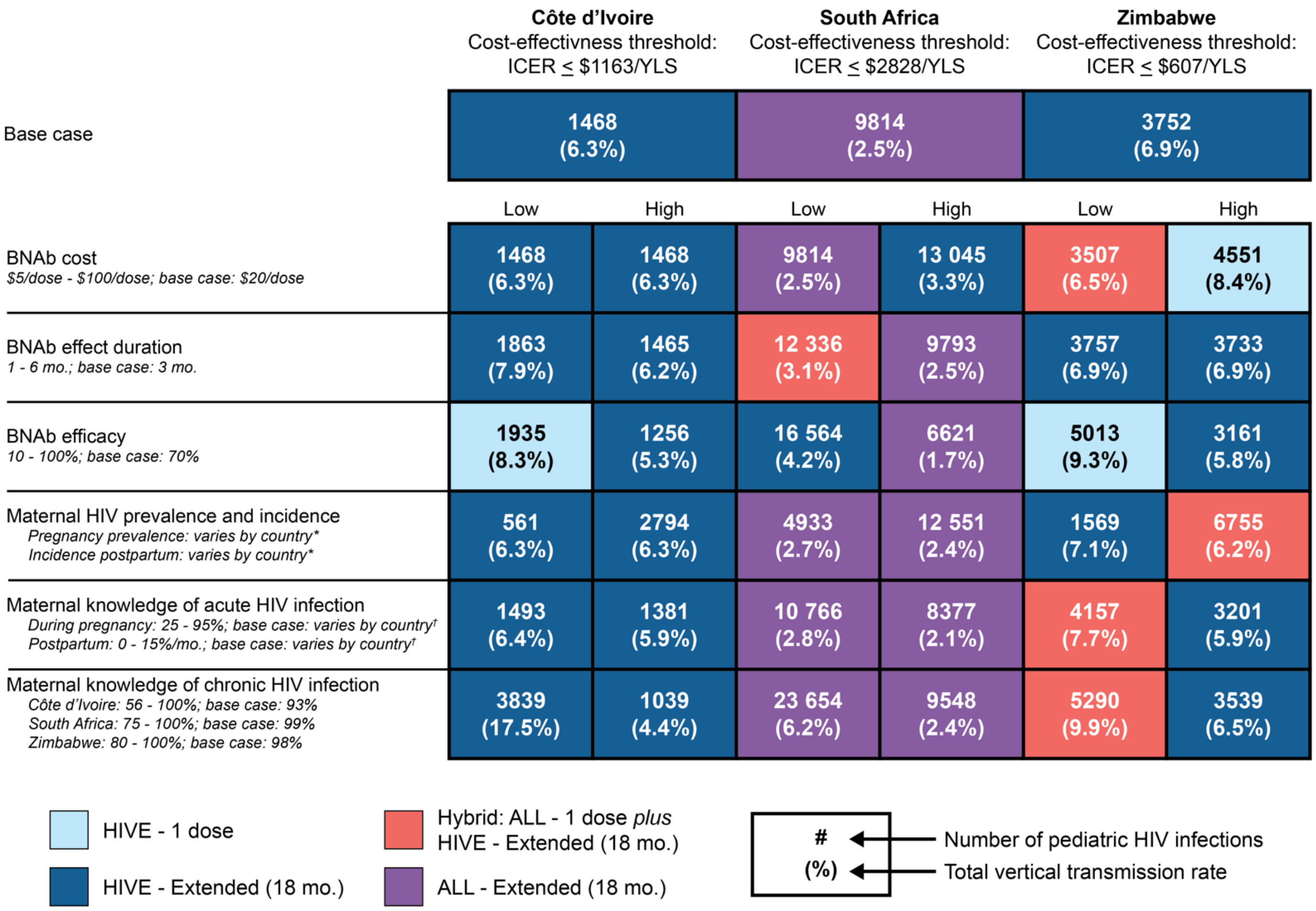

ICER, incremental cost-effectiveness ratio; YLS, year of life saved; bNAbs, broadly neutralizing antibodies; mo., month; HIVE, bNAb strategy targeting infants with known HIV exposure at birth; ALL, bNAb strategy targeting all infants regardless of known HIV exposure.

* Maternal HIV prevalence at the time of delivery is inclusive of chronic HIV infection and incident infection during pregnancy. Maternal HIV prevalence was varied according to the lowest and highest subnational estimates in each country (Côte d'Ivoire: 1-5%, base case: 3%; South Africa: 15-45%, base case: 33%; and Zimbabwe: 5-25%, base case: 12%). Postpartum maternal HIV incidence was also decreased (Côte d'Ivoire: 0.001%/month, South Africa: 0.107%/month, Zimbabwe: 0.022%/month) or increased (Côte d'Ivoire: 0.005%/month, South Africa: 0.331%/month, Zimbabwe: 0.116%/month) proportional to the change in prevalence.

† In the base case, maternal knowledge of acute HIV infection in Côte d'Ivoire is 56% for infection acquired during pregnancy and 2% per month postpartum, in South Africa is 55% during pregnancy and 9% per month postpartum, and in Zimbabwe is 70% during pregnancy and 5% per month postpartum.

**Supplementary Figure 7.** Cost-effective bNAb strategy across non-influential univariate sensitivity analyses, by country

| | **Côte d'Ivoire** Cost-effectivness threshold: ICER ≤ $1163/YLS | **South Africa** Cost-effectiveness threshold: ICER ≤ $2828/YLS | **Zimbabwe** Cost-effectiveness threshold: ICER ≤ $607/YLS |
|---|---|---|---|
| Base case | 1468 (6.3%) | 9814 (2.5%) | 3752 (6.9%) |
| BNAbs have no intrapartum efficacy | 1549 (6.6%) | 11 728 (3.0%) | 4070 (7.6%) |
| Infants receive either bNAbs or standard of care oral infant prophylaxis, but not both | 1518 (6.5%) | 10 213 (2.6%) | 3928 (7.2%) |

| | Côte d'Ivoire Low | Côte d'Ivoire High | South Africa Low | South Africa High | Zimbabwe Low | Zimbabwe High |
|---|---|---|---|---|---|---|
| ART costs<br>*0.5 - 2.0x; base case: 1.0x* | 1468 (6.3%) | 1468 (6.3%) | 9814 (2.5%) | 9814 (2.5%) | 3752 (6.9%) | 3752 (6.9%) |
| BNAb uptake*<br>*Côte d'Ivoire: 56 - 83%; base case: varies by age*<br>*South Africa: 85 - 96%; base case: varies by age*<br>*Zimbabwe: 71 - 92%; base case: varies by age* | 1578 (6.7%) | 1392 (5.9%) | 10 461 (2.7%) | 9655 (2.5%) | 3978 (7.3%) | 3634 (6.7%) |
| Breastfeeding duration<br>*Duration among women with/without HIV:*<br>*Côte d'Ivoire: 8/8 - 24/24 mo.; base case: 14/19 mo.*<br>*South Africa: 2/2 - 18/18 mo.; base case: 6/12 mo.*<br>*Zimbabwe: 8/15 - 20/20 mo.; base case: 13/18 mo.* | 1114 (4.8%) | 1974 (8.4%) | 7957 (2.1%) | 14 640 (3.7%) | 3265 (6.1%) | 4428 (8.1%) |
| Cost of ascertaining high-risk status<br>*$0 - $31.66; base case: $31.66* | 1468 (6.3%) | 1468 (6.3%) | 9814 (2.5%) | 9814 (2.5%) | 3752 (6.9%) | 3752 (6.9%) |
| Maternal ART coverage during pregnancy†<br>*Côte d'Ivoire: 70 - 100%; base case: 95%*<br>*South Africa: 60 - 100%; base case: 97%*<br>*Zimbabwe: 65 - 100%; base case: 89%* | 2140 (9.4%) | 1346 (5.7%) | 25 351 (6.7%) | 8752 (2.2%) | 5078 (9.6%) | 3112 (5.7%) |
| Oral infant prophylaxis efficacy<br>*Intrapartum efficacy: 40 - 90%; base case: 69%*<br>*Postnatal efficacy: 58 - 90%; base case: 71%* | 1491 (6.4%) | 1452 (6.2%) | 10 002 (2.6%) | 9681 (2.5%) | 3860 (7.1%) | 3676 (6.8%) |
| Postnatal vertical transmission risk<br>*0.5 - 2.0x; base case: 1.0x* | 1119 (4.8%) | 2129 (9.1%) | 8299 (2.1%) | 12 770 (3.3%) | 2831 (5.2%) | 5431 (10.0%) |
| Postpartum maternal ART retention<br>*50% - 100% at 6+ mo. among mothers with known status; base case: varies by month and country* | 1729 (7.4%) | 1359 (5.8%) | 10 658 (2.7%) | 9 528 (2.4%) | 4203 (7.8%) | 3574 (6.6%) |
| Postpartum maternal HIV incidence<br>*0.5 - 2.0x; base case: 1.0x* | 1433 (6.2%) | 1544 (6.5%) | 9293 (2.4%) | 10 830 (2.6%) | 3435 (6.6%) | 4576 (7.9%) |

HIVE - 1 dose
HIVE - Extended (18 mo.)
Hybrid: ALL - 1 dose *plus* HIVE - Extended (18 mo.)
ALL - Extended (18 mo.)

\# → Number of pediatric HIV infections
(%) → Total vertical transmission rate

ICER, incremental cost-effectiveness ratio; YLS, year of life saved; bNAbs, broadly neutralizing antibodies; ART, antiretroviral therapy; mo., month; HIVE, bNAb strategy targeting infants with known HIV exposure at birth; ALL, bNAb strategy targeting all infants regardless of known HIV exposure.

* In the base case, bNAb uptake varies by age informed by the country-specific percentage of infants delivered in a healthcare facility and uptake of vaccines in the routine infant vaccination schedule. In sensitivity analyses, the lowest and highest of each country's age-specific uptake was used as the uptake at every timepoint for the low and high scenarios, respectively.

† Maternal ART coverage during pregnancy is a proportion of all women known to have HIV during pregnancy.

**Supplementary Figure 8.** Cost-effectiveness results of a three-way sensitivity analysis assessing bNAb efficacy, cost, and cost-effectiveness threshold, by country

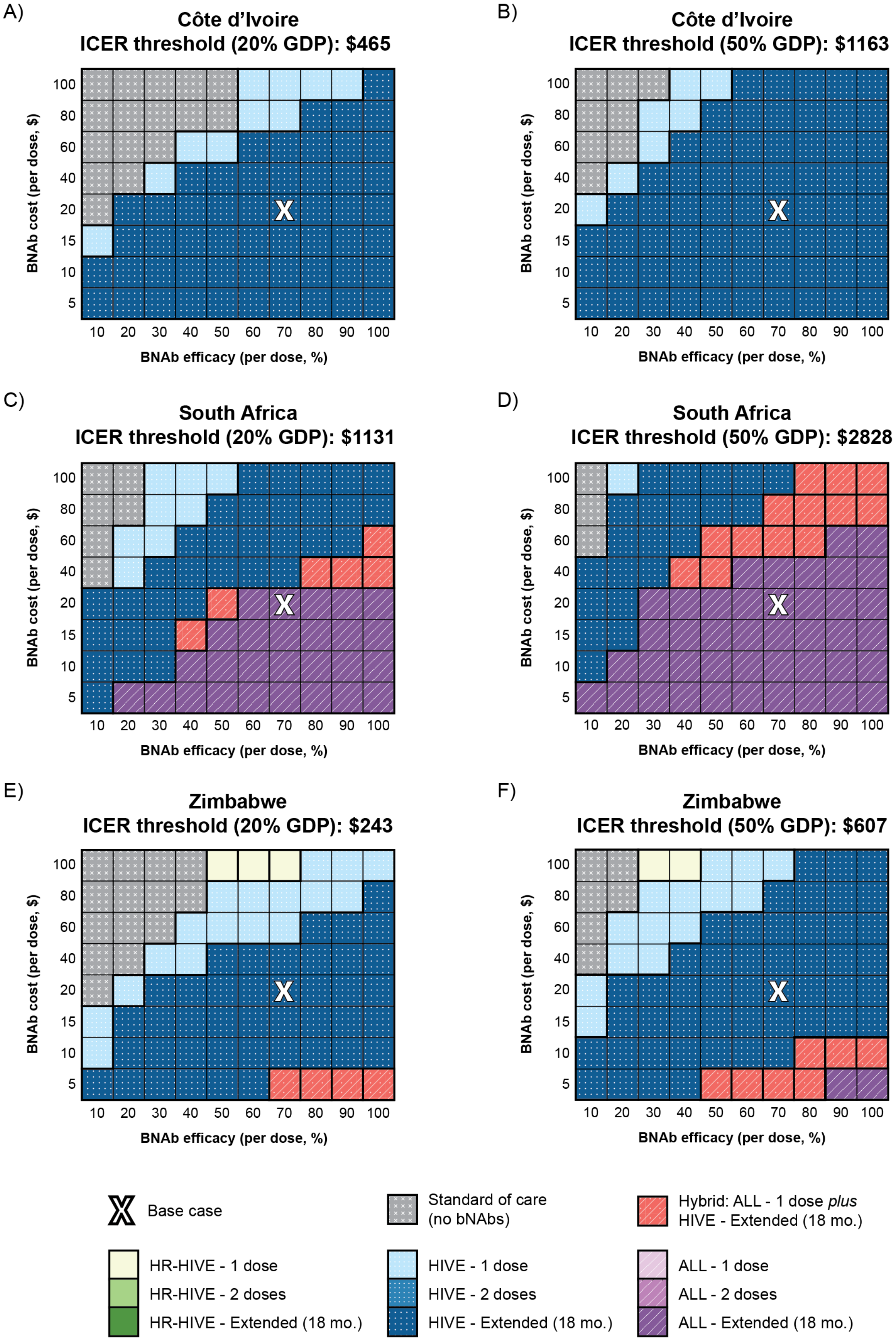

ICER, incremental cost-effectiveness ratio; GDP, gross domestic product; bNAb, broadly neutralizing antibody; HR-HIVE, bNAb strategy targeting infants with known, high-risk HIV exposure at birth; HIVE, bNAb strategy targeting infants with known HIV exposure at birth; ALL, bNAb strategy targeting all infants regardless of known HIV exposure; mo., month.

**Supplementary Figure 9.** Cost-effectiveness results of a three-way sensitivity analysis assessing bNAb efficacy, cost, and efficacy duration, by country

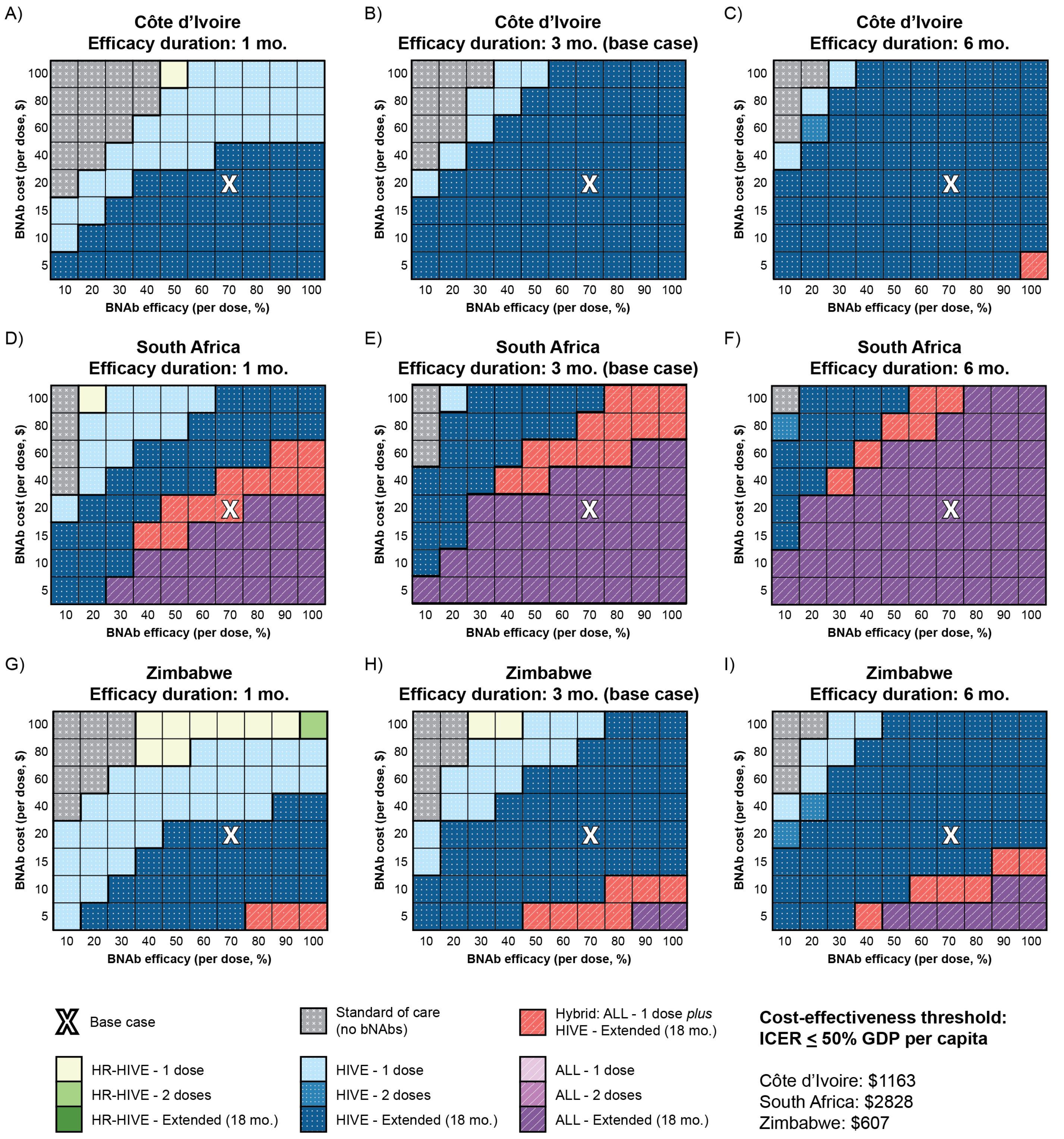

mo., month; bNAb, broadly neutralizing antibody; HR-HIVE, bNAb strategy targeting infants with known, high-risk HIV exposure at birth; HIVE, bNAb strategy targeting infants with known HIV exposure at birth; ALL, bNAb strategy targeting all infants regardless of known HIV exposure; ICER, incremental cost-effectiveness ratio; GDP, gross domestic product.

**Supplementary Table 1.** Consolidated Health Economic Evaluation Reporting Standards (CHEERS) checklist

| Topic | No. | Item | Location where item is reported |
|---|---|---|---|
| **Title** | | | |
| Title | 1 | Identify the study as an economic evaluation and specify the interventions being compared. | Title, Page 1 |
| **Abstract** | | | |
| Abstract | 2 | Provide a structured summary that highlights context, key methods, results, and alternative analyses. | Abstract, Pages 4-5 |
| **Introduction** | | | |
| Background and objectives | 3 | Give the context for the study, the study question, and its practical relevance for decision making in policy or practice. | Introduction, Pages 6-7 |
| **Methods** | | | |
| Health economic analysis plan | 4 | Indicate whether a health economic analysis plan was developed and where available. | n/a |
| Study population | 5 | Describe characteristics of the study population (such as age range, demographics, socioeconomic, or clinical characteristics). | Methods, Third Paragraph |
| Setting and location | 6 | Provide relevant contextual information that may influence findings. | Methods, First Paragraph |
| Comparators | 7 | Describe the interventions or strategies being compared and why chosen. | Methods, Fourth Paragraph |
| Perspective | 8 | State the perspective(s) adopted by the study and why chosen. | Methods, Tenth Paragraph |
| Time horizon | 9 | State the time horizon for the study and why appropriate. | Methods, Tenth Paragraph |
| Discount rate | 10 | Report the discount rate(s) and reason chosen. | Methods, Tenth Paragraph |
| Selection of outcomes | 11 | Describe what outcomes were used as the measure(s) of benefit(s) and harm(s). | Methods, Tenth Paragraph |
| Measurement of outcomes | 12 | Describe how outcomes used to capture benefit(s) and harm(s) were measured. | n/a |
| Valuation of outcomes | 13 | Describe the population and methods used to measure and value outcomes. | n/a |
| Measurement and valuation of resources and costs | 14 | Describe how costs were valued. | Methods, Sixth and Ninth Paragraphs |
| Currency, price date, and conversion | 15 | Report the dates of the estimated resource quantities and unit costs, plus the currency and year of conversion. | Methods, Ninth Paragraph |
| Rationale and description of model | 16 | If modelling is used, describe in detail and why used. Report if the model is publicly available and where it can be accessed. | Methods, First paragraph; Appendix Supplementary Methods; Supplementary Figure 1 |
| Analytics and assumptions | 17 | Describe any methods for analysing or statistically transforming data, any extrapolation methods, and approaches for validating any model used. | Methods, Second Paragraph; Appendix Supplemental Methods; Supplementary Table 2 |
| Characterising heterogeneity | 18 | Describe any methods used for estimating how the results of the study vary for subgroups. | n/a |
| Characterising distributional effects | 19 | Describe how impacts are distributed across different individuals or adjustments made to reflect priority populations. | Methods , Third Paragraph |
| Characterising uncertainty | 20 | Describe methods to characterise any sources of uncertainty in the analysis. | Methods, Last Paragraph |
| Approach to engagement with patients and others affected by the study | 21 | Describe any approaches to engage patients or service recipients, the general public, communities, or stakeholders (such as clinicians or payers) in the design of the study. | n/a |
| **Results** | | | |
| Study parameters | 22 | Report all analytic inputs (such as values, ranges, references) including uncertainty or distributional assumptions. | Table 1; Supplementary Table 2 |
| Summary of main results | 23 | Report the mean values for the main categories of costs and outcomes of interest and summarise them in the most appropriate overall measure. | Results, First and Second Paragraphs; Table 2 |
| Effect of uncertainty | 24 | Describe how uncertainty about analytic judgments, inputs, or projections affect findings. Report the effect of choice of discount rate and time horizon, if applicable. | Results, Third through Eighth Paragraphs; Figures 4-5; Supplementary Figures 5-7; Supplementary Tables 6-22 |
| Effect of engagement with patients and others affected by the study | 25 | Report on any difference patient/service recipient, general public, community, or stakeholder involvement made to the approach or findings of the study | n/a |
| **Discussion** | | | |
| Study findings, limitations, generalisability, and current knowledge | 26 | Report key findings, limitations, ethical or equity considerations not captured, and how these could affect patients, policy, or practice. | Discussion, First through Ninth Paragraphs |
| **Other relevant information** | | | |
| Source of funding | 27 | Describe how the study was funded and any role of the funder in the identification, design, conduct, and reporting of the analysis | Title, Pages 2-3 |
| Conflicts of interest | 28 | Report authors conflicts of interest according to journal or International Committee of Medical Journal Editors requirements. | Title, Page 3 |

**Supplementary Table 2.** Extended model input parameters

| Parameter | Côte d'Ivoire | South Africa | Zimbabwe | Reference(s) |
|---|---|---|---|---|
| **Maternal characteristics** | | | | |
| Proportion of mothers with chronic infection, % | | | | |
| High-risk cohort | 98 | 79 | 62 | Derived from [38–55] |
| Low-risk cohort | 100 | 100 | 100 | Assumption |
| Unrecognized exposed cohort | 94 | 12 | 27 | Derived from [38–50,53–56] |
| Unexposed cohort | 0 | 0 | 0 | Assumption |
| Probability of acute HIV infection in pregnancy, % | 0·03 | 3·30 | 1·94 | [55,45–48] |
| Probability of acute HIV infection postnatally, %/mo. | 0·003 | 0·240 | 0·056 | [52,55,57–59] |
| Probability of known HIV status in pregnancy, % | | | | Derived from [38–41,49,52,56,60,61] |
| Among mothers with chronic infection | 93 | 99 | 98 | [42,43,49,53,55,62] |
| Among mothers with acute infection | 56 | 55 | 70 | [38–44,56] |
| Probability of learning HIV status postnatally, %/mo. | 2 | 9 | 5 | [43,53,55] |
| Probability of being on ART during pregnancy, % | | | | |
| High-risk cohort | | | | |
| Among those with chronic infection | 86 | 84 | 41 | Derived from [42,43,49–55] |
| Among those with acute infection | 95 | 97 | 89 | Derived from [38–50,53–56,63] |
| Low-risk cohort | 100 | 100 | 100 | Assumption |
| Unrecognized exposed and unexposed cohorts | 0 | 0 | 0 | Assumption |
| Viral load if on ART at delivery, % | | | | |
| Non-high-risk cohort | | | | Derived from [42,43,49–53,60,64] |
| HIV RNA <u><</u>50 c/mL | 76 | 76 | 76 | |
| HIV RNA 50-1,000 c/mL | 24 | 24 | 24 | |
| HIV RNA >1,000 c/mL | 0 | 0 | 0 | |
| High-risk and unrecognized exposed cohorts | | | | Assumption |
| HIV RNA >1,000 c/mL | 100 | 100 | 100 | |
| Probability of delivering in a healthcare facility, % | 70 | 96 | 86 | [42–44] |
| HIV RNA <1,000 c/mL among postpartum women on ART, by month postpartum | 77-92 | 77-92 | 77-92 | [65–74] |
| Retention in HIV care, % | | | | [75] |
| High-risk and low-risk cohorts | | | | |
| 6 mo. postpartum | 87 | 88 | 88 | |
| 12 mo. postpartum | 87 | 87 | 87 | |
| 24 mo. postpartum | 79 | 79 | 79 | |
| Unrecognized exposed and unexposed cohorts | | | | |
| 6 mo. postpartum | 9 | 37 | 19 | |
| 12 mo. postpartum | 17 | 57 | 33 | |
| 24 mo. postpartum | 27 | 69 | 47 | |
| **Infant cohort characteristics and transmission risks** | | | | |
| Total number of live births (thousands) | 907 | 1153 | 425 | [76] |
| Age, mean (SD), mo. | 0 (0) | 0 (0) | 0 (0) | Assumption |
| Proportion female, % | 52 | 52 | 52 | [77] |
| Initial CD4% at infection, mean (SD), % | 45 (9) | 45 (9) | 45 (9) | [1] |
| Breastfed infants, % | | | | [43,53,55,78] |
| Unexposed or Unrecognized HIV-exposed infants | 99 | 84 | 99 | |
| Infants with known HIV exposure | 99 | 66 | 94 | |
| Breastfeeding duration, mean (SD) mo. | | | | [42–44,68,79] |
| Unexposed or Unrecognized HIV-exposed infants | 19 (7) | 12 (6) | 18 (7) | |
| Infants with known HIV exposure | 14 (7) | 6 (6) | 13 (7) | |
| Intrauterine infections as a share of all perinatal infections, % | 67 | 67 | 67 | [80] |
| Perinatal transmission, one-time % | | | | |
| Chronic maternal HIV infection in pregnancy | | | | |
| On ART, HIV RNA <u><</u>50 c/mL at delivery | 0·24 | 0·24 | 0·24 | [81] |
| On ART, HIV RNA 50-1,000 c/mL at delivery | 1·45 | 1·45 | 1·45 | [81] |

| Parameter | Côte d'Ivoire | South Africa | Zimbabwe | Reference(s) |
|---|---|---|---|---|
| On ART, HIV RNA >1,000 c/mL at delivery | 4·14 | 4·14 | 4·14 | [81] |
| Not on ART at delivery | 19·70 | 19·70 | 19·70 | [82] |
| Acute maternal HIV infection in pregnancy | | | | |
| On ART at delivery | 8·79 | 8·79 | 8·79 | [81,82] |
| Not on ART at delivery | 18·10 | 18·10 | 18·10 | [82] |
| Postnatal transmission rate, %/mo.[†, ϕ] | | | | |
| On ART, HIV RNA <u><</u>50 c/mL | 0·06 | 0·06 | 0·06 | [81] |
| On ART, HIV RNA 50-1,000 c/mL | 0·39 | 0·39 | 0·39 | [81] |
| On ART, HIV RNA >1,000 c/mL | 0·78 | 0·78 | 0·78 | [81] |
| Not on ART | 0·89 | 0·89 | 0·89 | [82] |
| **Early infant diagnosis** | | | | |
| Probability of uptake, % by age | | | | |
| Birth | 0 | 93 | 9 | [83,84] |
| 6-8 weeks | 69 | 64 | 54 | [53,61,68,85–89] |
| 6/9 months (country-specific timepoint) | 25 | 25 | 26 | [53,90] |
| 18 months | 22 | 22 | 22 | [91] |
| Nucleic acid amplification test characteristics | | | | |
| Sensitivity, 0 months after infection, % | 0 | 0 | 0 | Assumption |
| Sensitivity, ≥1 month after infection, % | 99·4 | 99·4 | 99·4 | [92] |
| Specificity, % | 99·6 | 99·6 | 99·6 | [92] |
| Probability of result return, % | 84 | 96 | 83 | [61,93,94] |
| Delay between primary test and result return, mean mo. (SD) | 2 (1) | 2 (1) | 2 (1) | [61,93] |
| Probability of linkage to care upon detection, % | 59·8 | 80·7 | 59·8 | [95,96] |
| Antibody test characteristics | | | | |
| Sensitivity, 0 months after infection, % | 0 | 0 | 0 | Assumption |
| Sensitivity, ≥1 month after infection, % | 100 | 100 | 100 | [97,98] |
| Specificity, % | | | | |
| Before seroconversion | 0·01 | 0·01 | 0·01 | Assumption |
| After seroconversion | 99 | 99 | 99 | [98] |
| Probability of result return, % | 99·6 | 99·5 | 99·7 | [93,95] |
| Delay between primary test and result return, mean mo. (SD) | 0 (0) | 0 (0) | 0 (0) | [93]/Assumption |
| Probability of linkage to care upon detection, % | 92·6 | 92·6 | 92·6 | [95] |
| Probability of HIV detection after presenting to care with a severe OI, % | 100 | 100 | 100 | Assumption |
| **Infant HIV prophylaxis** | | | | |
| Probability of receiving scheduled prophylaxis, % | | | | |
| Standard-of-care oral infant prophylaxis (NVP +/- ZDV) [‡] | 86 | 86 | 86 | [99] |
| bNAb prophylaxis (varies by age)[§] | 56-83 | 85-96 | 71-92 | [42–44,100] |
| Efficacy of standard-of-care oral infant prophylaxis (NVP +/- ZDV)[‡,¶] % | | | | [14,15] |
| Against intrapartum transmission | 69 | 69 | 69 | |
| Against postnatal transmission | 71 | 71 | 71 | |
| Efficacy of bNAb prophylaxis against intrapartum and postnatal transmission (in addition to efficacy of standard-of-care), % | 70 | 70 | 70 | Assumption based on [20,21] |
| Duration of bNAb effect, mo. | 3 | 3 | 3 | [16,17] |
| Probability of infant standard-of-care oral prophylaxis major toxicity, one-time % | 0 | 0 | 0 | Assumption based on [101–103] |
| Probability of infant bNAb prophylaxis minor toxicity, one-time % | 50 | 50 | 50 | Assumption |
| Probability of infant bNAb prophylaxis major toxicity, one-time % | 0 | 0 | 0 | Assumption |
| **Natural history** | | | | |
| Monthly CD4% or CD4 decline in absence of ART, by age | | | | |
| <5 years (range by age and time of transmission), % | 0·5 – 4·0 | 0·5 – 4·0 | 0·5 – 4·0 | [1] |
| ≥5 years, mean (SD) (range by viral load and CD4), cells/mm$^3$ | 3·0 (0·3) – 6·4 (0·3) | 3·0 (0·3) – 6·4 (0·3) | 3·0 (0·3) – 6·4 (0·3) | [1,104] |
| Monthly risk of clinical events, by age, % | | | | |
| 0-5 years (range by age and CD4%) | | | | [77] |

| Parameter | Côte d'Ivoire | South Africa | Zimbabwe | Reference(s) |
|---|---|---|---|---|
| WHO stage 3 event (except tuberculosis) | 3·3 – 11·6 | 3·3 – 11·6 | 3·3 – 11·6 | |
| WHO stage 4 event (except tuberculosis) | 1·4 – 6·4 | 1·4 – 6·4 | 1·4 – 6·4 | |
| Tuberculosis (any body site) | 0·5 – 3·8 | 0·5 – 3·8 | 0·5 – 3·8 | |
| ≥5 years (range by CD4) | | | | [105,106] |
| Mild fungal infection | 0·75 – 9·42 | 1·76 – 3·14 | 1·8 – 3·1 | |
| Mild bacterial infection | 0·93 – 2·01 | 0 | 0 | |
| Bacterial gastroenteritis | 0·18 – 1·33 | 0 | 0 | |
| Malaria | 0·85 – 2·97 | 0 | 0 | |
| Visceral bacterial infection | 0·15 – 1·70 | 0·04 – 0·71 | 0·04 – 0·71 | |
| WHO stage 3 or 4 visceral disease | 0·03 – 2·92 | 0·03 – 1·43 | 0·03 – 1·43 | |
| WHO stage 3 or 4 mucocutaneous disease | 0·01 – 2·74 | 0·03 – 2·26 | 0·03 – 2·26 | |
| Other WHO stage 3 or 4 disease | 0·04 – 2·08 | 0·02 – 0·73 | 0·02 – 0·73 | |
| Other severe disease | 0·75 – 3·79 | 0·19 – 1·67 | 0·19 – 1·67 | |
| Other mild infection | 0·72 – 4·16 | 2·39 | 2·39 | |
| Tuberculosis (any body site) | 0·06 – 0·66 | 0·03 – 1·74 | 0·03 – 1·74 | |
| Risk of death within 30 days of clinical event, by age, % | | | | |
| 0-5 years | | | | [1,77] |
| WHO stage 3 or 4 event | 13·5 | 13·5 | 13·5 | |
| Tuberculosis | 11·1 | 11·1 | 11·1 | |
| ≥5 years (range by CD4) | | | | [105,106] |
| Mild fungal infection | 0 | 0 | 0 | |
| Mild bacterial infection | 0 | ... | ... | |
| Bacterial gastroenteritis | 0 | ... | ... | |
| Malaria | 0 – 16·7 | ... | ... | |
| Visceral bacterial infection | 0 – 16·7 | 0 | 0 | |
| WHO stage 3 or 4 visceral disease | 6·7 – 12·3 | 0·5 | 0·5 | |
| WHO stage 3 or 4 mucocutaneous disease | 0 | 0 | 0 | |
| Other WHO stage 3 or 4 disease | 6·7 – 12·3 | 0 | 0 | |
| Other severe disease | 0 – 16·7 | 0·4 | 0·4 | |
| Other mild infection | 0 | 2·4 | 2·4 | |
| Tuberculosis (any body site) | 6·5 – 50·0 | 9·2 | 9·2 | |
| Monthly risk of HIV-related mortality, by age, % (range by CD4 and OI history) | | | | [1,77,105,106] |
| 0-5 months | 5·3 – 40·8 | 5·3 – 40·8 | 5·3 – 40·8 | |
| 6-11 months | 2·2 – 16·8 | 2·2 – 16·8 | 2·2 – 16·8 | |
| 12-23 months | 0·9 – 7·2 | 0·9 – 7·2 | 0·9 – 7·2 | |
| 24-35 months | 0·5 – 3·6 | 0·5 – 3·6 | 0·5 – 3·6 | |
| 36-47 months | 0·4 – 2·9 | 0·4 – 2·9 | 0·4 – 2·9 | |
| 48-59 months | 0·1 – 1·0 | 0·1 – 1·0 | 0·1 – 1·0 | |
| 5-13 years | 0·1 – 1·0 | 0·1 – 1·0 | 0·1 – 1·0 | |
| ≥13 years | 0·04 – 5·4 | 0·2 – 9·5 | 0·2 – 9·5 | |
| Monthly risk of non-HIV related mortality, % | | | | |
| HIV-exposed, uninfected children, born to mothers not on ART, by age | | | | [107,108] |
| 0-2 months | 1·01 | 1·01 | 1·01 | |
| 3-5 months | 0·41 | 0·41 | 0·41 | |
| 6-11 months | 0·28 | 0·28 | 0·28 | |
| 12-17 months | 0·14 | 0·14 | 0·14 | |
| 18-23 months | 0·07 | 0·07 | 0·07 | |
| ≥24 months | See below | See below | See below | |
| All others, by age | | | | [109,110] |
| 0-11 months (range by age and sex) | 0·50 – 0·66 | 0·24 – 0·31 | 0·30 – 0·38 | |
| 12-23 months (range by sex) | 0·05 – 0·06 | 0·02 | 0·03 | |
| 24-59 months (range by sex) | 0·05 – 0·06 | 0·02 | 0·03 | |
| 5-13 years (range by age and sex) | 0·02 – 0·03 | 0·003 – 0·006 | 0·01 – 0·02 | |

| Parameter | Côte d'Ivoire | South Africa | Zimbabwe | Reference(s) |
|---|---|---|---|---|
| 13-99 years | 0·02 – 2·40 | 0·004 – 1·930 | 0·01 – 1·89 | |
| 100 years | 100 | 100 | 100 | |
| **Treatment** | | | | |
| Efficacy of adult OI prophylaxis (co-trimoxazole) (range by OI type), % | 18 – 88 | 18 – 50 | 18 – 88 | [106,111] |
| Probability of adult OI prophylaxis (co-trimoxazole) minor toxicity, one-time % | 17 | 17 | 17 | [106,111] |
| Probability of adult OI prophylaxis (co-trimoxazole) major toxicity, one-time % | 6 | 6 | 6 | [106,111] |
| Probability of virologic suppression (<1,000 c/mL for children, <50 c/mL for adults) at 48 weeks while on ART, one-time % | | | | |
| Pediatric 1st line ART (LPV/r or EFV-based) | 76 | 76 | 76 | [112–118] |
| Pediatric 2nd and subsequent lines of ART (DTG-based) | 87 | 87 | 87 | [119] |
| Adult 1st line ART (DTG-based) | 90 | 90 | 90 | [120,121] |
| Adult 2nd and subsequent lines of ART (PI-based) | 83 | 83 | 83 | [122] |
| Probability of virologic failure after initial suppression, %/mo. | | | | |
| Pediatric 1st line ART | 0·52 | 0·52 | 0·52 | [113,116] |
| Pediatric 2nd and subsequent lines of ART | 0·37 | 0·37 | 0·37 | [119] |
| Adult 1st line ART | 0·60 | 0·60 | 0·60 | [120,121,123,124] |
| Adult 2nd and subsequent lines of ART | 0·20 | 0·20 | 0·20 | [125] |
| Probability of resuppression when reinitiating ART, one-time % | | | | |
| Pediatric ART lines | 31 | 31 | 31 | [126] |
| Adult ART lines | 67 | 67 | 67 | [127,128] |
| Effect of ART on CD4 when suppressed, by age | | | | |
| 0-4 years, absolute CD4% gain/mo., mean (SD) | | | | |
| 0-6 months after ART initiation | 2·2 (0·6) | 2·2 (0·6) | 2·2 (0·6) | [129,130] |
| ≥6 months after ART initiation | 0·7 (0·2) | 0·7 (0·2) | 0·7 (0·2) | [129,130] |
| ≥5 years, absolute CD4 gain/mo., mean (SD) | | | | |
| 0-2 months after ART initiation, cells/mm$^3$ | 83·2 (38·2) | 83·2 (38·2) | 83·2 (38·2) | [131] |
| ≥2 months after ART initiation, cells/mm$^3$ | 4·2 (1·9) | 4·2 (1·9) | 4·2 (1·9) | [131] |
| Relative risk reduction of HIV-related mortality for patients on ART, by age, % | | | | |
| 0-5 years | 90 | 90 | 90 | [1] |
| 5-13 years | 90 | 90 | 90 | [1] |
| ≥13 years (range by viral load and CD4) | 55 – 96 | 55 – 96 | 55 – 96 | [132] |
| Relative risk reduction of clinical events for patients on ART, by age, % | | | | |
| 0-5 years | 85 | 85 | 85 | [1] |
| 5-13 years (range by OI type) | 0 – 85 | 0 – 85 | 0 – 85 | [1] |
| ≥13 years (range by OI type) | 0 – 32 | 0 – 32 | 0 – 32 | [132] |
| Loss to follow-up probability after ART initiation, by age, %/mo. | | | | |
| 0-13 years | 1·6 | 0·8 | 0·4 | [133–142] |
| ≥13 years | 1·0 | 2·1 | 1·1 | [143–147] |
| Return to care probability after 6 months of loss to follow-up, %/mo. | | | | |
| With presentation of a severe OI | 100 | 100 | 100 | Assumption |
| Without presentation of a severe OI | 1·3 | 1·3 | 1·3 | [148] |
| Frequency of CD4 monitoring while in care, test interval, mo. | | | | [80] |
| First 12 months post-ART initiation | 6 | 6 | 6 | |
| After 12 months post-ART initiation | 12 | 12 | 12 | |
| CD4 threshold to stop CD4 monitoring, cells/mm$^3$ | >350 | >350 | >350 | |
| Frequency of viral load monitoring while in care, mo. | | | | [80] |
| First 12 months post-ART initiation | 6 | 6 | 6 | |
| After 12 months post-ART initiation | 12 | 12 | 12 | |
| **Costs (in 2020 USD)** | | | | |
| Early infant diagnosis program, $/test | | | | |
| Nucleic acid amplification test | 26·06 | 26·06 | 26·06 | [149] |
| Antibody test | 4·06 | 4·06 | 4·06 | [150] |
| Negative result return | 2·09 | 2·09 | 2·09 | [151] |

| Parameter | Côte d'Ivoire | South Africa | Zimbabwe | Reference(s) |
|---|---|---|---|---|
| Positive result return | 3·48 | 3·48 | 3·48 | |
| Pediatric ART (range by age and weight, per mo.) | | | | |
| 1st line | 5·65 – 11·70 | 5·65 – 11·70 | 5·65 – 11·70 | 152,153 |
| 2nd line | 12·43 – 25·99 | 12·43 – 25·99 | 12·43 – 25·99 | 152,153 |
| Adult ART (per mo.) | | | | |
| 1st line | 5·50 | 5·50 | 5·50 | 154 |
| 2nd line | 23·00 | 23·00 | 23·00 | 154 |
| Infant oral prophylaxis (NVP +/- ZDV), per mo. | 0·91 – 1·74 | 0·91 – 1·74 | 0·91 – 1·74 | 152,153 |
| bNAb prophylaxis, per dose | 20·00 | 20·00 | 20·00 | 24–27 |
| Routine HIV care, by age, $/mo. (range by CD4%/CD4 and ART status) | | | | |
| 0-18 years | 4·59 – 181·90 | 3·73 – 148·88 | 7·04 – 35·18 | 105,155–158 |
| ≥18 years | 4·16 – 28·48 | 3·73 – 148·88 | 7·04 – 35·18 | 105,106,156–158 |
| Routine CD4 monitoring test, $/test | 3·98 | 3·98 | 3·98 | 159,160 |
| Routine HIV viral load test, $/test | 24·05 | 24·05 | 24·05 | 149 |
| Adult OI prophylaxis (co-trimoxazole), $/mo. | 1·04 | 1·04 | 1·04 | 161 |
| Acute OI care (range by OI type), $/mo. | | | | |
| 0-5 years | 128·96 – 456·65 | 867·99 – 1525·15 | 0 | 155,162,163 |
| ≥5 years | 82·51 – 574·89 | 230·20 – 780·51 | 0 | 105,106,158 |
| Toxicity, $/event | | | | |
| Standard-of-care oral infant prophylaxis, minor (outpatient visit) | 0 | 0 | 0 | 106,158,164 |
| Infant bNAb prophylaxis, minor/major (outpatient visit) | 2·72 | 22·03 | 1·93 | 106,158,164 |
| Adult OI oral prophylaxis (co-trimoxazole), minor (outpatient visit) | 2·72 | 22·03 | 1·93 | 106,158,164 |
| Adult OI oral prophylaxis (co-trimoxazole), major (hospitalization) | 83·60 | 1611·45 | 52·61 | 106,158,164 |
| Death-related costs, $/death | 90·49 | 586·16 | 0 | 106,111,158,164 |
| Cost of identifying high-risk HIV-exposed infant | 31.66 | 31.66 | 31.66 | 26,27,149,151 |

*ART, antiretroviral therapy; mo, month; SD, standard deviation; WHO, World Health Organization; OI, opportunistic infection; NVP, nevirapine; ZDV, zidovudine; bNAb, broadly neutralizing antibody; LPV/r, lopinavir/ritonavir; EFV, efavirenz; DTG, dolutegravir; PI, protease inhibitor.*

† All women included in this analysis are known to have HIV infection by delivery. Therefore, we did not model acute HIV infection during breastfeeding.

ϕ Postnatal transmission risks among infants whose mothers are on ART while breastfeeding account for the potential impact of antiretrovirals transferred through breastmilk.

‡ Infants with known, high-risk HIV exposure received dual oral infant prophylaxis with nevirapine (NVP) + zidovudine (ZDV) for 12 weeks. Infants with known, low-risk HIV exposure received NVP alone for 6 weeks.

§ The probability of receiving broadly neutralizing antibody (bNAb) prophylaxis was based on the probability of receiving World Health Organization (WHO) Expanded Program on Immunization vaccines, by age at recommended immunization.

¶ A reduction in the risk of intrapartum transmission with use of oral infant prophylaxis was only applied to infants born to mothers who were known to have HIV infection, but who were not on antiretroviral therapy (ART) at delivery. The impact of oral infant prophylaxis on intrapartum transmission among mothers on ART at delivery is already captured in the on ART perinatal transmission estimates.

**Supplementary Table 3.** Justification of sensitivity analysis ranges

| Model input parameter | Base case value | Sensitivity analysis range | Justification |
|---|---|---|---|
| Antiretroviral therapy (ART) costs (per mo.) | Pediatric ART:<br>*1st Line:* \$5·65 - \$11·70<br>*2nd Line:* \$12·43 - \$25·99<br><br>Adult ART:<br>*1st Line:* \$5·50<br>*2nd Line:* \$23·00 | 0·5x – 2·0x | By halving and doubling all base case ART costs, we aim to capture a plausible range of potential cost changes due to reasons such as new formulations coming to market or price adjustments of existing formulations. |
| Broadly neutralizing antibody (bNAb) cost (per dose) | \$20 | \$5 - \$100 | This range was chosen to capture the lower and upper bounds of projected bNAb costs: \$6·26 and \$27·32, respectively (see appendix pp 5-6). Given that our bNAb efficacy estimate assumes a three-bNAb product, we also wanted to capture the price if production cost for each bNAb was separate and equal to the upper bound (i.e., 3 x \$20 for production alone). |
| BNAb effect duration | 3 mo. | 1 mo. - 6 mo. | A single, subcutaneous 20 mg/kg dose of VRC01—an older generation, single bNAb product—achieved the mean serum concentration target in infants of >50 ug/mL for 28 days following administration.[18] VRC01's pharmacokinetic data informed the lower bound of the sensitivity analysis range, while the upper bound was 2x the suggested efficacy duration of newer generation antibodies, such as VRC07-523LS, from similar pharmacokinetic studies.[16] |
| BNAb efficacy | 70% | 10% - 100% | We varied bNAb efficacy widely in increments of 10% from 10% to 100% to capture all potential efficacies of a bNAb product used for infant HIV prophylaxis. |
| BNAb uptake (varying by age) | Côte d'Ivoire:<br>56% - 83%<br><br><br>South Africa:<br>85% - 96%<br><br><br>Zimbabwe:<br>71% - 92% | Côte d'Ivoire:<br>56% at all time points – 83% at all time points<br><br>South Africa:<br>85% at all time points – 92% at all time points<br><br>Zimbabwe:<br>71% at all time points – 92% at all time points | The lower bound of bNAb uptake was informed by a linear regression model of average uptake of all country-specific recommended vaccinations through 18 months.[100] The upper bound was informed by the average uptake of recommended vaccines at birth for South Africa or the proportion of infants delivered in a health facility for Côte d'Ivoire and Zimbabwe reported by UNICEF.[42,44] |
| Mean breastfeeding duration | Côte d'Ivoire:<br>*Women with HIV:* 14 mo.<br>*Women w/o HIV:* 19 mo.<br>South Africa:<br>*Women with HIV:* 6 mo.<br>*Women w/o HIV:* 12 mo.<br><br>Zimbabwe:<br>*Women with HIV:* 13 mo.<br>*Women w/o HIV:* 18 mo. | Côte d'Ivoire:<br>*Women with HIV:* 8 mo. - 24 mo.<br>*Women w/o HIV:* 8 mo. - 24 mo.<br>South Africa:<br>*Women with HIV:* 2 mo. - 18 mo.<br>*Women w/o HIV:* 2 mo. - 18 mo.<br><br>Zimbabwe:<br>*Women with HIV:* 8 mo. - 20 mo.<br>*Women w/o HIV:* 15 mo. - 20 mo. | In Côte d'Ivoire, we varied the range of mean breastfeeding duration to capture all plausible scenarios in a clinical setting. The same range was used for all women, irrespective of HIV status. We chose a lower bound of 8 months based on clinical assumptions. The upper bound was partially informed by subnational Multiple Indicator Cluster Survey (MICS) estimates: Nord-Est region has the longest duration of breastfeeding (21·5 months).[42]<br><br>In South Africa, the range for women without HIV was informed by published literature and survey estimates. In the Mother-Infant Health Study, women without HIV predominantly breastfed for 2 months, informing the lower bound.[166] Demographic Health Survey (DHS) data reported that the longest median duration of breastfeeding among any subgroup was 16·5 months among women in the lowest wealth quintile, which was used to inform the upper bound.[43]<br><br>In Zimbabwe, the range for women without HIV was informed by provincial MICS data: Harare had the shortest breastfeeding duration (15·9 months) and Mashonaland Central had the longest breastfeeding duration (19·4 months).[44] The range for women with HIV was informed by the IQR of a nationally representative sample of mother and HIV-exposed infant pairs between 2013 and 2014.[79] |

| Model input parameter | Base case value | Sensitivity analysis range | Justification |
|---|---|---|---|
| Cost of ascertaining high-risk status | $31·66 | $0 - $31·66 | The cost of ascertaining whether an infant has low- or high-risk HIV exposure was assumed to include the cost of a maternal viral load test. The cost components included the cost of a viral load test ($24·05), result return with counseling ($3·48), and personnel/overhead ($4·13), reported in 2020 USD.[26,27,149,151] In sensitivity analyses, we wanted to assess whether assuming zero additional cost would make the *HR-HIVE* strategies cost-effective relative to other bNAb strategies. We chose not to increase the cost of ascertaining risk status because, compared to the base case results, it would only make the *HR-HIVE* strategies even more costly and less clinically beneficial than other strategies (i.e., dominated). |
| Maternal ART coverage during pregnancy | Côte d'Ivoire: 95%<br><br>South Africa: 97%<br><br>Zimbabwe: 89% | Côte d'Ivoire: 70% - 100%<br><br>South Africa: 60% - 100%<br><br>Zimbabwe: 65% - 100% | We chose the ranges for maternal ART coverage during pregnancy based on the 95% confidence intervals from UNAIDS 2021 data (Côte d'Ivoire: 70 - >98%; South Africa: 60 - >98%; Zimbabwe: 68 - >98%).[54] |
| Maternal HIV prevalence and incidence | Côte d'Ivoire:<br>*Prevalence at delivery:* 2·6%<br>*Incidence postpartum:* 0·003%/mo.<br><br>South Africa:<br>*Prevalence at delivery:* 33·0%<br>*Incidence postpartum:* 0·240%/mo.<br><br>Zimbabwe:<br>*Prevalence at delivery:* 12·3%<br>*Incidence postpartum:* 0·056%/mo. | Côte d'Ivoire:<br>*Prevalence at delivery:* 1·0% - 5·0%<br>*Incidence postpartum:*<br>0·001%/mo. – 0·005%/mo.<br><br>South Africa:<br>*Prevalence at delivery:* 15·0% - 45·0%<br>*Incidence postpartum:*<br>0·107%/mo. – 0·331%/mo.<br><br>Zimbabwe:<br>*Prevalence at delivery:* 5·0% - 25·0%<br>*Incidence postpartum:*<br>0·022%/mo. – 0·116%/mo. | We chose the ranges for maternal HIV prevalence and incidence based on subnational estimates among women ages 15-49 years old from Population-Based HIV Impact Assessments (PHIA) data for Côte d'Ivoire and Zimbabwe and National Antenatal Sentinel HIV Survey data for South Africa. In our analysis, maternal HIV prevalence at delivery is inclusive of prevalence and incidence during pregnancy. We applied a multiplier to the base case estimate of HIV prevalence at delivery such that it captured the range of subnational estimates. We assumed the same multiplier applies to incidence postpartum.<br><br>In Côte d'Ivoire, Woroba had the lowest prevalence (1·5%) and Montagnes had the highest prevalence (5·0%).[55] In South Africa, Western Cape province had the lowest prevalence (15·9%) KwaZulu-Natal province had the highest prevalence (41·1%).[49] In Zimbabwe, the lower bound of maternal HIV prevalence was informed by the lower 95% confidence interval estimate of maternal HIV prevalence (9·9%). The HIV prevalence for women in Matabeleland South (24·2%) informs the upper bound.[53] |
| Maternal knowledge of acute HIV infection | Côte d'Ivoire:<br>*During pregnancy:* 56%<br>*Postpartum:* 2%/mo.<br><br>South Africa:<br>*During pregnancy:* 55%<br>*Postpartum:* 9%/mo.<br><br>Zimbabwe:<br>*During pregnancy:* 70%<br>*Postpartum:* 5%/mo. | All countries:<br>*During pregnancy:* 25% - 95%<br>*Postpartum:* 0%/mo. – 15%/mo. | The lower bound of maternal knowledge of acute HIV infection during pregnancy was informed by published literature. In a study from Kenya, 26·7% of women who initially tested HIV-negative in antenatal care and returned to antenatal care to retest prior to delivery.[39] We aimed to capture all plausible scenarios of maternal knowledge of acute HIV infection by setting the upper bound of obtaining knowledge during pregnancy at 95% and during the postpartum period at 15%/mo. |
| Maternal knowledge of chronic HIV infection | Côte d'Ivoire:<br>93%<br><br>South Africa:<br>99%<br><br>Zimbabwe:<br>98% | Côte d'Ivoire:<br>56-100%<br><br>South Africa:<br>75-100%<br><br>Zimbabwe:<br>80-100% | The ranges for maternal knowledge of chronic HIV infection were informed by the upper and lower bounds of 95% confidence intervals for knowledge of HIV status as reported in South Africa DHS 2016 and UNAIDS 2020 data.[43,60] |
| Oral infant prophylaxis efficacy | Against intrapartum transmission: 69%<br><br>Against postnatal transmission: 71% | Against intrapartum transmission: 40% - 90%<br><br>Against postnatal transmission: 58% - 80% | We chose the range of oral infant prophylaxis efficacy against intrapartum transmission based on ranges reported in the PEPI, HPTN 040, and PHT-5 studies.[14,167,168] |

| Model input parameter | Base case value | Sensitivity analysis range | Justification |
|---|---|---|---|
| Oral infant prophylaxis efficacy (continued) | | | The range for oral infant prophylaxis efficacy against intrapartum transmission was informed by the 95% confidence interval of relative risk reduction for infants given a short course of nevirapine (NVP) + zidovudine (ZDV) compared to a single dose of NVP.[14] The range for oral infant prophylaxis efficacy against postnatal transmission was informed by the 95% confidence interval of infant HIV infection for infants given 6 weeks of NVP compared to no oral infant prophylaxis.[15] |
| Postnatal vertical transmission risk | On ART<br>*Acute HIV*: 4·0%<br>*Chronic HIV, suppressed*: 0·06%<br>*Chronic HIV, not suppressed*:<br>0·39% – 0·78%<br><br>Off ART<br>*Acute HIV*: 4·6%<br>*Chronic HIV*: 0·89% | 0·5x – 2·0x | By halving and doubling all base case postnatal vertical transmission risks, we aim to capture a plausible range of potential transmission risks due to reasons such as adherence to ART regimens or varying HIV viral loads. |
| Postpartum maternal ART retention | Côte d'Ivoire:<br>*Known, high-risk / low-risk:*<br>*0 mo.*: 86% / 100%<br>*6 mo.*: 87%<br>*12 mo.*: 87%<br>*24 mo.*: 79%<br><br>*Unrecognized exposed and unexposed:*<br>*0 mo.*: 0%<br>*6 mo.*: 9%<br>*12 mo.*: 17%<br>*24 mo.*: 27%<br><br>South Africa:<br>*Known, high-risk / low-risk:*<br>*0 mo.*: 87% / 100%<br>*6 mo.*: 88%<br>*12 mo.*: 87%<br>*24 mo.*: 79%<br><br>*Unrecognized exposed and unexposed:*<br>*0 mo.*: 0%<br>*6 mo.*: 37%<br>*12 mo.*: 57%<br>*24 mo.*: 69%<br><br>Zimbabwe:<br>*Known, high-risk / low-risk:*<br>*0 mo.*: 59% / 100%<br>*6 mo.*: 88%<br>*12 mo.*: 87%<br>*24 mo.*: 79%<br><br>*Unrecognized exposed and unexposed:*<br>*0 mo.*: 0%<br>*6 mo.*: 19%<br>*12 mo.*: 33%<br>*24 mo.*: 47% | All countries:<br>*Known, high-risk and low-risk*<br>*6+ mo.*: 50% - 100%<br><br>Côte d'Ivoire:<br>*Unrecognized exposed and unexposed:*<br>*0 mo.*: 0%<br>*6 mo.*: 5% - 10%<br>*12 mo.*: 10% - 19%<br>*24 mo.*: 17% - 35%<br><br>South Africa:<br>*Unrecognized exposed and unexposed:*<br>*0 mo.*: 0%<br>*6 mo.*: 21% - 42%<br>*12 mo.*: 33% - 66%<br>*24 mo.*: 44% - 87%<br><br>Zimbabwe:<br>*Unrecognized exposed and unexposed:*<br>*0 mo.*: 0%<br>*6 mo.*: 11% - 21%<br>*12 mo.*: 19% - 38%<br>*24 mo.*: 30% - 59% | We varied postpartum maternal ART retention for the known, high-risk and low-risk cohorts from 50% to 100% after 6 months to capture all plausible scenarios in a clinical setting. For the unrecognized as HIV exposed and unexposed cohorts, we calculated the ranges based on receiving ART during pregnancy if HIV status is known and maternal knowledge of HIV during breastfeeding.[43,53–55] |

| Model input parameter | Base case value | Sensitivity analysis range | Justification |
|---|---|---|---|
| Postpartum maternal HIV incidence | Côte d'Ivoire:<br>0·003%/mo.<br><br>South Africa:<br>0·240%/mo.<br><br>Zimbabwe:<br>0·056%/mo. | 0·5x – 2·0x | By halving and doubling all base case postpartum maternal HIV incidence, we aim to capture a plausible range of postpartum incidence changes due to reasons such as subnational incidence rates and missing cases in survey data. |

*mo.: month; w/o: without; IQR: interquartile range.*

**Supplementary Table 4.** Validation of CEPAC-P infant HIV vertical transmission projections

| Country | CEPAC-P projected vertical transmission rate in the standard of care (SOC) strategy, % | UNAIDS 2021 estimates, % (95% CI)[62] |
|---|---|---|
| Côte d'Ivoire | 8·4 | 8 (4-12) |
| South Africa | 4·4 | 4 (3-8) |
| Zimbabwe | 9·5 | 9 (6-12) |

*SOC: standard-of-care; CI: confidence interval.*
Model-projected estimates of vertical transmission are often higher than those observed in clinical studies since they capture infants who may have acquired HIV infection, but be undiagnosed due to loss to follow up from EID programs or death prior to diagnosis.

**Supplementary Table 5.** Clinical and economic outcomes of all modeled bNAb infant prophylaxis programs, by country

| | Clinical outcomes | | | Lifetime efficacy and costs | | | |
|---|---|---|---|---|---|---|---|
| **Country/strategy** | **IU/IP cumulative HIV incidence (%)** | **Postnatal cumulative HIV incidence (%)** | **Total cumulative HIV incidence (%)** | **Undiscounted life expectancy (yrs)** | **Discounted life expectancy (yrs)** | **Discounted costs ($)** | **ICER ($/YLS)** |
| **Côte d'Ivoire [CET: ICER ≤ $465/YLS (20% GDP per capita), ICER ≤ $1163/YLS (50% GDP per capita)]** | | | | | | | |
| Standard-of-care | 3·6 | 5·0 | 8·4 | 60·129 | 26·373 | 15 | Reference |
| HR-HIVE – 1 dose | 3·3 | 4·8 | 8·0 | 60·133 | 26·375 | 15 | dominated |
| HR-HIVE – 2 doses | 3·3 | 4·7 | 7·8 | 60·134 | 26·375 | 15 | dominated |
| HIVE – 1 dose | 3·3 | 4·7 | 7·8 | 60·134 | 26·375 | 14 | dominated |
| HIVE – 1 dose plus HR-HIVE – 2 doses | 3·3 | 4·6 | 7·6 | 60·136 | 26·376 | 15 | dominated |
| HR-HIVE – Extended | 3·3 | 4·3 | 7·5 | 60·136 | 26·376 | 14 | dominated |
| HIVE – 2 doses | 3·3 | 4·3 | 7·4 | 60·138 | 26·377 | 14 | dominated |
| HIVE – 1 dose plus HR-HIVE Extended | 3·3 | 4·2 | 7·3 | 60·138 | 26·377 | 14 | dominated |
| ALL – 1 dose | 3·0 | 4·6 | 7·5 | 60·139 | 26·377 | 27 | dominated |
| ALL – 1 dose plus HR-HIVE – 2 doses | 3·0 | 4·5 | 7·3 | 60·140 | 26·378 | 28 | dominated |
| ALL – 2 doses | 3·0 | 4·2 | 7·0 | 60·140 | 26·378 | 43 | dominated |
| ALL – 1 dose plus HIVE – 2 doses | 3·0 | 4·3 | 7·1 | 60·143 | 26·379 | 27 | dominated |
| ALL – 1 dose plus HR-HIVE - Extended | 3·0 | 4·1 | 7·0 | 60·143 | 26·379 | 28 | dominated |
| HIVE – Extended | 3·3 | 3·2 | 6·3 | 60·146 | 26·379 | 13 | cost-saving*·† |
| ALL – 1 dose plus HIVE - Extended | 3·0 | 3·1 | 6·0 | 60·150 | 26·382 | 26 | 6242 |
| ALL - Extended | 3·0 | 2·7 | 5·5 | 60·156 | 26·383 | 91 | 57 860 |
| **South Africa [CET: ICER ≤ $1131/YLS (20% GDP per capita), ICER ≤ $2828/YLS (50% GDP per capita)]** | | | | | | | |
| Standard-of-care | 2·3 | 2·2 | 4·4 | 68·924 | 28·499 | 112 | Reference |
| HR-HIVE – 1 dose | 2·1 | 2·1 | 4·0 | 68·979 | 28·518 | 115 | dominated |
| HR-HIVE – 2 doses | 2·1 | 2·1 | 4·0 | 68·986 | 28·521 | 115 | dominated |
| HR-HIVE – Extended | 2·1 | 2·0 | 3·9 | 68·995 | 28·523 | 114 | dominated |
| HIVE – 1 dose | 2·0 | 2·0 | 3·8 | 69·011 | 28·529 | 105 | dominated |
| HIVE – 1 dose plus HR-HIVE – 2 doses | 2·0 | 1·9 | 3·8 | 69·018 | 28·532 | 115 | dominated |
| HIVE – 1 dose plus HR-HIVE - Extended | 2·0 | 1·9 | 3·7 | 69·027 | 28·534 | 114 | dominated |
| HIVE – 2 doses | 2·0 | 1·8 | 3·6 | 69·045 | 28·541 | 104 | dominated |
| ALL – 1 dose | 1·8 | 1·8 | 3·5 | 69·063 | 28·548 | 116 | dominated |
| ALL – 1 dose plus HR-HIVE – 2 doses | 1·8 | 1·7 | 3·4 | 69·070 | 28·550 | 126 | dominated |
| ALL – 1 dose plus HR-HIVE - Extended | 1·8 | 1·7 | 3·4 | 69·079 | 28·553 | 125 | dominated |
| HIVE – Extended | 2·0 | 1·5 | 3·3 | 69·095 | 28·557 | 100 | cost-saving |
| ALL – 1 dose plus HIVE – 2 doses | 1·8 | 1·6 | 3·3 | 69·097 | 28·559 | 115 | dominated |
| ALL – 2 doses | 1·8 | 1·5 | 3·2 | 69·122 | 28·568 | 122 | dominated |
| ALL – 1 dose plus HIVE - Extended | 1·8 | 1·3 | 3·0 | 69·147 | 28·575 | 111 | 606 |
| ALL - Extended | 1·8 | 0·8 | 2·5 | 69·228 | 28·602 | 135 | 882*·† |

**Supplemental Table 5.** Clinical and economic outcomes of all modeled bNAb infant prophylaxis programs, by country (cont.)

| Country/strategy | Clinical outcomes: IU/IP cumulative HIV incidence (%) | Clinical outcomes: Postnatal cumulative HIV incidence (%) | Clinical outcomes: Total cumulative HIV incidence (%) | Lifetime efficacy and costs: Undiscounted life expectancy (yrs) | Lifetime efficacy and costs: Discounted life expectancy (yrs) | Lifetime efficacy and costs: Discounted costs ($) | Lifetime efficacy and costs: ICER ($/YLS) |
|---|---|---|---|---|---|---|---|
| **Zimbabwe [CET: ICER ≤ $243/YLS (20% GDP per capita), ICER ≤ $607/YLS (50% GDP per capita)]** | | | | | | | |
| Standard-of-care | 4·3 | 5·6 | 9·5 | 68·321 | 28·034 | 72 | Reference |
| HR-HIVE – 1 dose | 3·7 | 5·3 | 8·6 | 68·366 | 28·050 | 71 | dominated |
| HR-HIVE – 2 doses | 3·7 | 5·2 | 8·5 | 68·373 | 28·053 | 70 | dominated |
| HIVE – 1 dose | 3·7 | 5·2 | 8·4 | 68·380 | 28·055 | 67 | dominated |
| HR-HIVE – Extended | 3·7 | 4·9 | 8·2 | 68·387 | 28·057 | 69 | dominated |
| HIVE – 1 dose plus HR-HIVE – 2 doses | 3·7 | 5·0 | 8·3 | 68·387 | 28·057 | 70 | dominated |
| HIVE – 1 dose plus HR-HIVE - Extended | 3·7 | 4·7 | 8·0 | 68·401 | 28·062 | 69 | dominated |
| ALL – 1 dose | 3·4 | 4·9 | 7·9 | 68·400 | 28·062 | 79 | dominated |
| HIVE – 2 doses | 3·7 | 4·8 | 8·0 | 68·402 | 28·063 | 66 | dominated |
| ALL – 1 dose plus HR-HIVE – 2 doses | 3·4 | 4·7 | 7·8 | 68·408 | 28·065 | 83 | dominated |
| ALL – 1 dose plus HR-HIVE - Extended | 3·4 | 4·5 | 7·5 | 68·422 | 28·069 | 82 | dominated |
| ALL – 1 dose plus HIVE – 2 doses | 3·4 | 4·5 | 7·5 | 68·423 | 28·070 | 79 | dominated |
| ALL – 2 doses | 3·4 | 4·3 | 7·4 | 68·438 | 28·074 | 93 | dominated |
| HIVE – Extended | 3·7 | 3·6 | 6·9 | 68·458 | 28·081 | 61 | cost-saving*,† |
| ALL – 1 dose plus HIVE - Extended | 3·4 | 3·4 | 6·5 | 68·478 | 28·088 | 74 | 1731 |
| ALL - Extended | 3·4 | 2·6 | 5·7 | 68·517 | 28·101 | 134 | 4516 |

*bNAb: broadly neutralizing antibody; IU/IP: intrauterine/intrapartum; yr: year; ICER: incremental cost-effectiveness ratio; YLS: years of life saved; CET: cost-effectiveness threshold; HR-HIVE: high-risk HIV-exposed infants; HIVE: all HIV-exposed infants; ALL: all live infants at birth.*

Pediatric HIV incidence is rounded to the nearest tenth of a percent. IU/IP HIV incidence is calculated based on the number of infants exposed to HIV at birth. Postnatal and total HIV incidence is calculated based on the number of infants ever exposed to HIV through 36 months of life. Undiscounted and discounted life expectancies are rounded to the nearest ten thousandth. Costs are rounded to the nearest dollar and are presented in 2020 USD. Discounted values are discounted at 3% per year. ICERs are rounded to the nearest dollar and are calculated using unrounded discounted life expectancy and discounted costs. The cost-effective bNAb strategy was the strategy that offered the greatest increase in overall population life expectancy while still having an ICER less than the cost-effectiveness threshold when compared to the next best performing, non-dominated strategy.

*Indicates the cost-effective strategy using a cost-effectiveness threshold of 20% GDP per capita. †Indicates the cost-effective strategy using a cost-effectiveness threshold of 50% GDP per capita.

**Supplementary Table 6.** Scenario analysis: bNAbs do not reduce intrapartum transmission (base case: 70% reduction)

| | **Clinical outcomes** | | | **Lifetime efficacy and costs** | | | |
|---|---|---|---|---|---|---|---|
| **Country/strategy** | **IU/IP cumulative HIV incidence (%)** | **Postnatal cumulative HIV incidence (%)** | **Total cumulative HIV incidence (%)** | **Undiscounted life expectancy (yrs)** | **Discounted life expectancy (yrs)** | **Discounted costs ($)** | **ICER ($/YLS)** |
| **Côte d'Ivoire [CET: ICER ≤ $465/YLS (20% GDP per capita), ICER ≤ $1163/YLS (50% GDP per capita)]** | | | | | | | |
| Standard-of-care | 3·6 | 5·0 | 8·4 | 60·129 | 26·373 | 15 | Reference |
| HR-HIVE – 1 dose | 3·6 | 4·9 | 8·3 | 60·130 | 26·374 | 15 | dominated |
| HR-HIVE – 2 doses | 3·6 | 4·7 | 8·1 | 60·131 | 26·374 | 15. | dominated |
| HIVE – 1 dose | 3·6 | 4·7 | 8·1 | 60·131 | 26·374 | 15 | dominated |
| HR-HIVE – Extended | 3·6 | 4·4 | 7·8 | 60·134 | 26·375 | 15 | dominated |
| HIVE – 2 doses | 3·6 | 4·3 | 7·8 | 60·135 | 26·375 | 14 | dominated |
| ALL – 1 dose | 3·6 | 4·7 | 8·1 | 60·134 | 26·375 | 28 | dominated |
| ALL – 2 doses | 3·6 | 4·2 | 7·6 | 60·135 | 26·376 | 43 | dominated |
| HIVE – Extended | 3·6 | 3·2 | 6·6 | 60·142 | 26·378 | 13 | cost-saving*·† |
| ALL – 1 dose plus HIVE - Extended | 3·6 | 3·1 | 6·6 | 60·145 | 26·380 | 27 | 10 008 |
| ALL - Extended | 3·6 | 2·7 | 6·1 | 60·151 | 26·381 | 92 | 59 542 |
| **South Africa [CET: ICER ≤ $1131/YLS (20% GDP per capita), ICER ≤ $2828/YLS (50% GDP per capita)]** | | | | | | | |
| Standard-of-care | 2·3 | 2·2 | 4·4 | 68·924 | 28·499 | 112 | Reference |
| HR-HIVE – 1 dose | 2·3 | 2·1 | 4·3 | 68·938 | 28·504 | 121 | dominated |
| HR-HIVE – 2 doses | 2·3 | 2·1 | 4·2 | 68·945 | 28·506 | 121 | dominated |
| HR-HIVE – Extended | 2·3 | 2·0 | 4·2 | 68·953 | 28·509 | 120 | dominated |
| HIVE – 1 dose | 2·3 | 2·0 | 4·2 | 68·955 | 28·510 | 113 | dominated |
| ALL – 1 dose | 2·3 | 1·8 | 4·0 | 68·986 | 28·521 | 127 | dominated |
| HIVE – 2 doses | 2·3 | 1·8 | 4·0 | 68·989 | 28·522 | 112 | dominated |
| HIVE – Extended | 2·3 | 1·5 | 3·7 | 69·038 | 28·538 | 109 | cost-saving |
| ALL – 2 doses | 2·3 | 1·5 | 3·7 | 69·044 | 28·541 | 133 | dominated |
| ALL – 1 dose plus HIVE - Extended | 2·3 | 1·3 | 3·5 | 69·069 | 28·549 | 122 | dominated |
| ALL - Extended | 2·3 | 0·8 | 3·0 | 69·150 | 28·575 | 146 | 998*·† |
| **Zimbabwe [CET: ICER ≤ $243/YLS (20% GDP per capita), ICER ≤ $607/YLS (50% GDP per capita)]** | | | | | | | |
| Standard-of-care | 4·3 | 5·6 | 9·5 | 68·321 | 28·034 | 72 | Reference |
| HR-HIVE – 1 dose | 4·3 | 5·4 | 9·2 | 68·337 | 28·040 | 74 | dominated |
| HR-HIVE – 2 doses | 4·3 | 5·2 | 9·1 | 68·344 | 28·042 | 74 | dominated |
| HIVE – 1 dose | 4·3 | 5·2 | 9·0 | 68·346 | 28·043 | 71 | dominated |
| HR-HIVE – Extended | 4·3 | 4·9 | 8·8 | 68·358 | 28·046 | 73 | dominated |
| ALL – 1 dose | 4·3 | 4·9 | 8·8 | 68·357 | 28·046 | 85 | dominated |
| HIVE – 2 doses | 4·3 | 4·8 | 8·6 | 68·369 | 28·051 | 70 | dominated |
| ALL – 2 doses | 4·3 | 4·4 | 8·2 | 68·395 | 28·058 | 98 | dominated |
| HIVE – Extended | 4·3 | 3·7 | 7·6 | 68·424 | 28·068 | 65 | cost-saving*·† |
| ALL – 1 dose plus HIVE - Extended | 4·3 | 3·4 | 7·3 | 68·434 | 28·072 | 79 | 3855 |
| ALL - Extended | 4·3 | 2·6 | 6·5 | 68·472 | 28·085 | 139 | 4584 |

*bNAb: broadly neutralizing antibody; IU/IP: intrauterine/intrapartum; yr: year; ICER: incremental cost-effectiveness ratio; YLS: years of life saved; CET: cost-effectiveness threshold; HR-HIVE: high-risk HIV-exposed infants; HIVE: all HIV-exposed infants; ALL: all live infants at birth.*

Pediatric HIV incidence is rounded to the nearest tenth of a percent. IU/IP HIV incidence is calculated based on the number of infants exposed to HIV at birth. Postnatal and total HIV incidence is calculated based on the number of infants ever exposed to HIV through 36 months of life. Undiscounted and discounted life expectancies are rounded to the nearest ten thousandth. Costs are rounded to the nearest dollar and are presented in 2020 USD. Discounted values are discounted at 3% per year. ICERs are rounded to the nearest dollar and are calculated using unrounded discounted life expectancy and discounted costs. The cost-effective bNAb strategy was the strategy that offered the greatest increase in overall population life expectancy while still having an ICER less than the cost-effectiveness threshold when compared to the next best performing, non-dominated strategy.

*Indicates the cost-effective strategy using a cost-effectiveness threshold of 20% GDP per capita. †Indicates the cost-effective strategy using a cost-effectiveness threshold of 50% GDP per capita.

**Supplementary Table 7.** Scenario analysis: bNAbs replace WHO-recommended standard-of-care oral infant prophylaxis

| | Clinical outcomes | | | Lifetime efficacy and costs | | | |
|---|---|---|---|---|---|---|---|
| **Country/strategy** | **IU/IP cumulative HIV incidence (%)** | **Postnatal cumulative HIV incidence (%)** | **Total cumulative HIV incidence (%)** | **Undiscounted life expectancy (yrs)** | **Discounted life expectancy (yrs)** | **Discounted costs ($)** | **ICER ($/YLS)** |
| **Côte d'Ivoire [CET: ICER ≤ $465/YLS (20% GDP per capita), ICER ≤ $1163/YLS (50% GDP per capita)]** | | | | | | | |
| Standard-of-care | 3·6 | 5·0 | 8·4 | 60·129 | 26·373 | 15 | Reference |
| HR-HIVE – 1 dose | 3·3 | 5·0 | 8·1 | 60·131 | 26·374 | 15 | dominated |
| HR-HIVE – 2 doses | 3·3 | 4·9 | 8·0 | 60·132 | 26·374 | 15 | dominated |
| HIVE – 1 dose | 3·3 | 4·9 | 8·0 | 60·132 | 26·374 | 14 | dominated |
| HR-HIVE – Extended | 3·3 | 4·5 | 7·6 | 60·135 | 26·375 | 15 | dominated |
| ALL – 1 dose | 3·0 | 4·9 | 7·7 | 60·134 | 26·375 | 28 | dominated |
| HIVE – 2 doses | 3·3 | 4·5 | 7·6 | 60·136 | 26·376 | 14 | dominated |
| ALL – 2 doses | 3·0 | 4·4 | 7·2 | 60·144 | 26·378 | 43 | dominated |
| HIVE – Extended | 3·3 | 3·4 | 6·5 | 60·143 | 26·379 | 13 | cost-saving*,† |
| ALL – 1 dose plus HIVE - Extended | 3·0 | 3·3 | 6·2 | 60·145 | 26·379 | 26 | 15 769 |
| ALL - Extended | 3·0 | 2·9 | 5·8 | 60·156 | 26·383 | 91 | 21 344 |
| **South Africa [CET: ICER ≤ $1131/YLS (20% GDP per capita), ICER ≤ $2828/YLS (50% GDP per capita)]** | | | | | | | |
| Standard-of-care | 2·3 | 2·2 | 4·4 | 68·924 | 28·499 | 112 | Reference |
| HR-HIVE – 1 dose | 2·1 | 2·2 | 4·1 | 68·969 | 28·514 | 116 | dominated |
| HR-HIVE – 2 doses | 2·1 | 2·1 | 4·0 | 68·975 | 28·517 | 116 | dominated |
| HR-HIVE – Extended | 2·1 | 2·1 | 4·0 | 68·984 | 28·519 | 115 | dominated |
| HIVE – 1 dose | 2·0 | 2·1 | 3·9 | 69·000 | 28·525 | 106 | dominated |
| HIVE – 2 doses | 2·0 | 1·9 | 3·7 | 69·032 | 28·536 | 105 | dominated |
| ALL – 1 dose | 1·8 | 1·9 | 3·6 | 69·058 | 28·544 | 118 | dominated |
| HIVE – Extended | 2·0 | 1·6 | 3·4 | 69·080 | 28·552 | 102 | cost-saving |
| ALL – 2 doses | 1·8 | 1·5 | 3·2 | 69·107 | 28·562 | 123 | dominated |
| ALL – 1 dose plus HIVE - Extended | 1·8 | 1·4 | 3·1 | 69·138 | 28·571 | 113 | 587 |
| ALL - Extended | 1·8 | 0·9 | 2·6 | 69·211 | 28·596 | 136 | 941*,† |
| **Zimbabwe [CET: ICER ≤ $243/YLS (20% GDP per capita), ICER ≤ $607/YLS (50% GDP per capita)]** | | | | | | | |
| Standard-of-care | 4·3 | 5·6 | 9·5 | 68·321 | 28·034 | 72 | Reference |
| HR-HIVE – 1 dose | 3·7 | 5·6 | 8·9 | 68·349 | 28·044 | 72 | dominated |
| HR-HIVE – 2 doses | 3·7 | 5·5 | 8·8 | 68·356 | 28·046 | 72 | dominated |
| HIVE – 1 dose | 3·7 | 5·5 | 8·7 | 68·361 | 28·048 | 69 | dominated |
| HR-HIVE – Extended | 3·7 | 5·2 | 8·5 | 68·370 | 28·051 | 71 | dominated |
| ALL – 1 dose | 3·4 | 5·2 | 8·3 | 68·381 | 28·056 | 81 | dominated |
| HIVE – 2 doses | 3·7 | 5·1 | 8·3 | 68·383 | 28·056 | 68 | dominated |
| ALL – 2 doses | 3·4 | 4·7 | 7·7 | 68·411 | 28·066 | 95 | dominated |
| HIVE – Extended | 3·7 | 4·0 | 7·2 | 68·438 | 28·074 | 63 | cost-saving*,† |
| ALL – 1 dose plus HIVE - Extended | 3·4 | 3·7 | 6·8 | 68·457 | 28·081 | 76 | 1696 |
| ALL - Extended | 3·4 | 2·9 | 6·0 | 68·497 | 28·093 | 136 | 5020 |

*bNAb: broadly neutralizing antibody; IU/IP: intrauterine/intrapartum; yr: year; ICER: incremental cost-effectiveness ratio; YLS: years of life saved; CET: cost-effectiveness threshold; HR-HIVE: high-risk HIV-exposed infants; HIVE: all HIV-exposed infants; ALL: all live infants at birth.*

In this scenario, infants receive either bNAbs or WHO-recommended standard-of-care oral prophylaxis, but not both; sub-populations not eligible to receive bNAbs in a strategy continue to be eligible for WHO-recommended standard-of-care oral prophylaxis. For example, in the HR-HIVE strategy, infants who are known, high-risk, HIV-exposed could only receive bNAbs but not oral prophylaxis while all other sub-populations would still be eligible to receive WHO-recommended standard-of-care oral prophylaxis. Pediatric HIV incidence is rounded to the nearest tenth of a percent. IU/IP HIV incidence is calculated based on the number of infants exposed to HIV at birth. Postnatal and total HIV incidence is calculated based on the number of infants ever exposed to HIV through 36 months of life. Undiscounted and discounted life expectancies are rounded to the nearest ten thousandth. Costs are rounded to the nearest dollar and are presented in 2020 USD. Discounted values are discounted at 3% per year. ICERs are rounded to the nearest dollar and are calculated using unrounded discounted life expectancy and discounted costs. The cost-effective bNAb strategy was the strategy that offered the greatest increase in overall population life expectancy while still having an ICER less than the cost-effectiveness threshold when compared to the next best performing, non-dominated strategy. *Indicates the cost-effective strategy using a cost-effectiveness threshold of 20% GDP per capita. †Indicates the cost-effective strategy using a cost-effectiveness threshold of 50% GDP per capita.

**Supplementary Table 8.** Scenario analysis: maternal HIV prevalence and incidence

| Country/strategy | Clinical outcomes: IU/IP cumulative HIV incidence (%) | Postnatal cumulative HIV incidence (%) | Total cumulative HIV incidence (%) | Lifetime efficacy and costs: Undiscounted life expectancy (yrs) | Discounted life expectancy (yrs) | Discounted costs ($) | ICER ($/YLS) |
|---|---|---|---|---|---|---|---|
| **Côte d'Ivoire [CET: ICER ≤ $465/YLS (20% GDP per capita), ICER ≤ $1163/YLS (50% GDP per capita)]** | | | | | | | |
| ***Low maternal HIV prevalence/incidence*** | | | | | | | |
| Standard-of-care | 3·6 | 5·0 | 8·4 | 60·177 | 26·392 | 5 | Reference |
| HR-HIVE – 1 dose | 3·3 | 4·9 | 8·0 | 60·178 | 26·393 | 5 | dominated |
| HR-HIVE – 2 doses | 3·3 | 4·7 | 7·8 | 60·179 | 26·393 | 5 | dominated |
| HIVE – 1 dose | 3·3 | 4·7 | 7·8 | 60·179 | 26·393 | 5 | dominated |
| HR-HIVE – Extended | 3·3 | 4·4 | 7·5 | 60·180 | 26·393 | 5 | dominated |
| HIVE – 2 doses | 3·3 | 4·3 | 7·4 | 60·180 | 26·393 | 5 | dominated |
| ALL – 2 doses | 3·0 | 4·2 | 7·0 | 60·181 | 26·394 | 35 | dominated |
| ALL – 1 dose | 3·0 | 4·7 | 7·5 | 60·183 | 26·394 | 20 | dominated |
| HIVE – Extended | 3·3 | 3·2 | 6·3 | 60·183 | 26·395 | 4 | cost-saving*·† |
| ALL – 1 dose plus HIVE - Extended | 3·0 | 3·1 | 6·0 | 60·187 | 26·396 | 19 | 9521 |
| ALL - Extended | 3·0 | 2·7 | 5·6 | 60·190 | 26·396 | 85 | 213 006 |
| ***High maternal HIV prevalence/incidence*** | | | | | | | |
| Standard-of-care | 3·6 | 5·0 | 8·4 | 60·059 | 26·345 | 27 | Reference |
| HR-HIVE – 1 dose | 3·3 | 4·8 | 7·9 | 60·065 | 26·348 | 27 | dominated |
| HR-HIVE – 2 doses | 3·3 | 4·7 | 7·8 | 60·067 | 26·349 | 27 | dominated |
| HIVE – 1 dose | 3·3 | 4·7 | 7·8 | 60·069 | 26·349 | 26 | dominated |
| HR-HIVE – Extended | 3·3 | 4·3 | 7·5 | 60·072 | 26·351 | 27 | dominated |
| HIVE – 2 doses | 3·3 | 4·3 | 7·4 | 60·075 | 26·352 | 25 | dominated |
| ALL – 1 dose | 3·0 | 4·6 | 7·5 | 60·077 | 26·353 | 40 | dominated |
| ALL – 2 doses | 3·0 | 4·1 | 7·0 | 60·081 | 26·355 | 54 | dominated |
| HIVE – Extended | 3·3 | 3·1 | 6·3 | 60·090 | 26·357 | 23 | cost-saving*·† |
| ALL – 1 dose plus HIVE - Extended | 3·0 | 3·1 | 6·0 | 60·098 | 26·361 | 37 | 4158 |
| ALL - Extended | 3·0 | 2·7 | 5·5 | 60·106 | 26·363 | 99 | 32 843 |
| **South Africa [CET: ICER ≤ $1131/YLS (20% GDP per capita), ICER ≤ $2828/YLS (50% GDP per capita)]** | | | | | | | |
| ***Low maternal HIV prevalence/incidence*** | | | | | | | |
| Standard-of-care | 2·5 | 2·5 | 4·8 | 69·322 | 28·639 | 56 | Reference |
| HR-HIVE – 1 dose | 2·2 | 2·3 | 4·4 | 69·349 | 28·648 | 57 | dominated |
| HR-HIVE – 2 doses | 2·2 | 2·3 | 4·4 | 69·352 | 28·649 | 57 | dominated |
| HR-HIVE – Extended | 2·2 | 2·2 | 4·3 | 69·356 | 28·650 | 56 | dominated |
| HIVE – 1 dose | 2·1 | 2·2 | 4·2 | 69·363 | 28·653 | 52 | dominated |
| HIVE – 2 doses | 2·1 | 2·0 | 4·0 | 69·378 | 28·658 | 52 | dominated |
| ALL – 1 dose | 2·0 | 2·0 | 3·8 | 69·389 | 28·661 | 70 | dominated |
| HIVE – Extended | 2·1 | 1·7 | 3·7 | 69·401 | 28·665 | 50 | cost-saving* |
| ALL – 2 doses | 2·0 | 1·6 | 3·4 | 69·423 | 28·673 | 81 | dominated |
| ALL – 1 dose plus HIVE - Extended | 2·0 | 1·5 | 3·3 | 69·427 | 28·674 | 68 | 2065 |
| ALL - Extended | 2·0 | 0·9 | 2·7 | 69·477 | 28·690 | 104 | 2262† |

**Supplementary Table 8.** Scenario analysis: maternal HIV prevalence and incidence (cont.)

| | **Clinical outcomes** | | | **Lifetime efficacy and costs** | | | |
|---|---|---|---|---|---|---|---|
| **Country/strategy** | **IU/IP cumulative HIV incidence (%)** | **Postnatal cumulative HIV incidence (%)** | **Total cumulative HIV incidence (%)** | **Undiscounted life expectancy (yrs)** | **Discounted life expectancy (yrs)** | **Discounted costs ($)** | **ICER ($/YLS)** |
| **South Africa [CET: ICER ≤ $1131/YLS (20% GDP per capita), ICER ≤ $2828/YLS (50% GDP per capita)]** | | | | | | | |
| ***High maternal HIV prevalence/incidence*** | | | | | | | |
| Standard-of-care | 2·2 | 2·0 | 4·1 | 68·714 | 28·426 | 144 | Reference |
| HR-HIVE – 1 dose | 1·9 | 1·9 | 3·7 | 68·786 | 28·450 | 149 | dominated |
| HR-HIVE – 2 doses | 1·9 | 1·9 | 3·7 | 68·795 | 28·453 | 149 | dominated |
| HR-HIVE – Extended | 1·9 | 1·8 | 3·7 | 68·807 | 28·457 | 148 | dominated |
| HIVE – 1 dose | 1·8 | 1·8 | 3·5 | 68·830 | 28·466 | 135 | dominated |
| HIVE – 2 doses | 1·8 | 1·6 | 3·3 | 68·876 | 28·482 | 134 | dominated |
| ALL – 1 dose | 1·7 | 1·6 | 3·2 | 68·888 | 28·487 | 142 | dominated |
| HIVE – Extended | 1·8 | 1·3 | 3·0 | 68·945 | 28·504 | 129 | cost-saving |
| ALL – 2 doses | 1·7 | 1·3 | 2·9 | 68·963 | 28·512 | 145 | dominated |
| ALL – 1 dose plus HIVE - Extended | 1·7 | 1·1 | 2·8 | 69·003 | 28·525 | 135 | 298 |
| ALL - Extended | 1·7 | 0·7 | 2·4 | 69·094 | 28·555 | 152 | 548*,† |
| **Zimbabwe [CET: ICER ≤ $243/YLS (20% GDP per capita), ICER ≤ $607/YLS (50% GDP per capita)]** | | | | | | | |
| ***Low maternal HIV prevalence/incidence*** | | | | | | | |
| Standard-of-care | 4·4 | 5·8 | 9·6 | 68·675 | 28·160 | 29 | Reference |
| HR-HIVE – 1 dose | 3·8 | 5·4 | 8·8 | 68·694 | 28·166 | 29 | dominated |
| HR-HIVE – 2 doses | 3·8 | 5·3 | 8·7 | 68·697 | 28·167 | 29 | dominated |
| HIVE – 1 dose | 3·7 | 5·3 | 8·6 | 68·699 | 28·168 | 27 | dominated |
| HR-HIVE – Extended | 3·8 | 5·0 | 8·4 | 68·703 | 28·169 | 28 | dominated |
| ALL – 1 dose | 3·5 | 5·0 | 8·1 | 68·708 | 28·170 | 43 | dominated |
| HIVE – 2 doses | 3·7 | 4·9 | 8·2 | 68·708 | 28·171 | 27 | dominated |
| ALL – 2 doses | 3·5 | 4·4 | 7·5 | 68·728 | 28·176 | 59 | dominated |
| HIVE – Extended | 3·7 | 3·8 | 7·1 | 68·731 | 28·179 | 25 | cost-saving*,† |
| ALL – 1 dose plus HIVE - Extended | 3·5 | 3·5 | 6·6 | 68·739 | 28·181 | 41 | 7410 |
| ALL - Extended | 3·5 | 2·6 | 5·8 | 68·756 | 28·187 | 109 | 10 687 |
| ***High maternal HIV prevalence/incidence*** | | | | | | | |
| Standard-of-care | 4·2 | 5·4 | 9·1 | 67·742 | 27·829 | 139 | Reference |
| HR-HIVE – 1 dose | 3·6 | 5·1 | 8·3 | 67·829 | 27·860 | 137 | dominated |
| HR-HIVE – 2 doses | 3·6 | 5·0 | 8·2 | 67·843 | 27·865 | 137 | dominated |
| HIVE – 1 dose | 3·5 | 4·9 | 8·1 | 67·857 | 27·870 | 130 | dominated |
| HR-HIVE – Extended | 3·6 | 4·7 | 7·9 | 67·871 | 27·874 | 134 | dominated |
| HIVE – 2 doses | 3·5 | 4·5 | 7·7 | 67·903 | 27·886 | 128 | dominated |
| ALL – 1 dose | 3·3 | 4·7 | 7·7 | 67·903 | 27·886 | 138 | dominated |
| ALL – 2 doses | 3·3 | 4·2 | 7·1 | 67·973 | 27·909 | 148 | dominated |
| HIVE – Extended | 3·5 | 3·4 | 6·6 | 68·016 | 27·922 | 119 | cost-saving* |
| ALL – 1 dose plus HIVE - Extended | 3·3 | 3·2 | 6·2 | 68·062 | 27·938 | 126 | 494† |
| ALL - Extended | 3·3 | 2·4 | 5·5 | 68·134 | 27·963 | 173 | 1894 |

*bNAb: broadly neutralizing antibody; IU/IP: intrauterine/intrapartum; yr: year; ICER: incremental cost-effectiveness ratio; YLS: years of life saved; CET: cost-effectiveness threshold; HR-HIVE: high-risk HIV-exposed infants; HIVE: all HIV-exposed infants; ALL: all live infants at birth.*

The setting-specific maternal HIV prevalence and incidence are specified in Supplementary Table 2 (base case) and Supplementary Table 3 (low/high scenarios). Pediatric HIV incidence is rounded to the nearest tenth of a percent. IU/IP HIV incidence is calculated based on the number of infants exposed to HIV at birth. Postnatal and total HIV incidence is calculated based on the number of infants ever exposed to HIV through 36 months of life. Undiscounted and discounted life expectancies are rounded to the nearest ten thousandth. Costs are rounded to the nearest dollar and are presented in 2020 USD. Discounted values are discounted at 3% per year. ICERs are rounded to the nearest dollar and are calculated using unrounded discounted life expectancy and discounted costs. The cost-effective bNAb strategy was the strategy that offered the greatest increase in overall population life expectancy while still having an ICER less than the cost-effectiveness threshold when compared to the next best performing, non-dominated strategy. * Indicates the cost-effective strategy using a cost-effectiveness threshold of 20% GDP per capita. † Indicates the cost-effective strategy using a cost-effectiveness threshold of 50% GDP per capita.

**Supplementary Table 9.** One-way sensitivity analysis: bNAb efficacy against intrapartum and postnatal transmission (base case: 70% reduction)

| | Clinical outcomes | | | Lifetime efficacy and costs | | | |
|---|---|---|---|---|---|---|---|
| **Country/strategy** | **IU/IP cumulative HIV incidence (%)** | **Postnatal cumulative HIV incidence (%)** | **Total cumulative HIV incidence (%)** | **Undiscounted life expectancy (yrs)** | **Discounted life expectancy (yrs)** | **Discounted costs ($)** | **ICER ($/YLS)** |
| **Côte d'Ivoire [CET: ICER ≤ $465/YLS (20% GDP per capita), ICER ≤ $1163/YLS (50% GDP per capita)]** | | | | | | | |
| ***bNAb efficacy: 10%*** | | | | | | | |
| Standard-of-care | 3·6 | 5·0 | 8·4 | 60·129 | 26·373 | 15 | Reference* |
| HR-HIVE – 1 dose | 3·6 | 5·0 | 8·3 | 60·130 | 26·373 | 15 | dominated |
| HR-HIVE – 2 doses | 3·6 | 4·9 | 8·3 | 60·130 | 26·373 | 15 | dominated |
| HIVE – 1 dose | 3·6 | 4·9 | 8·3 | 60·130 | 26·374 | 15 | 706† |
| HR-HIVE – Extended | 3·6 | 4·9 | 8·2 | 60·130 | 26·374 | 16 | dominated |
| HIVE – 2 doses | 3·6 | 4·9 | 8·2 | 60·131 | 26·374 | 15 | dominated |
| ALL – 2 doses | 3·5 | 4·9 | 8·2 | 60·130 | 26·374 | 45 | dominated |
| HIVE – Extended | 3·6 | 4·7 | 8·1 | 60·132 | 26·374 | 16 | 1593 |
| ALL – 1 dose | 3·5 | 4·9 | 8·2 | 60·133 | 26·375 | 28 | dominated |
| ALL – 1 dose plus HIVE - Extended | 3·5 | 4·7 | 8·0 | 60·134 | 26·375 | 29 | 11 561 |
| ALL - Extended | 3·5 | 4·7 | 8·0 | 60·137 | 26·375 | 95 | 615 409 |
| ***bNAb efficacy: 100%*** | | | | | | | |
| Standard-of-care | 3·6 | 5·0 | 8·4 | 60·129 | 26·373 | 15 | Reference |
| HR-HIVE – 1 dose | 3·2 | 4·8 | 7·8 | 60·134 | 26·375 | 15 | dominated |
| HR-HIVE – 2 doses | 3·2 | 4·6 | 7·6 | 60·136 | 26·376 | 14 | dominated |
| HIVE – 1 dose | 3·1 | 4·6 | 7·5 | 60·137 | 26·376 | 14 | dominated |
| HR-HIVE – Extended | 3·2 | 4·1 | 7·1 | 60·139 | 26·377 | 14 | dominated |
| HIVE – 2 doses | 3·1 | 4·0 | 7·0 | 60·141 | 26·378 | 13 | dominated |
| ALL – 1 dose | 2·8 | 4·5 | 7·1 | 60·143 | 26·379 | 27 | dominated |
| ALL – 2 doses | 2·8 | 3·8 | 6·4 | 60·146 | 26·380 | 42 | dominated |
| HIVE – Extended | 3·1 | 2·4 | 5·3 | 60·153 | 26·382 | 11 | cost-saving*·† |
| ALL – 1 dose plus HIVE - Extended | 2·8 | 2·3 | 4·9 | 60·159 | 26·385 | 24 | 5017 |
| ALL - Extended | 2·8 | 1·6 | 4·3 | 60·166 | 26·387 | 89 | 38 627 |
| **South Africa [CET: ICER ≤ $1131/YLS (20% GDP per capita), ICER ≤ $2828/YLS (50% GDP per capita)]** | | | | | | | |
| ***bNAb efficacy: 10%*** | | | | | | | |
| Standard-of-care | 2·3 | 2·2 | 4·4 | 68·924 | 28·499 | 112 | Reference |
| HR-HIVE – 1 dose | 2·3 | 2·2 | 4·3 | 68·931 | 28·502 | 122 | dominated |
| HR-HIVE – 2 doses | 2·3 | 2·2 | 4·3 | 68·933 | 28·502 | 122 | dominated |
| HR-HIVE – Extended | 2·3 | 2·2 | 4·3 | 68·934 | 28·503 | 122 | dominated |
| HIVE – 1 dose | 2·3 | 2·2 | 4·3 | 68·934 | 28·503 | 115 | 1009 |
| ALL – 1 dose | 2·3 | 2·1 | 4·2 | 68·938 | 28·504 | 132 | dominated |
| HIVE – 2 doses | 2·3 | 2·1 | 4·3 | 68·940 | 28·505 | 118 | dominated |
| HIVE – Extended | 2·3 | 2·1 | 4·2 | 68·947 | 28·507 | 120 | 1118*·† |
| ALL – 2 doses | 2·3 | 2·1 | 4·2 | 68·950 | 28·508 | 143 | dominated |
| ALL – 1 dose plus HIVE - Extended | 2·3 | 2·1 | 4·2 | 68·951 | 28·509 | 136 | dominated |
| ALL - Extended | 2·3 | 2·0 | 4·1 | 68·967 | 28·513 | 169 | 7902 |

**Supplementary Table 9.** One-way sensitivity analysis: bNAb efficacy against intrapartum and postnatal transmission (base case: 70% reduction) (cont.)

| Country/strategy | Clinical outcomes | | | Lifetime efficacy and costs | | | |
|---|---|---|---|---|---|---|---|
| | IU/IP cumulative HIV incidence (%) | Postnatal cumulative HIV incidence (%) | Total cumulative HIV incidence (%) | Undiscounted life expectancy (yrs) | Discounted life expectancy (yrs) | Discounted costs ($) | ICER ($/YLS) |
| **South Africa [CET: ICER ≤ $1131/YLS (20% GDP per capita), ICER ≤ $2828/YLS (50% GDP per capita)]** | | | | | | | |
| ***bNAb efficacy: 100%*** | | | | | | | |
| Standard-of-care | 2·3 | 2·2 | 4·4 | 68·924 | 28·499 | 112 | Reference |
| HR-HIVE – 1 dose | 1·9 | 2·1 | 3·9 | 69·004 | 28·527 | 112 | dominated |
| HR-HIVE – 2 doses | 1·9 | 2·0 | 3·8 | 69·013 | 28·530 | 111 | dominated |
| HR-HIVE – Extended | 1·9 | 1·9 | 3·7 | 69·026 | 28·534 | 110 | dominated |
| HIVE – 1 dose | 1·8 | 1·9 | 3·6 | 69·050 | 28·543 | 100 | dominated |
| HIVE – 2 doses | 1·8 | 1·6 | 3·3 | 69·098 | 28·559 | 97 | dominated |
| ALL – 1 dose | 1·6 | 1·6 | 3·1 | 69·127 | 28·570 | 108 | dominated |
| HIVE – Extended | 1·8 | 1·2 | 2·9 | 69·169 | 28·582 | 90 | cost-saving |
| ALL – 2 doses | 1·6 | 1·1 | 2·6 | 69·210 | 28·599 | 111 | dominated |
| ALL – 1 dose plus HIVE - Extended | 1·6 | 0·9 | 2·4 | 69·246 | 28·609 | 98 | 295 |
| ALL - Extended | 1·6 | 0·2 | 1·7 | 69·363 | 28·648 | 117 | 476*,† |
| **Zimbabwe [CET: ICER ≤ $243/YLS (20% GDP per capita), ICER ≤ $607/YLS (50% GDP per capita)]** | | | | | | | |
| ***bNAb efficacy: 10%*** | | | | | | | |
| Standard-of-care | 4·3 | 5·6 | 9·5 | 68·321 | 28·034 | 72 | Reference* |
| ALL – 1 dose | 4·2 | 5·5 | 9·2 | 68·328 | 28·036 | 87 | dominated |
| HR-HIVE – 1 dose | 4·3 | 5·6 | 9·3 | 68·327 | 28·036 | 75 | dominated |
| HR-HIVE – 2 doses | 4·3 | 5·6 | 9·3 | 68·328 | 28·037 | 75 | dominated |
| HIVE – 1 dose | 4·2 | 5·6 | 9·3 | 68·330 | 28·037 | 72 | dominated |
| HR-HIVE – Extended | 4·3 | 5·5 | 9·3 | 68·330 | 28·037 | 76 | 276† |
| HIVE – 2 doses | 4·2 | 5·5 | 9·3 | 68·333 | 28·038 | 74 | dominated |
| ALL – 2 doses | 4·2 | 5·5 | 9·2 | 68·339 | 28·039 | 104 | dominated |
| ALL – 1 dose plus HIVE - Extended | 4·2 | 5·3 | 9·0 | 68·338 | 28·039 | 92 | 1379 |
| HIVE – Extended | 4·2 | 5·4 | 9·1 | 68·340 | 28·040 | 77 | dominated |
| ALL - Extended | 4·2 | 5·2 | 8·9 | 68·342 | 28·041 | 157 | 82 283 |
| ***bNAb efficacy: 100%*** | | | | | | | |
| Standard-of-care | 4·3 | 5·6 | 9·5 | 68·321 | 28·034 | 72 | Reference |
| HR-HIVE – 1 dose | 3·5 | 5·2 | 8·3 | 68·386 | 28·057 | 68 | dominated |
| HR-HIVE – 2 doses | 3·5 | 5·0 | 8·1 | 68·396 | 28·061 | 67 | dominated |
| HIVE – 1 dose | 3·4 | 4·9 | 7·9 | 68·405 | 28·064 | 64 | dominated |
| HR-HIVE – Extended | 3·5 | 4·6 | 7·7 | 68·417 | 28·067 | 65 | dominated |
| HIVE – 2 doses | 3·4 | 4·4 | 7·4 | 68·437 | 28·075 | 62 | dominated |
| ALL – 1 dose | 3·1 | 4·5 | 7·3 | 68·437 | 28·076 | 75 | dominated |
| ALL – 2 doses | 3·1 | 3·7 | 6·5 | 68·490 | 28·093 | 87 | dominated |
| HIVE – Extended | 3·4 | 2·7 | 5·8 | 68·518 | 28·101 | 53 | cost-saving*,† |
| ALL – 1 dose plus HIVE - Extended | 3·1 | 2·4 | 5·1 | 68·550 | 28·113 | 65 | 995 |
| ALL - Extended | 3·1 | 1·2 | 4·0 | 68·609 | 28·132 | 121 | 2849 |

*bNAb: broadly neutralizing antibody; IU/IP: intrauterine/intrapartum; yr: year; ICER: incremental cost-effectiveness ratio; YLS: years of life saved; CET: cost-effectiveness threshold; HR-HIVE: high-risk HIV-exposed infants; HIVE: all HIV-exposed infants; ALL: all live infants at birth.*

Pediatric HIV incidence is rounded to the nearest tenth of a percent. IU/IP HIV incidence is calculated based on the number of infants exposed to HIV at birth. Postnatal and total HIV incidence is calculated based on the number of infants ever exposed to HIV through 36 months of life. Undiscounted and discounted life expectancies are rounded to the nearest ten thousandth. Costs are rounded to the nearest dollar and are presented in 2020 USD. Discounted values are discounted at 3% per year. ICERs are rounded to the nearest dollar and are calculated using unrounded discounted life expectancy and discounted costs. The cost-effective bNAb strategy was the strategy that offered the greatest increase in overall population life expectancy while still having an ICER less than the cost-effectiveness threshold when compared to the next best performing, non-dominated strategy. * Indicates the cost-effective strategy using a cost-effectiveness threshold of 20% GDP per capita. † Indicates the cost-effective strategy using a cost-effectiveness threshold of 50% GDP per capita.

**Supplementary Table 10.** One-way sensitivity analysis: bNAb cost (base case: $20/dose)

| | Clinical outcomes | | | Lifetime efficacy and costs | | | |
|---|---|---|---|---|---|---|---|
| **Country/strategy** | **IU/IP cumulative HIV incidence (%)** | **Postnatal cumulative HIV incidence (%)** | **Total cumulative HIV incidence (%)** | **Undiscounted life expectancy (yrs)** | **Discounted life expectancy (yrs)** | **Discounted costs ($)** | **ICER ($/YLS)** |
| **Côte d'Ivoire [CET: ICER ≤ $465/YLS (20% GDP per capita), ICER ≤ $1163/YLS (50% GDP per capita)]** | | | | | | | |
| ***bNAb cost: $5/dose*** | | | | | | | |
| Standard-of-care | 3·6 | 5·0 | 8·4 | 60·129 | 26·373 | 15 | Reference |
| HR-HIVE – 1 dose | 3·3 | 4·8 | 8·0 | 60·133 | 26·375 | 15 | dominated |
| HR-HIVE – 2 doses | 3·3 | 4·7 | 7·8 | 60·134 | 26·375 | 15 | dominated |
| HIVE – 1 dose | 3·3 | 4·7 | 7·8 | 60·135 | 26·375 | 14 | dominated |
| HR-HIVE – Extended | 3·3 | 4·3 | 7·5 | 60·136 | 26·376 | 14 | dominated |
| HIVE – 2 doses | 3·3 | 4·3 | 7·4 | 60·138 | 26·377 | 13 | dominated |
| ALL – 1 dose | 3·0 | 4·6 | 7·5 | 60·139 | 26·377 | 17 | dominated |
| ALL – 2 doses | 3·0 | 4·2 | 7·0 | 60·140 | 26·378 | 21 | dominated |
| HIVE – Extended | 3·3 | 3·2 | 6·3 | 60·146 | 26·379 | 11 | cost-saving*,† |
| ALL – 1 dose plus HIVE - Extended | 3·0 | 3·1 | 6·0 | 60·151 | 26·382 | 15 | 1673 |
| ALL - Extended | 3·0 | 2·7 | 5·5 | 60·156 | 26·383 | 31 | 14 322 |
| ***bNAb cost: $100/dose*** | | | | | | | |
| Standard-of-care | 3·6 | 5·0 | 8·4 | 60·129 | 26·373 | 15 | Reference |
| HR-HIVE – 1 dose | 3·3 | 4·8 | 8·0 | 60·133 | 26·375 | 15 | dominated |
| HR-HIVE – 2 doses | 3·3 | 4·7 | 7·8 | 60·134 | 26·375 | 16 | dominated |
| HIVE – 1 dose | 3·3 | 4·7 | 7·8 | 60·135 | 26·375 | 15 | 400* |
| HR-HIVE – Extended | 3·3 | 4·3 | 7·5 | 60·136 | 26·376 | 16 | dominated |
| HIVE – 2 doses | 3·3 | 4·3 | 7·4 | 60·138 | 26·377 | 17 | dominated |
| ALL – 1 dose | 3·0 | 4·6 | 7·5 | 60·139 | 26·377 | 82 | dominated |
| ALL – 2 doses | 3·0 | 4·2 | 7·0 | 60·140 | 26·378 | 162 | dominated |
| HIVE – Extended | 3·3 | 3·2 | 6·3 | 60·146 | 26·379 | 19 | 874† |
| ALL – 1 dose plus HIVE - Extended | 3·0 | 3·1 | 6·0 | 60·151 | 26·382 | 86 | 31 377 |
| ALL - Extended | 3·0 | 2·7 | 5·5 | 60·156 | 26·383 | 410 | 289 474 |
| **South Africa [CET: ICER ≤ $1131/YLS (20% GDP per capita), ICER ≤ $2828/YLS (50% GDP per capita)]** | | | | | | | |
| ***bNAb cost: $5/dose*** | | | | | | | |
| Standard-of-care | 2·3 | 2·2 | 4·4 | 68·924 | 28·499 | 112 | Reference |
| HR-HIVE – 1 dose | 2·1 | 2·1 | 4·0 | 68·979 | 28·518 | 114 | dominated |
| HR-HIVE – 2 doses | 2·1 | 2·1 | 4·0 | 68·986 | 28·521 | 114 | dominated |
| HR-HIVE – Extended | 2·1 | 2·0 | 3·9 | 68·995 | 28·523 | 113 | dominated |
| HIVE – 1 dose | 2·0 | 2·0 | 3·8 | 69·011 | 28·529 | 103 | dominated |
| HIVE – 2 doses | 2·0 | 1·8 | 3·6 | 69·045 | 28·541 | 99 | dominated |
| ALL – 1 dose | 1·8 | 1·8 | 3·5 | 69·063 | 28·548 | 105 | dominated |
| HIVE – Extended | 2·0 | 1·5 | 3·3 | 69·095 | 28·557 | 93 | cost-saving |
| ALL – 2 doses | 1·8 | 1·5 | 3·2 | 69·122 | 28·568 | 102 | dominated |
| ALL – 1 dose plus HIVE - Extended | 1·8 | 1·3 | 3·0 | 69·147 | 28·575 | 96 | dominated |
| ALL - Extended | 1·8 | 0·8 | 2·5 | 69·228 | 28·602 | 94 | 24*,† |

**Supplementary Table 10.** One-way sensitivity analysis: bNAb cost (base case: $20/dose) (cont.)

| Country/strategy | Clinical outcomes | | | Lifetime efficacy and costs | | | |
|---|---|---|---|---|---|---|---|
| | IU/IP cumulative HIV incidence (%) | Postnatal cumulative HIV incidence (%) | Total cumulative HIV incidence (%) | Undiscounted life expectancy (yrs) | Discounted life expectancy (yrs) | Discounted costs ($) | ICER ($/YLS) |
| **South Africa [CET: ICER ≤ $1131/YLS (20% GDP per capita), ICER ≤ $2828/YLS (50% GDP per capita)]** | | | | | | | |
| ***bNAb cost: $100/dose*** | | | | | | | |
| Standard-of-care | 2·3 | 2·2 | 4·4 | 68·924 | 28·499 | 112 | Reference |
| HR-HIVE – 1 dose | 2·1 | 2·1 | 4·0 | 68·979 | 28·518 | 117 | dominated |
| HR-HIVE – 2 doses | 2·1 | 2·1 | 4·0 | 68·986 | 28·521 | 119 | dominated |
| HR-HIVE – Extended | 2·1 | 2·0 | 3·9 | 68·995 | 28·523 | 121 | dominated |
| HIVE – 1 dose | 2·0 | 2·0 | 3·8 | 69·011 | 28·529 | 118 | 214 |
| HIVE – 2 doses | 2·0 | 1·8 | 3·6 | 69·045 | 28·541 | 127 | dominated |
| ALL – 1 dose | 1·8 | 1·8 | 3·5 | 69·063 | 28·548 | 172 | dominated |
| HIVE – Extended | 2·0 | 1·5 | 3·3 | 69·095 | 28·557 | 138 | 723*,† |
| ALL – 2 doses | 1·8 | 1·5 | 3·2 | 69·122 | 28·568 | 228 | dominated |
| ALL – 1 dose plus HIVE - Extended | 1·8 | 1·3 | 3·0 | 69·147 | 28·575 | 192 | 2918 |
| ALL - Extended | 1·8 | 0·8 | 2·5 | 69·228 | 28·602 | 350 | 5977 |
| **Zimbabwe [CET: ICER ≤ $243/YLS (20% GDP per capita), ICER ≤ $607/YLS (50% GDP per capita)]** | | | | | | | |
| ***bNAb cost: $5/dose*** | | | | | | | |
| Standard-of-care | 4·3 | 5·6 | 9·5 | 68·321 | 28·034 | 72 | Reference |
| HR-HIVE – 1 dose | 3·7 | 5·3 | 8·6 | 68·366 | 28·050 | 70 | dominated |
| HR-HIVE – 2 doses | 3·7 | 5·2 | 8·5 | 68·373 | 28·053 | 69 | dominated |
| HIVE – 1 dose | 3·7 | 5·2 | 8·4 | 68·380 | 28·055 | 65 | dominated |
| HR-HIVE – Extended | 3·7 | 4·9 | 8·2 | 68·387 | 28·057 | 67 | dominated |
| ALL – 1 dose | 3·4 | 4·9 | 7·9 | 68·400 | 28·062 | 67 | dominated |
| HIVE – 2 doses | 3·7 | 4·8 | 8·0 | 68·402 | 28·063 | 63 | dominated |
| ALL – 2 doses | 3·4 | 4·3 | 7·4 | 68·438 | 28·074 | 67 | dominated |
| HIVE – Extended | 3·7 | 3·6 | 6·9 | 68·458 | 28·081 | 55 | cost-saving |
| ALL – 1 dose plus HIVE - Extended | 3·4 | 3·4 | 6·5 | 68·478 | 28·088 | 56 | 160*,† |
| ALL - Extended | 3·4 | 2·6 | 5·7 | 68·517 | 28·101 | 67 | 791 |
| ***bNAb cost: $100/dose*** | | | | | | | |
| Standard-of-care | 4·3 | 5·6 | 9·5 | 68·321 | 28·034 | 72 | Reference |
| HR-HIVE – 1 dose | 3·7 | 5·3 | 8·6 | 68·366 | 28·050 | 72 | 54* |
| HR-HIVE – 2 doses | 3·7 | 5·2 | 8·5 | 68·373 | 28·053 | 74 | dominated |
| HIVE – 1 dose | 3·7 | 5·2 | 8·4 | 68·380 | 28·055 | 74 | 295† |
| HR-HIVE – Extended | 3·7 | 4·9 | 8·2 | 68·387 | 28·057 | 78 | dominated |
| ALL – 1 dose | 3·4 | 4·9 | 7·9 | 68·400 | 28·062 | 146 | dominated |
| HIVE – 2 doses | 3·7 | 4·8 | 8·0 | 68·402 | 28·063 | 80 | dominated |
| ALL – 2 doses | 3·4 | 4·3 | 7·4 | 68·438 | 28·074 | 228 | dominated |
| HIVE – Extended | 3·7 | 3·6 | 6·9 | 68·458 | 28·081 | 93 | 756 |
| ALL – 1 dose plus HIVE - Extended | 3·4 | 3·4 | 6·5 | 68·478 | 28·088 | 165 | 9833 |
| ALL - Extended | 3·4 | 2·6 | 5·7 | 68·517 | 28·101 | 488 | 24 338 |

*bNAb: broadly neutralizing antibody; IU/IP: intrauterine/intrapartum; yr: year; ICER: incremental cost-effectiveness ratio; YLS: years of life saved; CET: cost-effectiveness threshold; HR-HIVE: high-risk HIV-exposed infants; HIVE: all HIV-exposed infants; ALL: all live infants at birth.*

Pediatric HIV incidence is rounded to the nearest tenth of a percent. IU/IP HIV incidence is calculated based on the number of infants exposed to HIV at birth. Postnatal and total HIV incidence is calculated based on the number of infants ever exposed to HIV through 36 months of life. Undiscounted and discounted life expectancies are rounded to the nearest ten thousandth. Costs are rounded to the nearest dollar and are presented in 2020 USD. Discounted values are discounted at 3% per year. ICERs are rounded to the nearest dollar and are calculated using unrounded discounted life expectancy and discounted costs. The cost-effective bNAb strategy was the strategy that offered the greatest increase in overall population life expectancy while still having an ICER less than the cost-effectiveness threshold when compared to the next best performing, non-dominated strategy. * Indicates the cost-effective strategy using a cost-effectiveness threshold of 20% GDP per capita. † Indicates the cost-effective strategy using a cost-effectiveness threshold of 50% GDP per capita.

**Supplementary Table 11.** One-way sensitivity analysis: bNAb effect duration (base case: 3 months)

| Country/strategy | Clinical outcomes | | | Lifetime efficacy and costs | | | |
|---|---|---|---|---|---|---|---|
| | **IU/IP cumulative HIV incidence (%)** | **Postnatal cumulative HIV incidence (%)** | **Total cumulative HIV incidence (%)** | **Undiscounted life expectancy (yrs)** | **Discounted life expectancy (yrs)** | **Discounted costs ($)** | **ICER ($/YLS)** |
| **Côte d'Ivoire [CET: ICER ≤ $465/YLS (20% GDP per capita), ICER ≤ $1163/YLS (50% GDP per capita)]** | | | | | | | |
| ***bNAb effect duration: 1 month*** | | | | | | | |
| Standard-of-care | 3·6 | 5·0 | 8·4 | 60·129 | 26·373 | 15 | Reference |
| HR-HIVE – 1 dose | 3·3 | 4·9 | 8·0 | 60·132 | 26·374 | 15 | dominated |
| HR-HIVE – 2 doses | 3·3 | 4·9 | 8·0 | 60·133 | 26·375 | 15 | dominated |
| HIVE – 1 dose | 3·3 | 4·9 | 7·9 | 60·133 | 26·375 | 14 | cost-saving |
| HIVE – 2 doses | 3·3 | 4·8 | 7·8 | 60·134 | 26·375 | 15 | dominated |
| HR-HIVE – Extended | 3·3 | 4·4 | 7·5 | 60·136 | 26·376 | 15 | dominated |
| ALL – 1 dose | 3·0 | 4·9 | 7·7 | 60·137 | 26·377 | 28 | dominated |
| ALL – 2 doses | 3·0 | 4·7 | 7·6 | 60·140 | 26·378 | 45 | dominated |
| HIVE – Extended | 3·3 | 3·2 | 6·3 | 60·145 | 26·379 | 16 | 267*,† |
| ALL – 1 dose plus HIVE - Extended | 3·0 | 3·2 | 6·1 | 60·150 | 26·381 | 29 | 7052 |
| ALL - Extended | 3·0 | 2·7 | 5·6 | 60·158 | 26·384 | 235 | 81 484 |
| ***bNAb effect duration: 6 months*** | | | | | | | |
| Standard-of-care | 3·6 | 5·0 | 8·4 | 60·129 | 26·373 | 15 | Reference |
| HR-HIVE – 1 dose | 3·3 | 4·7 | 7·8 | 60·134 | 26·375 | 15 | dominated |
| HR-HIVE – 2 doses | 3·3 | 4·5 | 7·6 | 60·135 | 26·376 | 14 | dominated |
| HR-HIVE – Extended | 3·3 | 4·3 | 7·5 | 60·136 | 26·376 | 14 | dominated |
| HIVE – 1 dose | 3·3 | 4·4 | 7·4 | 60·137 | 26·376 | 14 | dominated |
| HIVE – 2 doses | 3·3 | 3·7 | 6·7 | 60·142 | 26·378 | 13 | dominated |
| ALL – 1 dose | 3·0 | 4·2 | 7·1 | 60·143 | 26·379 | 27 | dominated |
| HIVE – Extended | 3·3 | 3·1 | 6·2 | 60·146 | 26·379 | 12 | cost-saving*,† |
| ALL – 2 doses | 3·0 | 3·3 | 6·2 | 60·145 | 26·381 | 40 | dominated |
| ALL – 1 dose plus HIVE - Extended | 3·0 | 3·0 | 5·9 | 60·151 | 26·382 | 25 | 5467 |
| ALL - Extended | 3·0 | 2·6 | 5·5 | 60·158 | 26·383 | 55 | 19 636 |
| **South Africa [CET: ICER ≤ $1131/YLS (20% GDP per capita), ICER ≤ $2828/YLS (50% GDP per capita)]** | | | | | | | |
| ***bNAb effect duration: 1 month*** | | | | | | | |
| Standard-of-care | 2·3 | 2·2 | 4·4 | 68·924 | 28·499 | 112 | Reference |
| HR-HIVE – 1 dose | 2·1 | 2·1 | 4·1 | 68·970 | 28·515 | 116 | dominated |
| HR-HIVE – 2 doses | 2·1 | 2·1 | 4·0 | 68·976 | 28·517 | 116 | dominated |
| HIVE – 1 dose | 2·0 | 2·1 | 4·0 | 68·985 | 28·520 | 108 | cost-saving |
| HR-HIVE – Extended | 2·1 | 2·0 | 3·9 | 68·995 | 28·523 | 117 | dominated |
| HIVE – 2 doses | 2·0 | 2·1 | 3·9 | 68·997 | 28·525 | 110 | dominated |
| ALL – 1 dose | 1·8 | 2·1 | 3·8 | 69·011 | 28·529 | 121 | dominated |
| ALL – 2 doses | 1·8 | 1·9 | 3·6 | 69·038 | 28·539 | 133 | dominated |
| HIVE – Extended | 2·0 | 1·5 | 3·3 | 69·093 | 28·557 | 116 | 222* |
| ALL – 1 dose plus HIVE - Extended | 1·8 | 1·4 | 3·1 | 69·119 | 28·566 | 130 | 1462† |
| ALL - Extended | 1·8 | 0·8 | 2·5 | 69·221 | 28·600 | 227 | 2861 |

**Supplementary Table 11.** One-way sensitivity analysis: bNAb effect duration (base case: 3 months) (cont.)

| Country/strategy | Clinical outcomes: IU/IP cumulative HIV incidence (%) | Postnatal cumulative HIV incidence (%) | Total cumulative HIV incidence (%) | Lifetime efficacy and costs: Undiscounted life expectancy (yrs) | Discounted life expectancy (yrs) | Discounted costs ($) | ICER ($/YLS) |
|---|---|---|---|---|---|---|---|
| **South Africa [CET: ICER ≤ $1131/YLS (20% GDP per capita), ICER ≤ $2828/YLS (50% GDP per capita)]** | | | | | | | |
| ***bNAb effect duration: 6 months*** | | | | | | | |
| Standard-of-care | 2·3 | 2·2 | 4·4 | 68·924 | 28·499 | 112 | Reference |
| HR-HIVE – 1 dose | 2·1 | 2·1 | 4·0 | 68·986 | 28·520 | 114 | dominated |
| HR-HIVE – 2 doses | 2·1 | 2·0 | 3·9 | 68·994 | 28·523 | 113 | dominated |
| HR-HIVE – Extended | 2·1 | 2·0 | 3·9 | 68·995 | 28·523 | 113 | dominated |
| HIVE – 1 dose | 2·0 | 1·8 | 3·6 | 69·044 | 28·541 | 101 | dominated |
| HIVE – 2 doses | 2·0 | 1·5 | 3·4 | 69·086 | 28·554 | 97 | dominated |
| HIVE – Extended | 2·0 | 1·5 | 3·3 | 69·097 | 28·558 | 96 | cost-saving |
| ALL – 1 dose | 1·8 | 1·5 | 3·2 | 69·119 | 28·567 | 109 | dominated |
| ALL – 1 dose plus HIVE - Extended | 1·8 | 1·1 | 2·8 | 69·171 | 28·584 | 105 | dominated |
| ALL – 2 doses | 1·8 | 1·0 | 2·7 | 69·200 | 28·593 | 110 | dominated |
| ALL - Extended | 1·8 | 0·8 | 2·5 | 69·231 | 28·603 | 111 | 322*·† |
| **Zimbabwe [CET: ICER ≤ $243/YLS (20% GDP per capita), ICER ≤ $607/YLS (50% GDP per capita)]** | | | | | | | |
| ***bNAb effect duration: 1 month*** | | | | | | | |
| Standard-of-care | 4·3 | 5·6 | 9·5 | 68·321 | 28·034 | 72 | Reference |
| HR-HIVE – 1 dose | 3·7 | 5·5 | 8·8 | 68·356 | 28·046 | 72 | dominated |
| HR-HIVE – 2 doses | 3·7 | 5·4 | 8·7 | 68·361 | 28·048 | 72 | dominated |
| HIVE – 1 dose | 3·7 | 5·5 | 8·7 | 68·361 | 28·048 | 69 | cost-saving |
| ALL – 1 dose | 3·4 | 5·4 | 8·4 | 68·370 | 28·051 | 82 | dominated |
| HIVE – 2 doses | 3·7 | 5·3 | 8·6 | 68·370 | 28·051 | 70 | dominated |
| HR-HIVE – Extended | 3·7 | 4·9 | 8·2 | 68·388 | 28·057 | 73 | dominated |
| ALL – 2 doses | 3·4 | 5·1 | 8·2 | 68·392 | 28·058 | 99 | dominated |
| HIVE – Extended | 3·7 | 3·6 | 6·9 | 68·457 | 28·080 | 76 | 223*·† |
| ALL – 1 dose plus HIVE - Extended | 3·4 | 3·6 | 6·7 | 68·466 | 28·083 | 89 | 4366 |
| ALL - Extended | 3·4 | 2·6 | 5·7 | 68·511 | 28·101 | 294 | 11 905 |
| ***bNAb effect duration: 6 months*** | | | | | | | |
| Standard-of-care | 4·3 | 5·6 | 9·5 | 68·321 | 28·034 | 72 | Reference |
| HR-HIVE – 1 dose | 3·7 | 5·2 | 8·5 | 68·373 | 28·052 | 70 | dominated |
| HR-HIVE – 2 doses | 3·7 | 5·0 | 8·3 | 68·382 | 28·055 | 69 | dominated |
| HR-HIVE – Extended | 3·7 | 4·9 | 8·2 | 68·387 | 28·057 | 68 | dominated |
| HIVE – 1 dose | 3·7 | 4·8 | 8·0 | 68·401 | 28·062 | 64 | dominated |
| ALL – 1 dose | 3·4 | 4·4 | 7·4 | 68·430 | 28·072 | 76 | dominated |
| HIVE – 2 doses | 3·7 | 4·1 | 7·4 | 68·436 | 28·074 | 60 | dominated |
| HIVE – Extended | 3·7 | 3·6 | 6·9 | 68·459 | 28·081 | 58 | cost-saving*·† |
| ALL – 2 doses | 3·4 | 3·3 | 6·4 | 68·487 | 28·090 | 83 | dominated |
| ALL – 1 dose plus HIVE - Extended | 3·4 | 3·2 | 6·3 | 68·488 | 28·091 | 69 | 1108 |
| ALL - Extended | 3·4 | 2·5 | 5·6 | 68·520 | 28·102 | 94 | 2223 |

*bNAb: broadly neutralizing antibody; IU/IP: intrauterine/intrapartum; yr: year; ICER: incremental cost-effectiveness ratio; YLS: years of life saved; CET: cost-effectiveness threshold; HR-HIVE: high-risk HIV-exposed infants; HIVE: all HIV-exposed infants; ALL: all live infants at birth.*

Pediatric HIV incidence is rounded to the nearest tenth of a percent. IU/IP HIV incidence is calculated based on the number of infants exposed to HIV at birth. Postnatal and total HIV incidence is calculated based on the number of infants ever exposed to HIV through 36 months of life. Undiscounted and discounted life expectancies are rounded to the nearest ten thousandth. Costs are rounded to the nearest dollar and are presented in 2020 USD. Discounted values are discounted at 3% per year. ICERs are rounded to the nearest dollar and are calculated using unrounded discounted life expectancy and discounted costs. The cost-effective bNAb strategy was the strategy that offered the greatest increase in overall population life expectancy while still having an ICER less than the cost-effectiveness threshold when compared to the next best performing, non-dominated strategy. * Indicates the cost-effective strategy using a cost-effectiveness threshold of 20% GDP per capita. † Indicates the cost-effective strategy using a cost-effectiveness threshold of 50% GDP per capita.

**Supplementary Table 12.** One-way sensitivity analysis: bNAb uptake

| | Clinical outcomes | | | Lifetime efficacy and costs | | | |
|---|---|---|---|---|---|---|---|
| Country/strategy | IU/IP cumulative HIV incidence (%) | Postnatal cumulative HIV incidence (%) | Total cumulative HIV incidence (%) | Undiscounted life expectancy (yrs) | Discounted life expectancy (yrs) | Discounted costs ($) | ICER ($/YLS) |
| **Côte d'Ivoire [CET: ICER ≤ $465/YLS (20% GDP per capita), ICER ≤ $1163/YLS (50% GDP per capita)]** | | | | | | | |
| ***56% uptake at all time points*** | | | | | | | |
| Standard-of-care | 3·6 | 5·0 | 8·4 | 60·129 | 26·373 | 15 | Reference |
| HR-HIVE – 1 dose | 3·4 | 4·9 | 8·0 | 60·132 | 26·374 | 15 | dominated |
| HR-HIVE – 2 doses | 3·4 | 4·8 | 7·9 | 60·133 | 26·375 | 15 | dominated |
| HIVE – 1 dose | 3·3 | 4·8 | 7·9 | 60·133 | 26·375 | 14 | dominated |
| HR-HIVE – Extended | 3·4 | 4·5 | 7·7 | 60·135 | 26·375 | 15 | dominated |
| HIVE – 2 doses | 3·3 | 4·5 | 7·6 | 60·136 | 26·376 | 14 | dominated |
| ALL – 1 dose | 3·1 | 4·7 | 7·7 | 60·138 | 26·376 | 25 | dominated |
| ALL – 2 doses | 3·1 | 4·4 | 7·3 | 60·142 | 26·378 | 35 | dominated |
| HIVE – Extended | 3·3 | 3·6 | 6·7 | 60·142 | 26·378 | 13 | cost-saving*·† |
| ALL – 1 dose plus HIVE - Extended | 3·1 | 3·5 | 6·5 | 60·146 | 26·380 | 24 | 6745 |
| ALL - Extended | 3·1 | 3·2 | 6·2 | 60·153 | 26·381 | 75 | 38 825 |
| ***83% uptake at all time points*** | | | | | | | |
| Standard-of-care | 3·6 | 5·0 | 8·4 | 60·129 | 26·373 | 15 | Reference |
| HR-HIVE – 1 dose | 3·3 | 4·8 | 7·9 | 60·134 | 26·375 | 15 | dominated |
| HR-HIVE – 2 doses | 3·3 | 4·7 | 7·7 | 60·135 | 26·375 | 15 | dominated |
| HIVE – 1 dose | 3·2 | 4·7 | 7·7 | 60·136 | 26·376 | 14 | dominated |
| HR-HIVE – Extended | 3·3 | 4·2 | 7·3 | 60·137 | 26·376 | 14 | dominated |
| HIVE – 2 doses | 3·2 | 4·3 | 7·3 | 60·139 | 26·377 | 14 | dominated |
| ALL – 1 dose | 2·9 | 4·6 | 7·3 | 60·141 | 26·378 | 30 | dominated |
| ALL – 2 doses | 2·9 | 4·1 | 6·8 | 60·143 | 26·379 | 45 | dominated |
| HIVE – Extended | 3·2 | 2·9 | 5·9 | 60·148 | 26·380 | 12 | cost-saving*·† |
| ALL – 1 dose plus HIVE - Extended | 2·9 | 2·8 | 5·6 | 60·154 | 26·383 | 28 | 6494 |
| ALL - Extended | 2·9 | 2·3 | 5·1 | 60·158 | 26·384 | 103 | 65 508 |
| **South Africa [CET: ICER ≤ $1131/YLS (20% GDP per capita), ICER ≤ $2828/YLS (50% GDP per capita)]** | | | | | | | |
| ***85% uptake at all time points*** | | | | | | | |
| Standard-of-care | 2·3 | 2·2 | 4·4 | 68·924 | 28·499 | 112 | Reference |
| HR-HIVE – 1 dose | 2·1 | 2·1 | 4·1 | 68·973 | 28·516 | 116 | dominated |
| HR-HIVE – 2 doses | 2·1 | 2·1 | 4·0 | 68·980 | 28·518 | 115 | dominated |
| HR-HIVE – Extended | 2·1 | 2·0 | 4·0 | 68·987 | 28·521 | 115 | dominated |
| HIVE – 1 dose | 2·0 | 2·0 | 3·9 | 69·002 | 28·526 | 106 | dominated |
| HIVE – 2 doses | 2·0 | 1·8 | 3·7 | 69·032 | 28·537 | 105 | dominated |
| ALL – 1 dose | 1·9 | 1·8 | 3·6 | 69·050 | 28·543 | 115 | dominated |
| HIVE – Extended | 2·0 | 1·5 | 3·4 | 69·078 | 28·552 | 102 | cost-saving |
| ALL – 2 doses | 1·9 | 1·5 | 3·3 | 69·098 | 28·560 | 122 | dominated |
| ALL – 1 dose plus HIVE - Extended | 1·9 | 1·4 | 3·1 | 69·126 | 28·568 | 111 | 589 |
| ALL - Extended | 1·9 | 0·9 | 2·7 | 69·201 | 28·592 | 134 | 930*·† |

**Supplementary Table 12.** One-way sensitivity analysis: bNAb uptake (cont.)

| | Clinical outcomes | | | Lifetime efficacy and costs | | | |
|---|---|---|---|---|---|---|---|
| Country/strategy | IU/IP cumulative HIV incidence (%) | Postnatal cumulative HIV incidence (%) | Total cumulative HIV incidence (%) | Undiscounted life expectancy (yrs) | Discounted life expectancy (yrs) | Discounted costs ($) | ICER ($/YLS) |
| **South Africa [CET: ICER ≤ $1131/YLS (20% GDP per capita), ICER ≤ $2828/YLS (50% GDP per capita)]** | | | | | | | |
| ***96% uptake at all time points*** | | | | | | | |
| Standard-of-care | 2·3 | 2·2 | 4·4 | 68·924 | 28·499 | 112 | Reference |
| HR-HIVE – 1 dose | 2·1 | 2·1 | 4·0 | 68·979 | 28·518 | 115 | dominated |
| HR-HIVE – 2 doses | 2·1 | 2·1 | 4·0 | 68·986 | 28·521 | 115 | dominated |
| HR-HIVE – Extended | 2·1 | 2·0 | 3·9 | 68·996 | 28·524 | 114 | dominated |
| HIVE – 1 dose | 2·0 | 2·0 | 3·8 | 69·011 | 28·529 | 105 | dominated |
| HIVE – 2 doses | 2·0 | 1·8 | 3·6 | 69·045 | 28·541 | 104 | dominated |
| ALL – 1 dose | 1·8 | 1·8 | 3·5 | 69·063 | 28·548 | 116 | dominated |
| HIVE – Extended | 2·0 | 1·5 | 3·3 | 69·098 | 28·558 | 100 | cost-saving |
| ALL – 2 doses | 1·8 | 1·4 | 3·1 | 69·122 | 28·568 | 122 | dominated |
| ALL – 1 dose plus HIVE - Extended | 1·8 | 1·3 | 3·0 | 69·150 | 28·576 | 111 | 606 |
| ALL - Extended | 1·8 | 0·8 | 2·5 | 69·235 | 28·604 | 136 | 891*·† |
| **Zimbabwe [CET: ICER ≤ $243/YLS (20% GDP per capita), ICER ≤ $607/YLS (50% GDP per capita)]** | | | | | | | |
| ***71% uptake at all time points*** | | | | | | | |
| Standard-of-care | 4·3 | 5·6 | 9·5 | 68·321 | 28·034 | 72 | Reference |
| HR-HIVE – 1 dose | 3·8 | 5·4 | 8·8 | 68·358 | 28·047 | 71 | dominated |
| HR-HIVE – 2 doses | 3·8 | 5·3 | 8·7 | 68·363 | 28·049 | 71 | dominated |
| HIVE – 1 dose | 3·8 | 5·2 | 8·6 | 68·370 | 28·051 | 68 | dominated |
| HR-HIVE – Extended | 3·8 | 5·0 | 8·4 | 68·375 | 28·053 | 70 | dominated |
| ALL – 1 dose | 3·6 | 5·0 | 8·2 | 68·381 | 28·056 | 78 | dominated |
| HIVE – 2 doses | 3·8 | 4·9 | 8·3 | 68·387 | 28·057 | 67 | dominated |
| ALL – 2 doses | 3·6 | 4·6 | 7·8 | 68·419 | 28·067 | 89 | dominated |
| HIVE – Extended | 3·8 | 4·0 | 7·3 | 68·433 | 28·072 | 63 | cost-saving*·† |
| ALL – 1 dose plus HIVE - Extended | 3·6 | 3·7 | 7·0 | 68·445 | 28·077 | 73 | 2444 |
| ALL - Extended | 3·6 | 3·1 | 6·3 | 68·488 | 28·090 | 124 | 3706 |
| ***92% uptake at all time points*** | | | | | | | |
| Standard-of-care | 4·3 | 5·6 | 9·5 | 68·321 | 28·034 | 72 | Reference |
| HR-HIVE – 1 dose | 3·7 | 5·3 | 8·6 | 68·369 | 28·051 | 70 | dominated |
| HR-HIVE – 2 doses | 3·7 | 5·2 | 8·4 | 68·377 | 28·054 | 70 | dominated |
| HIVE – 1 dose | 3·6 | 5·1 | 8·3 | 68·384 | 28·056 | 66 | dominated |
| HR-HIVE – Extended | 3·7 | 4·8 | 8·1 | 68·392 | 28·059 | 68 | dominated |
| HIVE – 2 doses | 3·6 | 4·7 | 7·9 | 68·406 | 28·064 | 65 | dominated |
| ALL – 1 dose | 3·4 | 4·8 | 7·8 | 68·409 | 28·065 | 80 | dominated |
| ALL – 2 doses | 3·4 | 4·3 | 7·3 | 68·441 | 28·076 | 93 | dominated |
| HIVE – Extended | 3·6 | 3·4 | 6·7 | 68·469 | 28·084 | 60 | cost-saving*·† |
| ALL – 1 dose plus HIVE - Extended | 3·4 | 3·2 | 6·2 | 68·494 | 28·093 | 74 | 1545 |
| ALL - Extended | 3·4 | 2·3 | 5·3 | 68·535 | 28·107 | 140 | 4822 |

*bNAb: broadly neutralizing antibody; IU/IP: intrauterine/intrapartum; yr: year; ICER: incremental cost-effectiveness ratio; YLS: years of life saved; CET: cost-effectiveness threshold; HR-HIVE: high-risk HIV-exposed infants; HIVE: all HIV-exposed infants; ALL: all live infants at birth.*

The setting-specific base case uptake is specified in Supplementary Table 2. Pediatric HIV incidence is rounded to the nearest tenth of a percent. IU/IP HIV incidence is calculated based on the number of infants exposed to HIV at birth. Postnatal and total HIV incidence is calculated based on the number of infants ever exposed to HIV through 36 months of life. Undiscounted and discounted life expectancies are rounded to the nearest ten thousandth. Costs are rounded to the nearest dollar and are presented in 2020 USD. Discounted values are discounted at 3% per year. ICERs are rounded to the nearest dollar and are calculated using unrounded discounted life expectancy and discounted costs. The cost-effective bNAb strategy was the strategy that offered the greatest increase in overall population life expectancy while still having an ICER less than the cost-effectiveness threshold when compared to the next best performing, non-dominated strategy. * Indicates the cost-effective strategy using a cost-effectiveness threshold of 20% GDP per capita. † Indicates the cost-effective strategy using a cost-effectiveness threshold of 50% GDP per capita.

**Supplementary Table 13.** One-way sensitivity analysis: proportion of mothers on antiretroviral therapy during pregnancy

| | Clinical outcomes | | | Lifetime efficacy and costs | | | |
|---|---|---|---|---|---|---|---|
| **Country/strategy** | **IU/IP cumulative HIV incidence (%)** | **Postnatal cumulative HIV incidence (%)** | **Total cumulative HIV incidence (%)** | **Undiscounted life expectancy (yrs)** | **Discounted life expectancy (yrs)** | **Discounted costs ($)** | **ICER ($/YLS)** |
| **Côte d'Ivoire [CET: ICER ≤ $465/YLS (20% GDP per capita), ICER ≤ $1163/YLS (50% GDP per capita)]** | | | | | | | |
| ***Proportion of mothers on ART during pregnancy: 70%*** | | | | | | | |
| Standard-of-care | 7·3 | 5·4 | 12·2 | 60·081 | 26·353 | 42 | Reference |
| HR-HIVE – 1 dose | 6·4 | 5·2 | 11·1 | 60·090 | 26·357 | 39 | dominated |
| HIVE – 1 dose | 6·3 | 5·1 | 11·0 | 60·091 | 26·357 | 38 | dominated |
| HR-HIVE – 2 doses | 6·4 | 4·9 | 10·8 | 60·092 | 26·357 | 39 | dominated |
| HIVE – 2 doses | 6·3 | 4·6 | 10·5 | 60·094 | 26·359 | 37 | dominated |
| HR-HIVE – Extended | 6·4 | 4·3 | 10·3 | 60·095 | 26·359 | 37 | dominated |
| ALL – 1 dose | 6·1 | 5·0 | 10·6 | 60·096 | 26·359 | 52 | dominated |
| ALL – 2 doses | 6·1 | 4·4 | 10·1 | 60·097 | 26·360 | 66 | dominated |
| HIVE – Extended | 6·3 | 3·4 | 9·4 | 60·102 | 26·361 | 33 | cost-saving*·† |
| ALL – 1 dose plus HIVE - Extended | 6·1 | 3·3 | 9·1 | 60·107 | 26·363 | 47 | 6244 |
| ALL - Extended | 6·1 | 2·9 | 8·6 | 60·113 | 26·365 | 112 | 57 665 |
| ***Proportion of mothers on ART during pregnancy: 100%*** | | | | | | | |
| Standard-of-care | 3·0 | 4·9 | 7·7 | 60·138 | 26·377 | 14 | Reference |
| HR-HIVE – 1 dose | 2·8 | 4·8 | 7·4 | 60·141 | 26·378 | 14 | dominated |
| HR-HIVE – 2 doses | 2·8 | 4·7 | 7·3 | 60·142 | 26·378 | 14 | dominated |
| HIVE – 1 dose | 2·7 | 4·7 | 7·2 | 60·143 | 26·379 | 13 | dominated |
| HR-HIVE – Extended | 2·8 | 4·3 | 7·0 | 60·144 | 26·379 | 14 | dominated |
| HIVE – 2 doses | 2·7 | 4·3 | 6·8 | 60·146 | 26·380 | 13 | dominated |
| ALL – 1 dose | 2·5 | 4·6 | 6·9 | 60·148 | 26·381 | 27 | dominated |
| ALL – 2 doses | 2·5 | 4·1 | 6·5 | 60·148 | 26·381 | 42 | dominated |
| HIVE – Extended | 2·7 | 3·1 | 5·7 | 60·154 | 26·383 | 12 | cost-saving*·† |
| ALL – 1 dose plus HIVE - Extended | 2·5 | 3·1 | 5·4 | 60·159 | 26·385 | 25 | 6242 |
| ALL - Extended | 2·5 | 2·6 | 5·0 | 60·164 | 26·386 | 90 | 57 888 |
| **South Africa [CET: ICER ≤ $1131/YLS (20% GDP per capita), ICER ≤ $2828/YLS (50% GDP per capita)]** | | | | | | | |
| ***Proportion of mothers on ART during pregnancy: 70%*** | | | | | | | |
| Standard-of-care | 8·1 | 2·6 | 10·1 | 67·590 | 28·013 | 232 | Reference |
| HR-HIVE – 1 dose | 6·4 | 2·3 | 8·3 | 67·824 | 28·091 | 209 | dominated |
| HIVE – 1 dose | 6·3 | 2·3 | 8·1 | 67·844 | 28·098 | 199 | dominated |
| HR-HIVE – 2 doses | 6·4 | 2·2 | 8·1 | 67·848 | 28·099 | 207 | dominated |
| HR-HIVE – Extended | 6·4 | 2·0 | 8·0 | 67·869 | 28·106 | 206 | dominated |
| HIVE – 2 doses | 6·3 | 2·0 | 7·9 | 67·884 | 28·112 | 196 | dominated |
| ALL – 1 dose | 6·2 | 2·0 | 7·8 | 67·895 | 28·116 | 209 | dominated |
| HIVE – Extended | 6·3 | 1·7 | 7·6 | 67·931 | 28·127 | 193 | cost-saving |
| ALL – 2 doses | 6·2 | 1·6 | 7·4 | 67·961 | 28·139 | 214 | dominated |
| ALL – 1 dose plus HIVE - Extended | 6·2 | 1·5 | 7·2 | 67·982 | 28·145 | 204 | 575 |
| ALL - Extended | 6·2 | 0·9 | 6·7 | 68·066 | 28·172 | 227 | 850*·† |

**Supplementary Table 13.** One-way sensitivity analysis: proportion of mothers on antiretroviral therapy during pregnancy (cont.)

| | Clinical outcomes | | | Lifetime efficacy and costs | | | |
|---|---|---|---|---|---|---|---|
| **Country/strategy** | **IU/IP cumulative HIV incidence (%)** | **Postnatal cumulative HIV incidence (%)** | **Total cumulative HIV incidence (%)** | **Undiscounted life expectancy (yrs)** | **Discounted life expectancy (yrs)** | **Discounted costs ($)** | **ICER ($/YLS)** |
| **South Africa [CET: ICER ≤ $1131/YLS (20% GDP per capita), ICER ≤ $2828/YLS (50% GDP per capita)]** | | | | | | | |
| ***Proportion of mothers on ART during pregnancy: 100%*** | | | | | | | |
| Standard-of-care | 2·0 | 2·2 | 4·0 | 69·018 | 28·534 | 104 | Reference |
| HR-HIVE – 1 dose | 1·8 | 2·1 | 3·7 | 69·061 | 28·548 | 109 | dominated |
| HR-HIVE – 2 doses | 1·8 | 2·0 | 3·7 | 69·067 | 28·550 | 108 | dominated |
| HR-HIVE – Extended | 1·8 | 2·0 | 3·7 | 69·075 | 28·553 | 108 | dominated |
| HIVE –1 dose | 1·7 | 2·0 | 3·5 | 69·094 | 28·560 | 99 | dominated |
| HIVE – 2 doses | 1·7 | 1·8 | 3·3 | 69·127 | 28·571 | 98 | dominated |
| ALL – 1 dose | 1·5 | 1·8 | 3·2 | 69·145 | 28·578 | 110 | dominated |
| HIVE – Extended | 1·7 | 1·5 | 3·0 | 69·177 | 28·588 | 94 | cost-saving |
| ALL – 2 doses | 1·5 | 1·4 | 2·9 | 69·204 | 28·598 | 116 | dominated |
| ALL – 1 dose plus HIVE - Extended | 1·5 | 1·3 | 2·7 | 69·229 | 28·606 | 105 | 606 |
| ALL - Extended | 1·5 | 0·8 | 2·2 | 69·310 | 28·632 | 129 | 884*·† |
| **Zimbabwe [CET: ICER ≤ $243/YLS (20% GDP per capita), ICER ≤ $607/YLS (50% GDP per capita)]** | | | | | | | |
| ***Proportion of mothers on ART during pregnancy: 70%*** | | | | | | | |
| Standard-of-care | 7·8 | 6·1 | 12·9 | 68·038 | 27·928 | 93 | Reference |
| HR-HIVE – 1 dose | 6·5 | 5·6 | 11·3 | 68·119 | 27·957 | 88 | dominated |
| HIVE – 1 dose | 6·4 | 5·5 | 11·1 | 68·130 | 27·960 | 84 | dominated |
| HR-HIVE – 2 doses | 6·5 | 5·3 | 11·0 | 68·133 | 27·961 | 87 | dominated |
| ALL– 1 dose | 6·2 | 5·2 | 10·6 | 68·150 | 27·967 | 97 | dominated |
| HR-HIVE – Extended | 6·5 | 4·8 | 10·6 | 68·156 | 27·969 | 85 | dominated |
| HIVE – 2 doses | 6·4 | 5·0 | 10·7 | 68·154 | 27·969 | 83 | dominated |
| ALL – 2 doses | 6·2 | 4·6 | 10·0 | 68·191 | 27·980 | 110 | dominated |
| HIVE – Extended | 6·4 | 3·9 | 9·6 | 68·208 | 27·986 | 78 | cost-saving*·† |
| ALL – 1 dose plus HIVE - Extended | 6·2 | 3·6 | 9·1 | 68·228 | 27·993 | 91 | 1739 |
| ALL - Extended | 6·2 | 2·7 | 8·3 | 68·267 | 28·007 | 151 | 4466 |
| ***Proportion of mothers on ART during pregnancy: 100%*** | | | | | | | |
| Standard-of-care | 2·7 | 5·4 | 7·8 | 68·461 | 28·087 | 61 | Reference |
| HR-HIVE – 1 dose | 2·5 | 5·2 | 7·4 | 68·488 | 28·096 | 62 | dominated |
| HR-HIVE – 2 doses | 2·5 | 5·1 | 7·3 | 68·492 | 28·098 | 62 | dominated |
| HR-HIVE – Extended | 2·5 | 4·9 | 7·1 | 68·502 | 28·101 | 61 | dominated |
| HIVE – 1 dose | 2·4 | 5·0 | 7·1 | 68·504 | 28·102 | 58 | dominated |
| HIVE – 2 doses | 2·4 | 4·6 | 6·8 | 68·525 | 28·109 | 58 | dominated |
| ALL – 1 dose | 2·2 | 4·7 | 6·7 | 68·524 | 28·109 | 71 | dominated |
| ALL – 2 doses | 2·2 | 4·2 | 6·2 | 68·561 | 28·121 | 85 | dominated |
| HIVE – Extended | 2·4 | 3·5 | 5·7 | 68·581 | 28·127 | 53 | cost-saving*·† |
| ALL – 1 dose plus HIVE - Extended | 2·2 | 3·3 | 5·2 | 68·602 | 28·135 | 66 | 1729 |
| ALL - Extended | 2·2 | 2·5 | 4·5 | 68·640 | 28·148 | 126 | 4533 |

*bNAb: broadly neutralizing antibody; IU/IP: intrauterine/intrapartum; yr: year; ICER: incremental cost-effectiveness ratio; YLS: years of life saved; CET: cost-effectiveness threshold; HR-HIVE: high-risk HIV-exposed infants; HIVE: all HIV-exposed infants; ALL: all live infants at birth.*

Pediatric HIV incidence is rounded to the nearest tenth of a percent. IU/IP HIV incidence is calculated based on the number of infants exposed to HIV at birth. Postnatal and total HIV incidence is calculated based on the number of infants ever exposed to HIV through 36 months of life. Undiscounted and discounted life expectancies are rounded to the nearest ten thousandth. Costs are rounded to the nearest dollar and are presented in 2020 USD. Discounted values are discounted at 3% per year. ICERs are rounded to the nearest dollar and are calculated using unrounded discounted life expectancy and discounted costs. The cost-effective bNAb strategy was the strategy that offered the greatest increase in overall population life expectancy while still having an ICER less than the cost-effectiveness threshold when compared to the next best performing, non-dominated strategy. * Indicates the cost-effective strategy using a cost-effectiveness threshold of 20% GDP per capita. † Indicates the cost-effective strategy using a cost-effectiveness threshold of 50% GDP per capita.

**Supplementary Table 14.** One-way sensitivity analysis: breastfeeding duration

| | Clinical outcomes | | | Lifetime efficacy and costs | | | |
|---|---|---|---|---|---|---|---|
| **Country/strategy** | **IU/IP cumulative HIV incidence (%)** | **Postnatal cumulative HIV incidence (%)** | **Total cumulative HIV incidence (%)** | **Undiscounted life expectancy (yrs)** | **Discounted life expectancy (yrs)** | **Discounted costs ($)** | **ICER ($/YLS)** |
| **Côte d'Ivoire [CET: ICER ≤ $465/YLS (20% GDP per capita), ICER ≤ $1163/YLS (50% GDP per capita)]** | | | | | | | |
| ***Mean breastfeeding duration: 8 months for all women*** | | | | | | | |
| Standard-of-care | 3·6 | 2·7 | 6·2 | 60·155 | 26·381 | 10 | Reference |
| HR-HIVE – 1 dose | 3·3 | 2·6 | 5·8 | 60·158 | 26·383 | 10 | dominated |
| HR-HIVE – 2 doses | 3·3 | 2·5 | 5·7 | 60·159 | 26·383 | 10 | dominated |
| HIVE – 1 dose | 3·3 | 2·5 | 5·7 | 60·160 | 26·383 | 9 | dominated |
| HR-HIVE – Extended | 3·3 | 2·3 | 5·5 | 60·160 | 26·384 | 10 | dominated |
| HIVE – 2 doses | 3·3 | 2·2 | 5·4 | 60·162 | 26·384 | 9 | dominated |
| ALL – 1 dose | 3·0 | 2·4 | 5·4 | 60·161 | 26·385 | 22 | dominated |
| HIVE – Extended | 3·3 | 1·6 | 4·8 | 60·166 | 26·386 | 9 | cost-saving*,† |
| ALL – 2 doses | 3·0 | 2·1 | 5·0 | 60·167 | 26·386 | 33 | dominated |
| ALL – 1 dose plus HIVE - Extended | 3·0 | 1·5 | 4·5 | 60·168 | 26·387 | 21 | 10 463 |
| ALL - Extended | 3·0 | 1·3 | 4·3 | 60·168 | 26·388 | 52 | 51 570 |
| ***Mean breastfeeding duration: 24 months for all women*** | | | | | | | |
| Standard-of-care | 3·6 | 7·9 | 11·2 | 60·121 | 26·369 | 20 | Reference |
| ALL – 1 dose | 3·0 | 7·6 | 10·3 | 60·120 | 26·370 | 34 | dominated |
| HR-HIVE – 1 dose | 3·3 | 7·8 | 10·8 | 60·125 | 26·371 | 20 | dominated |
| HR-HIVE – 2 doses | 3·3 | 7·7 | 10·7 | 60·126 | 26·371 | 20 | dominated |
| HIVE – 1 dose | 3·3 | 7·7 | 10·6 | 60·127 | 26·372 | 19 | dominated |
| ALL – 2 doses | 3·0 | 7·1 | 9·8 | 60·127 | 26·372 | 49 | dominated |
| HR-HIVE – Extended | 3·3 | 7·1 | 10·1 | 60·130 | 26·373 | 20 | dominated |
| HIVE – 2 doses | 3·3 | 7·2 | 10·2 | 60·130 | 26·373 | 19 | dominated |
| ALL – 1 dose plus HIVE - Extended | 3·0 | 5·3 | 8·1 | 60·136 | 26·376 | 31 | dominated |
| HIVE – Extended | 3·3 | 5·4 | 8·4 | 60·142 | 26·378 | 17 | cost-saving*,† |
| ALL - Extended | 3·0 | 4·8 | 7·6 | 60·146 | 26·380 | 103 | 40 559 |
| **South Africa [CET: ICER ≤ $1131/YLS (20% GDP per capita), ICER ≤ $2828/YLS (50% GDP per capita)]** | | | | | | | |
| ***Mean breastfeeding duration: 2 months for all women*** | | | | | | | |
| Standard-of-care | 2·3 | 0·9 | 3·2 | 69·125 | 28·566 | 84 | Reference |
| HR-HIVE – 1 dose | 2·1 | 0·9 | 2·9 | 69·178 | 28·583 | 87 | dominated |
| HR-HIVE – 2 doses | 2·1 | 0·8 | 2·9 | 69·181 | 28·585 | 87 | dominated |
| HR-HIVE – Extended | 2·1 | 0·8 | 2·8 | 69·185 | 28·586 | 87 | dominated |
| HIVE – 1 dose | 2·0 | 0·8 | 2·7 | 69·205 | 28·593 | 77 | dominated |
| HIVE – 2 doses | 2·0 | 0·7 | 2·6 | 69·225 | 28·600 | 76 | dominated |
| HIVE – Extended | 2·0 | 0·6 | 2·5 | 69·244 | 28·606 | 75 | cost-saving |
| ALL – 1 dose | 1·8 | 0·7 | 2·4 | 69·252 | 28·610 | 83 | dominated |
| ALL – 2 doses | 1·8 | 0·5 | 2·3 | 69·280 | 28·619 | 85 | dominated |
| ALL – 1 dose plus HIVE - Extended | 1·8 | 0·4 | 2·2 | 69·291 | 28·622 | 81 | 364* |
| ALL - Extended | 1·8 | 0·3 | 2·1 | 69·306 | 28·628 | 87 | 1145† |

**Supplementary Table 14.** One-way sensitivity analysis: breastfeeding duration (cont.)

| | Clinical outcomes | | | Lifetime efficacy and costs | | | |
|---|---|---|---|---|---|---|---|
| **Country/strategy** | **IU/IP cumulative HIV incidence (%)** | **Postnatal cumulative HIV incidence (%)** | **Total cumulative HIV incidence (%)** | **Undiscounted life expectancy (yrs)** | **Discounted life expectancy (yrs)** | **Discounted costs ($)** | **ICER ($/YLS)** |
| **South Africa [CET: ICER ≤ $1131/YLS (20% GDP per capita), ICER ≤ $2828/YLS (50% GDP per capita)]** | | | | | | | |
| ***Mean breastfeeding duration: 18 months for all women*** | | | | | | | |
| Standard-of-care | 2·3 | 4·9 | 6·9 | 68·494 | 28·361 | 175 | Reference |
| HR-HIVE – 1 dose | 2·1 | 4·7 | 6·6 | 68·553 | 28·381 | 178 | dominated |
| HR-HIVE – 2 doses | 2·1 | 4·7 | 6·5 | 68·563 | 28·385 | 178 | dominated |
| HIVE – 1 dose | 2·0 | 4·6 | 6·3 | 68·591 | 28·394 | 168 | dominated |
| HR-HIVE – Extended | 2·1 | 4·4 | 6·3 | 68·602 | 28·397 | 174 | dominated |
| HIVE – 2 doses | 2·0 | 4·3 | 6·0 | 68·645 | 28·413 | 166 | dominated |
| ALL – 1 dose | 1·8 | 4·4 | 6·0 | 68·645 | 28·413 | 180 | dominated |
| ALL – 2 doses | 1·8 | 3·9 | 5·6 | 68·717 | 28·439 | 186 | dominated |
| HIVE – Extended | 2·0 | 2·9 | 4·7 | 68·860 | 28·482 | 148 | cost-saving |
| ALL – 1 dose plus HIVE - Extended | 1·8 | 2·7 | 4·4 | 68·914 | 28·501 | 159 | 607 |
| ALL - Extended | 1·8 | 2·0 | 3·7 | 69·031 | 28·539 | 192 | 847*,† |
| **Zimbabwe [CET: ICER ≤ $243/YLS (20% GDP per capita), ICER ≤ $607/YLS (50% GDP per capita)]** | | | | | | | |
| ***Mean breastfeeding duration: 8 months for women with HIV and 15 months for women without HIV*** | | | | | | | |
| Standard-of-care | 4·3 | 4·1 | 8·0 | 68·397 | 28·058 | 59 | Reference |
| HR-HIVE – 1 dose | 3·7 | 3·8 | 7·2 | 68·440 | 28·074 | 58 | dominated |
| HR-HIVE – 2 doses | 3·7 | 3·7 | 7·1 | 68·446 | 28·076 | 57 | dominated |
| HIVE – 1 dose | 3·7 | 3·7 | 7·0 | 68·453 | 28·078 | 54 | dominated |
| HR-HIVE – Extended | 3·7 | 3·5 | 7·0 | 68·454 | 28·078 | 57 | dominated |
| HIVE – 2 doses | 3·7 | 3·3 | 6·7 | 68·470 | 28·084 | 53 | dominated |
| ALL – 1 dose | 3·4 | 3·4 | 6·5 | 68·480 | 28·087 | 67 | dominated |
| HIVE – Extended | 3·7 | 2·7 | 6·1 | 68·500 | 28·094 | 51 | cost-saving*,† |
| ALL – 2 doses | 3·4 | 2·9 | 6·1 | 68·502 | 28·096 | 80 | dominated |
| ALL – 1 dose plus HIVE - Extended | 3·4 | 2·5 | 5·6 | 68·527 | 28·103 | 64 | 1348 |
| ALL - Extended | 3·4 | 1·8 | 5·0 | 68·554 | 28·113 | 116 | 5362 |
| ***Mean breastfeeding duration: 20 months for all women*** | | | | | | | |
| Standard-of-care | 4·3 | 7·6 | 11·3 | 68·226 | 28·004 | 87 | Reference |
| HR-HIVE – 1 dose | 3·7 | 7·3 | 10·5 | 68·271 | 28·020 | 86 | dominated |
| HR-HIVE – 2 doses | 3·7 | 7·1 | 10·4 | 68·279 | 28·023 | 86 | dominated |
| HIVE – 1 dose | 3·7 | 7·1 | 10·3 | 68·285 | 28·025 | 82 | dominated |
| HR-HIVE – Extended | 3·7 | 6·7 | 9·9 | 68·301 | 28·030 | 84 | dominated |
| ALL – 1 dose | 3·4 | 6·8 | 9·8 | 68·298 | 28·031 | 95 | dominated |
| HIVE – 2 doses | 3·7 | 6·7 | 9·8 | 68·311 | 28·034 | 81 | dominated |
| ALL – 2 doses | 3·4 | 6·3 | 9·2 | 68·340 | 28·044 | 109 | dominated |
| HIVE – Extended | 3·7 | 4·9 | 8·1 | 68·398 | 28·062 | 74 | cost-saving*,† |
| ALL – 1 dose plus HIVE - Extended | 3·4 | 4·6 | 7·7 | 68·411 | 28·068 | 86 | 2140 |
| ALL - Extended | 3·4 | 3·8 | 6·8 | 68·467 | 28·086 | 151 | 3624 |

*bNAb: broadly neutralizing antibody; IU/IP: intrauterine/intrapartum; yr: year; ICER: incremental cost-effectiveness ratio; YLS: years of life saved; CET: cost-effectiveness threshold; HR-HIVE: high-risk HIV-exposed infants; HIVE: all HIV-exposed infants; ALL: all live infants at birth.*

Pediatric HIV incidence is rounded to the nearest tenth of a percent. IU/IP HIV incidence is calculated based on the number of infants exposed to HIV at birth. Postnatal and total HIV incidence is calculated based on the number of infants ever exposed to HIV through 36 months of life. Undiscounted and discounted life expectancies are rounded to the nearest ten thousandth. Costs are rounded to the nearest dollar and are presented in 2020 USD. Discounted values are discounted at 3% per year. ICERs are rounded to the nearest dollar and are calculated using unrounded discounted life expectancy and discounted costs. The cost-effective bNAb strategy was the strategy that offered the greatest increase in overall population life expectancy while still having an ICER less than the cost-effectiveness threshold when compared to the next best performing, non-dominated strategy. * Indicates the cost-effective strategy using a cost-effectiveness threshold of 20% GDP per capita. † Indicates the cost-effective strategy using a cost-effectiveness threshold of 50% GDP per capita.

**Supplementary Table 15.** One-way sensitivity analysis: maternal postpartum HIV incidence

| | Clinical outcomes | | | Lifetime efficacy and costs | | | |
|---|---|---|---|---|---|---|---|
| **Country/strategy** | **IU/IP cumulative HIV incidence (%)** | **Postnatal cumulative HIV incidence (%)** | **Total cumulative HIV incidence (%)** | **Undiscounted life expectancy (yrs)** | **Discounted life expectancy (yrs)** | **Discounted costs ($)** | **ICER ($/YLS)** |
| **Côte d'Ivoire [CET: ICER ≤ $465/YLS (20% GDP per capita), ICER ≤ $1163/YLS (50% GDP per capita)]** | | | | | | | |
| ***Postpartum incidence: 0·5x base case*** | | | | | | | |
| Standard-of-care | 3·6 | 4·9 | 8·3 | 60·130 | 26·374 | 14 | Reference |
| HR-HIVE – 1 dose | 3·3 | 4·7 | 7·9 | 60·134 | 26·375 | 14 | dominated |
| HR-HIVE – 2 doses | 3·3 | 4·6 | 7·7 | 60·135 | 26·375 | 14 | dominated |
| HIVE – 1 dose | 3·3 | 4·6 | 7·7 | 60·135 | 26·376 | 13 | dominated |
| HR-HIVE – Extended | 3·3 | 4·2 | 7·4 | 60·137 | 26·376 | 13 | dominated |
| HIVE – 2 doses | 3·3 | 4·2 | 7·3 | 60·138 | 26·377 | 13 | dominated |
| ALL – 1 dose | 3·0 | 4·5 | 7·4 | 60·141 | 26·378 | 27 | dominated |
| ALL – 2 doses | 3·0 | 4·0 | 6·9 | 60·141 | 26·378 | 43 | dominated |
| HIVE – Extended | 3·3 | 3·0 | 6·2 | 60·146 | 26·380 | 12 | cost-saving*·† |
| ALL – 1 dose plus HIVE - Extended | 3·0 | 3·0 | 5·9 | 60·152 | 26·382 | 26 | 6337 |
| ALL - Extended | 3·0 | 2·6 | 5·5 | 60·156 | 26·383 | 91 | 75 319 |
| ***Postpartum incidence: 2·0x base case*** | | | | | | | |
| Standard-of-care | 3·6 | 5·2 | 8·5 | 60·126 | 26·372 | 15 | Reference |
| HR-HIVE – 1 dose | 3·3 | 5·1 | 8·1 | 60·130 | 26·374 | 15 | dominated |
| HR-HIVE – 2 doses | 3·3 | 5·0 | 8·0 | 60·131 | 26·374 | 15 | dominated |
| HIVE – 1 dose | 3·3 | 5·0 | 8·0 | 60·131 | 26·374 | 14 | dominated |
| HR-HIVE –Extended | 3·3 | 4·6 | 7·7 | 60·133 | 26·375 | 14 | dominated |
| HIVE – 2 doses | 3·3 | 4·6 | 7·6 | 60·135 | 26·376 | 14 | dominated |
| ALL – 1 dose | 3·0 | 4·9 | 7·6 | 60·137 | 26·377 | 28 | dominated |
| ALL – 2 doses | 3·0 | 4·4 | 7·2 | 60·138 | 26·377 | 44 | dominated |
| HIVE – Extended | 3·3 | 3·4 | 6·5 | 60·143 | 26·378 | 13 | cost-saving*·† |
| ALL – 1 dose plus HIVE - Extended | 3·0 | 3·3 | 6·2 | 60·149 | 26·381 | 27 | 5993 |
| ALL - Extended | 3·0 | 2·8 | 5·6 | 60·154 | 26·382 | 91 | 58 257 |
| **South Africa [CET: ICER ≤ $1131/YLS (20% GDP per capita), ICER ≤ $2828/YLS (50% GDP per capita)]** | | | | | | | |
| ***Postpartum incidence: 0·5x base case*** | | | | | | | |
| Standard-of-care | 2·3 | 1·9 | 4·1 | 68·984 | 28·519 | 104 | Reference |
| HR-HIVE – 1 dose | 2·1 | 1·8 | 3·7 | 69·039 | 28·538 | 108 | dominated |
| HR-HIVE – 2 doses | 2·1 | 1·7 | 3·7 | 69·046 | 28·541 | 107 | dominated |
| HR-HIVE – Extended | 2·1 | 1·7 | 3·6 | 69·055 | 28·543 | 107 | dominated |
| HIVE – 1 dose | 2·0 | 1·7 | 3·5 | 69·071 | 28·549 | 98 | dominated |
| HIVE – 2 doses | 2·0 | 1·5 | 3·3 | 69·105 | 28·561 | 96 | dominated |
| ALL – 1 dose | 1·8 | 1·5 | 3·2 | 69·116 | 28·565 | 109 | dominated |
| HIVE – Extended | 2·0 | 1·1 | 3·0 | 69·155 | 28·577 | 93 | cost-saving |
| ALL – 2 doses | 1·8 | 1·2 | 2·9 | 69·169 | 28·583 | 116 | dominated |
| ALL – 1 dose plus HIVE - Extended | 1·8 | 1·0 | 2·7 | 69·200 | 28·593 | 104 | 700* |
| ALL - Extended | 1·8 | 0·7 | 2·4 | 69·251 | 28·609 | 131 | 1,675† |

**Supplementary Table 15.** One-way sensitivity analysis: maternal postpartum HIV incidence (cont.)

| | Clinical outcomes | | | Lifetime efficacy and costs | | | |
|---|---|---|---|---|---|---|---|
| **Country/strategy** | **IU/IP cumulative HIV incidence (%)** | **Postnatal cumulative HIV incidence (%)** | **Total cumulative HIV incidence (%)** | **Undiscounted life expectancy (yrs)** | **Discounted life expectancy (yrs)** | **Discounted costs ($)** | **ICER ($/YLS)** |
| **South Africa [CET: ICER ≤ $1131/YLS (20% GDP per capita), ICER ≤ $2828/YLS (50% GDP per capita)]** | | | | | | | |
| ***Postpartum incidence: 2·0x base case*** | | | | | | | |
| Standard-of-care | 2·3 | 2·8 | 4·8 | 68·813 | 28·462 | 127 | Reference |
| HR-HIVE – 1 dose | 2·1 | 2·7 | 4·5 | 68·868 | 28·481 | 130 | dominated |
| HR-HIVE – 2 doses | 2·1 | 2·6 | 4·5 | 68·875 | 28·483 | 130 | dominated |
| HR-HIVE – Extended | 2·1 | 2·6 | 4·4 | 68·884 | 28·486 | 129 | dominated |
| HIVE – 1 dose | 2·0 | 2·6 | 4·3 | 68·900 | 28·492 | 121 | dominated |
| HIVE – 2 doses | 2·0 | 2·4 | 4·1 | 68·934 | 28·504 | 119 | dominated |
| ALL – 1 dose | 1·8 | 2·3 | 3·9 | 68·960 | 28·513 | 130 | dominated |
| HIVE – Extended | 2·0 | 2·1 | 3·8 | 68·984 | 28·520 | 116 | cost-saving |
| ALL – 2 doses | 1·8 | 1·9 | 3·5 | 69·034 | 28·540 | 135 | dominated |
| ALL – 1 dose plus HIVE - Extended | 1·8 | 1·8 | 3·5 | 69·043 | 28·541 | 125 | dominated |
| ALL - Extended | 1·8 | 1·0 | 2·6 | 69·183 | 28·588 | 140 | 359*·† |
| **Zimbabwe [CET: ICER ≤ $243/YLS (20% GDP per capita), ICER ≤ $607/YLS (50% GDP per capita)]** | | | | | | | |
| ***Postpartum incidence: 0·5x base case*** | | | | | | | |
| Standard-of-care | 4·3 | 5·2 | 9·2 | 68·351 | 28·044 | 66 | Reference |
| HR-HIVE – 1 dose | 3·7 | 4·9 | 8·3 | 68·396 | 28·060 | 65 | dominated |
| HR-HIVE – 2 doses | 3·7 | 4·7 | 8·2 | 68·403 | 28·062 | 65 | dominated |
| HIVE – 1 dose | 3·7 | 4·7 | 8·1 | 68·410 | 28·065 | 61 | dominated |
| HR-HIVE – Extended | 3·7 | 4·4 | 7·9 | 68·417 | 28·067 | 64 | dominated |
| ALL – 1 dose | 3·4 | 4·4 | 7·6 | 68·430 | 28·072 | 75 | dominated |
| HIVE – 2 doses | 3·7 | 4·3 | 7·7 | 68·432 | 28·073 | 61 | dominated |
| ALL – 2 doses | 3·4 | 3·9 | 7·1 | 68·466 | 28·083 | 89 | dominated |
| HIVE – Extended | 3·7 | 3·1 | 6·6 | 68·488 | 28·090 | 56 | cost-saving*·† |
| ALL – 1 dose plus HIVE - Extended | 3·4 | 2·9 | 6·1 | 68·508 | 28·097 | 70 | 1976 |
| ALL - Extended | 3·4 | 2·3 | 5·6 | 68·534 | 28·107 | 131 | 6572 |
| ***Postpartum incidence: 2·0x base case*** | | | | | | | |
| Standard-of-care | 4·3 | 6·4 | 10·0 | 68·258 | 28·014 | 68 | Reference |
| HR-HIVE – 1 dose | 3·7 | 6·1 | 9·2 | 68·303 | 28·030 | 68 | dominated |
| HR-HIVE – 2 doses | 3·7 | 6·0 | 9·1 | 68·310 | 28·032 | 68 | dominated |
| HIVE – 1 dose | 3·7 | 6·0 | 9·0 | 68·317 | 28·035 | 68 | dominated |
| HR-HIVE – Extended | 3·7 | 5·7 | 8·8 | 68·325 | 28·037 | 68 | dominated |
| HIVE – 2 doses | 3·7 | 5·6 | 8·6 | 68·339 | 28·042 | 68 | dominated |
| ALL – 1 dose | 3·4 | 5·7 | 8·5 | 68·342 | 28·043 | 68 | dominated |
| ALL – 2 doses | 3·4 | 5·0 | 7·9 | 68·387 | 28·058 | 68 | dominated |
| HIVE – Extended | 3·7 | 4·5 | 7·6 | 68·395 | 28·060 | 68 | cost-saving*·† |
| ALL – 1 dose plus HIVE - Extended | 3·4 | 4·2 | 7·1 | 68·420 | 28·069 | 68 | 1503 |
| ALL - Extended | 3·4 | 3·0 | 5·9 | 68·486 | 28·092 | 68 | 2414 |

*bNAb: broadly neutralizing antibody; IU/IP: intrauterine/intrapartum; yr: year; ICER: incremental cost-effectiveness ratio; YLS: years of life saved; CET: cost-effectiveness threshold; HR-HIVE: high-risk HIV-exposed infants; HIVE: all HIV-exposed infants; ALL: all live infants at birth.*

Pediatric HIV incidence is rounded to the nearest tenth of a percent. IU/IP HIV incidence is calculated based on the number of infants exposed to HIV at birth. Postnatal and total HIV incidence is calculated based on the number of infants ever exposed to HIV through 36 months of life. Undiscounted and discounted life expectancies are rounded to the nearest ten thousandth. Costs are rounded to the nearest dollar and are presented in 2020 USD. Discounted values are discounted at 3% per year. ICERs are rounded to the nearest dollar and are calculated using unrounded discounted life expectancy and discounted costs. The cost-effective bNAb strategy was the strategy that offered the greatest increase in overall population life expectancy while still having an ICER less than the cost-effectiveness threshold when compared to the next best performing, non-dominated strategy. * Indicates the cost-effective strategy using a cost-effectiveness threshold of 20% GDP per capita. † Indicates the cost-effective strategy using a cost-effectiveness threshold of 50% GDP per capita.

**Supplementary Table 16.** One-way sensitivity analysis: antiretroviral therapy treatment cost

| | Clinical outcomes | | | Lifetime efficacy and costs | | | |
|---|---|---|---|---|---|---|---|
| **Country/strategy** | **IU/IP cumulative HIV incidence (%)** | **Postnatal cumulative HIV incidence (%)** | **Total cumulative HIV incidence (%)** | **Undiscounted life expectancy (yrs)** | **Discounted life expectancy (yrs)** | **Discounted costs ($)** | **ICER ($/YLS)** |
| **Côte d'Ivoire [CET: ICER ≤ $465/YLS (20% GDP per capita), ICER ≤ $1163/YLS (50% GDP per capita)]** | | | | | | | |
| ***ART treatment cost: 0·5x base case*** | | | | | | | |
| Standard-of-care | 3·6 | 5·0 | 8·4 | 60·129 | 26·373 | 12 | Reference |
| HR-HIVE – 1 dose | 3·3 | 4·8 | 8·0 | 60·133 | 26·375 | 12 | dominated |
| HR-HIVE – 2 doses | 3·3 | 4·7 | 7·8 | 60·134 | 26·375 | 12 | dominated |
| HIVE – 1 dose | 3·3 | 4·7 | 7·8 | 60·134 | 26·375 | 11 | dominated |
| HR-HIVE – Extended | 3·3 | 4·3 | 7·5 | 60·136 | 26·376 | 12 | dominated |
| HIVE – 2 doses | 3·3 | 4·3 | 7·4 | 60·138 | 26·377 | 11 | dominated |
| ALL – 1 dose | 3·0 | 4·6 | 7·5 | 60·139 | 26·377 | 26 | dominated |
| ALL – 2 doses | 3·0 | 4·2 | 7·0 | 60·140 | 26·378 | 41 | dominated |
| HIVE – Extended | 3·3 | 3·2 | 6·3 | 60·146 | 26·379 | 10 | cost-saving*·† |
| ALL – 1 dose plus HIVE - Extended | 3·0 | 3·1 | 6·0 | 60·150 | 26·382 | 25 | 6722 |
| ALL - Extended | 3·0 | 2·7 | 5·5 | 60·156 | 26·383 | 89 | 57 928 |
| ***ART treatment cost: 2·0x base case*** | | | | | | | |
| Standard-of-care | 3·6 | 5·0 | 8·4 | 60·129 | 26·373 | 18 | Reference |
| HR-HIVE – 1 dose | 3·3 | 4·8 | 8·0 | 60·133 | 26·375 | 19 | dominated |
| HR-HIVE – 2 doses | 3·3 | 4·7 | 7·8 | 60·134 | 26·375 | 18 | dominated |
| HIVE – 1 dose | 3·3 | 4·7 | 7·8 | 60·134 | 26·375 | 18 | dominated |
| HR-HIVE – Extended | 3·3 | 4·3 | 7·5 | 60·136 | 26·376 | 18 | dominated |
| HIVE – 2 doses | 3·3 | 4·3 | 7·4 | 60·138 | 26·377 | 17 | dominated |
| ALL – 1 dose | 3·0 | 4·6 | 7·5 | 60·139 | 26·377 | 31 | dominated |
| ALL – 2 doses | 3·0 | 4·2 | 7·0 | 60·140 | 26·378 | 47 | dominated |
| HIVE – Extended | 3·3 | 3·2 | 6·3 | 60·146 | 26·379 | 15 | cost-saving*·† |
| ALL – 1 dose plus HIVE - Extended | 3·0 | 3·1 | 6·0 | 60·150 | 26·382 | 29 | 6197 |
| ALL - Extended | 3·0 | 2·7 | 5·5 | 60·156 | 26·383 | 93 | 57 720 |
| **South Africa [CET: ICER ≤ $1131/YLS (20% GDP per capita), ICER ≤ $2828/YLS (50% GDP per capita)]** | | | | | | | |
| ***ART treatment cost: 0·5x base case*** | | | | | | | |
| Standard-of-care | 2·3 | 2·2 | 4·4 | 68·924 | 28·499 | 103 | Reference |
| HR-HIVE – 1 dose | 2·1 | 2·1 | 4·0 | 68·979 | 28·518 | 108 | dominated |
| HR-HIVE – 2 doses | 2·1 | 2·1 | 4·0 | 68·986 | 28·521 | 107 | dominated |
| HR-HIVE – Extended | 2·1 | 2·0 | 3·9 | 68·995 | 28·523 | 107 | dominated |
| HIVE – 1 dose | 2·0 | 2·0 | 3·8 | 69·011 | 28·529 | 98 | dominated |
| HIVE – 2 doses | 2·0 | 1·8 | 3·6 | 69·045 | 28·541 | 97 | dominated |
| ALL – 1 dose | 1·8 | 1·8 | 3·5 | 69·063 | 28·548 | 109 | dominated |
| HIVE – Extended | 2·0 | 1·5 | 3·3 | 69·095 | 28·557 | 94 | cost-saving |
| ALL – 2 doses | 1·8 | 1·5 | 3·2 | 69·122 | 28·568 | 116 | dominated |
| ALL – 1 dose plus HIVE - Extended | 1·8 | 1·3 | 3·0 | 69·147 | 28·575 | 105 | 596 |
| ALL - Extended | 1·8 | 0·8 | 2·5 | 69·228 | 28·602 | 129 | 913*·† |

**Supplementary Table 16.** One-way sensitivity analysis: antiretroviral therapy treatment cost (cont.)

| | Clinical outcomes | | | Lifetime efficacy and costs | | | |
|---|---|---|---|---|---|---|---|
| **Country/strategy** | **IU/IP cumulative HIV incidence (%)** | **Postnatal cumulative HIV incidence (%)** | **Total cumulative HIV incidence (%)** | **Undiscounted life expectancy (yrs)** | **Discounted life expectancy (yrs)** | **Discounted costs ($)** | **ICER ($/YLS)** |
| **South Africa [CET: ICER ≤ $1131/YLS (20% GDP per capita), ICER ≤ $2828/YLS (50% GDP per capita)]** | | | | | | | |
| ***ART treatment cost: 2·0x base case*** | | | | | | | |
| Standard-of-care | 2·3 | 2·2 | 4·4 | 68·924 | 28·499 | 130 | Reference |
| HR-HIVE – 1 dose | 2·1 | 2·1 | 4·0 | 68·979 | 28·518 | 131 | dominated |
| HR-HIVE – 2 doses | 2·1 | 2·1 | 4·0 | 68·986 | 28·521 | 131 | dominated |
| HR-HIVE – Extended | 2·1 | 2·0 | 3·9 | 68·995 | 28·523 | 130 | dominated |
| HIVE – 1 dose | 2·0 | 2·0 | 3·8 | 69·011 | 28·529 | 120 | dominated |
| HIVE – 2 doses | 2·0 | 1·8 | 3·6 | 69·045 | 28·541 | 119 | dominated |
| ALL – 1 dose | 1·8 | 1·8 | 3·5 | 69·063 | 28·548 | 130 | dominated |
| HIVE – Extended | 2·0 | 1·5 | 3·3 | 69·095 | 28·557 | 114 | cost-saving |
| ALL – 2 doses | 1·8 | 1·5 | 3·2 | 69·122 | 28·568 | 135 | dominated |
| ALL – 1 dose plus HIVE - Extended | 1·8 | 1·3 | 3·0 | 69·147 | 28·575 | 124 | 518 |
| ALL - Extended | 1·8 | 0·8 | 2·5 | 69·228 | 28·602 | 145 | 819*·† |
| **Zimbabwe [CET: ICER ≤ $243/YLS (20% GDP per capita), ICER ≤ $607/YLS (50% GDP per capita)]** | | | | | | | |
| ***ART treatment cost: 0·5x base case*** | | | | | | | |
| Standard-of-care | 4·3 | 5·6 | 9·5 | 68·321 | 28·034 | 62 | Reference |
| HR-HIVE – 1 dose | 3·7 | 5·3 | 8·6 | 68·366 | 28·050 | 61 | dominated |
| HR-HIVE – 2 doses | 3·7 | 5·2 | 8·5 | 68·373 | 28·053 | 61 | dominated |
| HIVE – 1 dose | 3·7 | 5·2 | 8·4 | 68·380 | 28·055 | 58 | dominated |
| HR-HIVE – Extended | 3·7 | 4·9 | 8·2 | 68·387 | 28·057 | 60 | dominated |
| ALL – 1 dose | 3·4 | 4·9 | 7·9 | 68·400 | 28·062 | 71 | dominated |
| HIVE – 2 doses | 3·7 | 4·8 | 8·0 | 68·402 | 28·063 | 57 | dominated |
| ALL – 2 doses | 3·4 | 4·3 | 7·4 | 68·438 | 28·074 | 85 | dominated |
| HIVE – Extended | 3·7 | 3·6 | 6·9 | 68·458 | 28·081 | 54 | cost-saving*·† |
| ALL – 1 dose plus HIVE - Extended | 3·4 | 3·4 | 6·5 | 68·478 | 28·088 | 67 | 1777 |
| ALL - Extended | 3·4 | 2·6 | 5·7 | 68·517 | 28·101 | 128 | 4601 |
| ***ART treatment cost: 2·0x base case*** | | | | | | | |
| Standard-of-care | 4·3 | 5·6 | 9·5 | 68·321 | 28·034 | 91 | Reference |
| HR-HIVE – 1 dose | 3·7 | 5·3 | 8·6 | 68·366 | 28·050 | 88 | dominated |
| HR-HIVE – 2 doses | 3·7 | 5·2 | 8·5 | 68·373 | 28·053 | 87 | dominated |
| HIVE – 1 dose | 3·7 | 5·2 | 8·4 | 68·380 | 28·055 | 84 | dominated |
| HR-HIVE – Extended | 3·7 | 4·9 | 8·2 | 68·387 | 28·057 | 86 | dominated |
| ALL – 1 dose | 3·4 | 4·9 | 7·9 | 68·400 | 28·062 | 95 | dominated |
| HIVE – 2 doses | 3·7 | 4·8 | 8·0 | 68·402 | 28·063 | 82 | dominated |
| ALL – 2 doses | 3·4 | 4·3 | 7·4 | 68·438 | 28·074 | 108 | dominated |
| HIVE – Extended | 3·7 | 3·6 | 6·9 | 68·458 | 28·081 | 75 | cost-saving*·† |
| ALL – 1 dose plus HIVE - Extended | 3·4 | 3·4 | 6·5 | 68·478 | 28·088 | 86 | 1522 |
| ALL - Extended | 3·4 | 2·6 | 5·7 | 68·517 | 28·101 | 145 | 4411 |

*bNAb: broadly neutralizing antibody; IU/IP: intrauterine/intrapartum; yr: year; ICER: incremental cost-effectiveness ratio; YLS: years of life saved; CET: cost-effectiveness threshold; HR-HIVE: high-risk HIV-exposed infants; HIVE: all HIV-exposed infants; ALL: all live infants at birth.*

Pediatric HIV incidence is rounded to the nearest tenth of a percent. IU/IP HIV incidence is calculated based on the number of infants exposed to HIV at birth. Postnatal and total HIV incidence is calculated based on the number of infants ever exposed to HIV through 36 months of life. Undiscounted and discounted life expectancies are rounded to the nearest ten thousandth. Costs are rounded to the nearest dollar and are presented in 2020 USD. Discounted values are discounted at 3% per year. ICERs are rounded to the nearest dollar and are calculated using unrounded discounted life expectancy and discounted costs. The cost-effective bNAb strategy was the strategy that offered the greatest increase in overall population life expectancy while still having an ICER less than the cost-effectiveness threshold when compared to the next best performing, non-dominated strategy. * Indicates the cost-effective strategy using a cost-effectiveness threshold of 20% GDP per capita. † Indicates the cost-effective strategy using a cost-effectiveness threshold of 50% GDP per capita.

**Supplementary Table 17.** One-way sensitivity analysis: postpartum vertical transmission risk

| | Clinical outcomes | | | Lifetime efficacy and costs | | | |
|---|---|---|---|---|---|---|---|
| **Country/strategy** | **IU/IP cumulative HIV incidence (%)** | **Postnatal cumulative HIV incidence (%)** | **Total cumulative HIV incidence (%)** | **Undiscounted life expectancy (yrs)** | **Discounted life expectancy (yrs)** | **Discounted costs ($)** | **ICER ($/YLS)** |
| **Côte d'Ivoire [CET: ICER ≤ $465/YLS (20% GDP per capita), ICER ≤ $1163/YLS (50% GDP per capita)]** | | | | | | | |
| ***Postpartum vertical transmission risk: 0·5x base case*** | | | | | | | |
| Standard-of-care | 3·6 | 2·5 | 6·0 | 60·147 | 26·380 | 10 | Reference |
| HR-HIVE – 1 dose | 3·3 | 2·5 | 5·7 | 60·150 | 26·381 | 10 | dominated |
| HR-HIVE – 2 doses | 3·3 | 2·4 | 5·6 | 60·150 | 26·381 | 10 | dominated |
| HIVE – 1 dose | 3·3 | 2·4 | 5·5 | 60·151 | 26·382 | 9 | dominated |
| HR-HIVE – Extended | 3·3 | 2·2 | 5·4 | 60·152 | 26·382 | 10 | dominated |
| HIVE – 2 doses | 3·3 | 2·2 | 5·3 | 60·153 | 26·382 | 9 | dominated |
| ALL – 2 doses | 3·0 | 2·1 | 5·0 | 60·155 | 26·383 | 39 | dominated |
| ALL – 1 dose | 3·0 | 2·4 | 5·3 | 60·156 | 26·384 | 24 | dominated |
| HIVE – Extended | 3·3 | 1·6 | 4·8 | 60·157 | 26·384 | 9 | cost-saving*·† |
| ALL – 1 dose plus HIVE - Extended | 3·0 | 1·6 | 4·5 | 60·161 | 26·386 | 24 | 7038 |
| ALL - Extended | 3·0 | 1·3 | 4·3 | 60·166 | 26·386 | 89 | 112 676 |
| ***Postpartum vertical transmission risk: 2·0x base case*** | | | | | | | |
| Standard-of-care | 3·6 | 9·6 | 12·8 | 60·096 | 26·361 | 22 | Reference |
| HR-HIVE – 1 dose | 3·3 | 9·3 | 12·3 | 60·101 | 26·362 | 22 | dominated |
| HR-HIVE – 2 doses | 3·3 | 9·1 | 12·0 | 60·103 | 26·363 | 22 | dominated |
| HIVE – 1 dose | 3·3 | 9·1 | 12·0 | 60·103 | 26·364 | 21 | dominated |
| HR-HIVE – Extended | 3·3 | 8·4 | 11·3 | 60·107 | 26·365 | 21 | dominated |
| ALL – 1 dose | 3·0 | 8·9 | 11·6 | 60·109 | 26·366 | 35 | dominated |
| HIVE – 2 doses | 3·3 | 8·3 | 11·2 | 60·110 | 26·366 | 20 | dominated |
| ALL – 2 doses | 3·0 | 8·0 | 10·7 | 60·113 | 26·368 | 50 | dominated |
| HIVE – Extended | 3·3 | 6·1 | 9·1 | 60·125 | 26·372 | 17 | cost-saving*·† |
| ALL – 1 dose plus HIVE - Extended | 3·0 | 5·9 | 8·7 | 60·130 | 26·374 | 31 | 6027 |
| ALL - Extended | 3·0 | 5·2 | 8·0 | 60·138 | 26·376 | 95 | 32 149 |
| **South Africa [CET: ICER ≤ $1131/YLS (20% GDP per capita), ICER ≤ $2828/YLS (50% GDP per capita)]** | | | | | | | |
| ***Postpartum vertical transmission risk: 0·5x base case*** | | | | | | | |
| Standard-of-care | 2·3 | 1·1 | 3·3 | 69·097 | 28·558 | 90 | Reference |
| HR-HIVE – 1 dose | 2·1 | 1·1 | 3·0 | 69·146 | 28·574 | 94 | dominated |
| HR-HIVE – 2 doses | 2·1 | 1·1 | 3·0 | 69·149 | 28·576 | 94 | dominated |
| HR-HIVE – Extended | 2·1 | 1·0 | 3·0 | 69·154 | 28·577 | 94 | dominated |
| HIVE – 1 dose | 2·0 | 1·0 | 2·9 | 69·169 | 28·582 | 85 | cost-saving |
| HIVE – 2 doses | 2·0 | 0·9 | 2·8 | 69·187 | 28·589 | 85 | 103 |
| ALL – 1 dose | 1·8 | 0·9 | 2·6 | 69·205 | 28·595 | 97 | dominated |
| HIVE – Extended | 2·0 | 0·8 | 2·6 | 69·212 | 28·597 | 85 | dominated |
| ALL – 2 doses | 1·8 | 0·7 | 2·5 | 69·237 | 28·606 | 107 | dominated |
| ALL – 1 dose plus HIVE - Extended | 1·8 | 0·7 | 2·4 | 69·248 | 28·609 | 98 | 590* |
| ALL - Extended | 1·8 | 0·4 | 2·1 | 69·291 | 28·623 | 126 | 2023† |

**Supplementary Table 17.** One-way sensitivity analysis: postpartum vertical transmission risk (cont.)

| | Clinical outcomes | | | Lifetime efficacy and costs | | | |
|---|---|---|---|---|---|---|---|
| Country/strategy | IU/IP cumulative HIV incidence (%) | Postnatal cumulative HIV incidence (%) | Total cumulative HIV incidence (%) | Undiscounted life expectancy (yrs) | Discounted life expectancy (yrs) | Discounted costs ($) | ICER ($/YLS) |
| **South Africa [CET: ICER ≤ $1131/YLS (20% GDP per capita), ICER ≤ $2828/YLS (50% GDP per capita)]** | | | | | | | |
| ***Postpartum vertical transmission risk: 2·0x base case*** | | | | | | | |
| Standard-of-care | 2·3 | 4·2 | 6·3 | 68·599 | 28·390 | 154 | Reference |
| HR-HIVE – 1 dose | 2·1 | 4·0 | 5·9 | 68·668 | 28·413 | 156 | dominated |
| HR-HIVE – 2 doses | 2·1 | 3·9 | 5·8 | 68·681 | 28·418 | 155 | dominated |
| HR-HIVE – Extended | 2·1 | 3·8 | 5·7 | 68·698 | 28·423 | 153 | dominated |
| HIVE – 1 dose | 2·0 | 3·8 | 5·6 | 68·719 | 28·431 | 143 | dominated |
| HIVE – 2 doses | 2·0 | 3·4 | 5·2 | 68·784 | 28·453 | 139 | dominated |
| ALL – 1 dose | 1·8 | 3·4 | 5·1 | 68·797 | 28·459 | 151 | dominated |
| HIVE – Extended | 2·0 | 2·8 | 4·6 | 68·880 | 28·484 | 129 | cost-saving |
| ALL – 2 doses | 1·8 | 2·8 | 4·5 | 68·904 | 28·496 | 151 | dominated |
| ALL – 1 dose plus HIVE - Extended | 1·8 | 2·4 | 4·1 | 68·958 | 28·512 | 136 | 260 |
| ALL - Extended | 1·8 | 1·6 | 3·3 | 69·103 | 28·560 | 151 | 315*,† |
| **Zimbabwe [CET: ICER ≤ $243/YLS (20% GDP per capita), ICER ≤ $607/YLS (50% GDP per capita)]** | | | | | | | |
| ***Postpartum vertical transmission risk: 0·5x base case*** | | | | | | | |
| Standard-of-care | 4·3 | 2·9 | 6·8 | 68·461 | 28·081 | 52 | Reference |
| HR-HIVE – 1 dose | 3·7 | 2·7 | 6·1 | 68·499 | 28·094 | 52 | dominated |
| HR-HIVE – 2 doses | 3·7 | 2·7 | 6·1 | 68·502 | 28·095 | 52 | dominated |
| HIVE – 1 dose | 3·7 | 2·6 | 6·0 | 68·508 | 28·097 | 48 | cost-saving |
| HR-HIVE – Extended | 3·7 | 2·5 | 5·9 | 68·510 | 28·098 | 52 | dominated |
| HIVE – 2 doses | 3·7 | 2·4 | 5·8 | 68·519 | 28·101 | 49 | dominated |
| ALL – 1 dose | 3·4 | 2·5 | 5·6 | 68·522 | 28·102 | 61 | dominated |
| ALL – 2 doses | 3·4 | 2·2 | 5·3 | 68·546 | 28·109 | 77 | dominated |
| HIVE – Extended | 3·7 | 1·9 | 5·2 | 68·548 | 28·110 | 49 | 18*,† |
| ALL – 1 dose plus HIVE - Extended | 3·4 | 1·7 | 4·9 | 68·562 | 28·115 | 62 | 2656 |
| ALL - Extended | 3·4 | 1·3 | 4·5 | 68·582 | 28·123 | 124 | 8712 |
| ***Postpartum vertical transmission risk: 2·0x base case*** | | | | | | | |
| Standard-of-care | 4·3 | 10·7 | 14·3 | 68·060 | 27·946 | 107 | Reference |
| HR-HIVE – 1 dose | 3·7 | 10·1 | 13·2 | 68·118 | 27·967 | 105 | dominated |
| HR-HIVE – 2 doses | 3·7 | 9·9 | 13·0 | 68·132 | 27·972 | 104 | dominated |
| HIVE – 1 dose | 3·7 | 9·8 | 12·9 | 68·141 | 27·976 | 100 | dominated |
| HR-HIVE – Extended | 3·7 | 9·3 | 12·5 | 68·159 | 27·981 | 100 | dominated |
| ALL – 1 dose | 3·4 | 9·3 | 12·2 | 68·173 | 27·987 | 112 | dominated |
| HIVE – 2 doses | 3·7 | 9·0 | 12·1 | 68·185 | 27·991 | 97 | dominated |
| ALL – 2 doses | 3·4 | 8·3 | 11·2 | 68·239 | 28·009 | 122 | dominated |
| HIVE – Extended | 3·7 | 6·8 | 10·0 | 68·292 | 28·025 | 84 | cost-saving*,† |
| ALL – 1 dose plus HIVE - Extended | 3·4 | 6·4 | 9·4 | 68·323 | 28·036 | 96 | 1061 |
| ALL - Extended | 3·4 | 5·0 | 8·0 | 68·393 | 28·060 | 152 | 2370 |

*bNAb: broadly neutralizing antibody; IU/IP: intrauterine/intrapartum; yr: year; ICER: incremental cost-effectiveness ratio; YLS: years of life saved; CET: cost-effectiveness threshold; HR-HIVE: high-risk HIV-exposed infants; HIVE: all HIV-exposed infants; ALL: all live infants at birth.*

Pediatric HIV incidence is rounded to the nearest tenth of a percent. IU/IP HIV incidence is calculated based on the number of infants exposed to HIV at birth. Postnatal and total HIV incidence is calculated based on the number of infants ever exposed to HIV through 36 months of life. Undiscounted and discounted life expectancies are rounded to the nearest ten thousandth. Costs are rounded to the nearest dollar and are presented in 2020 USD. Discounted values are discounted at 3% per year. ICERs are rounded to the nearest dollar and are calculated using unrounded discounted life expectancy and discounted costs. The cost-effective bNAb strategy was the strategy that offered the greatest increase in overall population life expectancy while still having an ICER less than the cost-effectiveness threshold when compared to the next best performing, non-dominated strategy. * Indicates the cost-effective strategy using a cost-effectiveness threshold of 20% GDP per capita. † Indicates the cost-effective strategy using a cost-effectiveness threshold of 50% GDP per capita.

**Supplementary Table 18.** One-way sensitivity analysis: infant oral prophylaxis efficacy (base case: 69% intrapartum efficacy and 71% postpartum efficacy)

| | Clinical outcomes | | | Lifetime efficacy and costs | | | |
|---|---|---|---|---|---|---|---|
| **Country/strategy** | **IU/IP cumulative HIV incidence (%)** | **Postnatal cumulative HIV incidence (%)** | **Total cumulative HIV incidence (%)** | **Undiscounted life expectancy (yrs)** | **Discounted life expectancy (yrs)** | **Discounted costs ($)** | **ICER ($/YLS)** |
| **Côte d'Ivoire [CET: ICER ≤ $465/YLS (20% GDP per capita), ICER ≤ $1163/YLS (50% GDP per capita)]** | | | | | | | |
| ***Low infant oral prophylaxis efficacy (intrapartum efficacy: 40%; postpartum efficacy: 58%)*** | | | | | | | |
| Standard-of-care | 3·7 | 5·1 | 8·5 | 60·128 | 26·373 | 15 | Reference |
| HR-HIVE – 1 dose | 3·4 | 4·9 | 8·1 | 60·132 | 26·374 | 15 | dominated |
| HR-HIVE – 2 doses | 3·4 | 4·8 | 7·9 | 60·133 | 26·375 | 15 | dominated |
| HIVE – 1 dose | 3·3 | 4·8 | 7·9 | 60·134 | 26·375 | 14 | dominated |
| HR-HIVE – Extended | 3·4 | 4·4 | 7·6 | 60·135 | 26·376 | 15 | dominated |
| HIVE – 2 doses | 3·3 | 4·4 | 7·5 | 60·137 | 26·376 | 14 | dominated |
| ALL – 1 dose | 3·1 | 4·7 | 7·6 | 60·139 | 26·377 | 28 | dominated |
| ALL – 2 doses | 3·1 | 4·2 | 7·1 | 60·139 | 26·378 | 43 | dominated |
| HIVE – Extended | 3·3 | 3·2 | 6·4 | 60·145 | 26·379 | 13 | cost-saving*·† |
| ALL – 1 dose plus HIVE - Extended | 3·1 | 3·1 | 6·1 | 60·150 | 26·381 | 26 | 6240 |
| ALL - Extended | 3·1 | 2·7 | 5·6 | 60·155 | 26·382 | 91 | 57 858 |
| ***High infant oral prophylaxis efficacy (intrapartum efficacy: 90%; postpartum efficacy: 80%)*** | | | | | | | |
| Standard-of-care | 3·6 | 4·9 | 8·3 | 60·130 | 26·374 | 14 | Reference |
| HR-HIVE – 1 dose | 3·3 | 4·8 | 7·9 | 60·133 | 26·375 | 15 | dominated |
| HR-HIVE – 2 doses | 3·3 | 4·7 | 7·7 | 60·135 | 26·375 | 15 | dominated |
| HIVE – 1 dose | 3·2 | 4·7 | 7·7 | 60·135 | 26·376 | 14 | dominated |
| HR-HIVE – Extended | 3·3 | 4·3 | 7·4 | 60·137 | 26·376 | 14 | dominated |
| HIVE – 2 doses | 3·2 | 4·3 | 7·3 | 60·138 | 26·377 | 14 | dominated |
| ALL – 1 dose | 3·0 | 4·6 | 7·4 | 60·140 | 26·378 | 27 | dominated |
| ALL – 2 doses | 3·0 | 4·1 | 6·9 | 60·141 | 26·378 | 43 | dominated |
| HIVE – Extended | 3·2 | 3·1 | 6·2 | 60·146 | 26·380 | 13 | cost-saving*·† |
| ALL – 1 dose plus HIVE - Extended | 3·0 | 3·1 | 5·9 | 60·151 | 26·382 | 26 | 6246 |
| ALL - Extended | 3·0 | 2·6 | 5·5 | 60·157 | 26·383 | 91 | 57 856 |
| **South Africa [CET: ICER ≤ $1131/YLS (20% GDP per capita), ICER ≤ $2828/YLS (50% GDP per capita)]** | | | | | | | |
| ***Low infant oral prophylaxis efficacy (intrapartum efficacy: 40%; postpartum efficacy: 58%)*** | | | | | | | |
| Standard-of-care | 2·4 | 2·2 | 4·5 | 68·909 | 28·494 | 114 | Reference |
| HR-HIVE – 1 dose | 2·1 | 2·1 | 4·1 | 68·970 | 28·515 | 116 | dominated |
| HR-HIVE – 2 doses | 2·1 | 2·1 | 4·0 | 68·977 | 28·517 | 116 | dominated |
| HR-HIVE – Extended | 2·1 | 2·0 | 4·0 | 68·985 | 28·520 | 115 | dominated |
| HIVE – 1 dose | 2·0 | 2·0 | 3·9 | 69·003 | 28·526 | 106 | dominated |
| HIVE – 2 doses | 2·0 | 1·8 | 3·7 | 69·037 | 28·538 | 105 | dominated |
| ALL – 1 dose | 1·8 | 1·8 | 3·5 | 69·055 | 28·545 | 117 | dominated |
| HIVE – Extended | 2·0 | 1·5 | 3·4 | 69·086 | 28·554 | 101 | cost-saving |
| ALL – 2 doses | 1·8 | 1·5 | 3·2 | 69·114 | 28·565 | 123 | dominated |
| ALL – 1 dose plus HIVE - Extended | 1·8 | 1·3 | 3·0 | 69·139 | 28·573 | 112 | 596 |
| ALL - Extended | 1·8 | 0·8 | 2·6 | 69·220 | 28·599 | 136 | 882*·† |

**Supplementary Table 18.** One-way sensitivity analysis: infant oral prophylaxis efficacy (base case: 69% intrapartum efficacy and 71% postpartum efficacy) (cont.)

| Country/strategy | Clinical outcomes: IU/IP cumulative HIV incidence (%) | Postnatal cumulative HIV incidence (%) | Total cumulative HIV incidence (%) | Lifetime efficacy and costs: Undiscounted life expectancy (yrs) | Discounted life expectancy (yrs) | Discounted costs ($) | ICER ($/YLS) |
|---|---|---|---|---|---|---|---|
| **South Africa [CET: ICER ≤ $1131/YLS (20% GDP per capita), ICER ≤ $2828/YLS (50% GDP per capita)]** | | | | | | | |
| ***High infant oral prophylaxis efficacy (intrapartum efficacy: 90%; postpartum efficacy: 80%)*** | | | | | | | |
| Standard-of-care | 2·3 | 2·2 | 4·3 | 68·934 | 28·503 | 110 | Reference |
| HR-HIVE – 1 dose | 2·0 | 2·1 | 4·0 | 68·986 | 28·521 | 114 | dominated |
| HR-HIVE – 2 doses | 2·0 | 2·0 | 3·9 | 68·993 | 28·523 | 114 | dominated |
| HR-HIVE – Extended | 2·0 | 2·0 | 3·9 | 69·002 | 28·526 | 113 | dominated |
| HIVE – 1 dose | 1·9 | 2·0 | 3·8 | 69·017 | 28·531 | 104 | dominated |
| HIVE – 2 doses | 1·9 | 1·8 | 3·6 | 69·051 | 28·543 | 103 | dominated |
| ALL – 1 dose | 1·8 | 1·8 | 3·4 | 69·069 | 28·550 | 115 | dominated |
| HIVE – Extended | 1·9 | 1·5 | 3·3 | 69·101 | 28·559 | 100 | cost-saving |
| ALL – 2 doses | 1·8 | 1·4 | 3·1 | 69·127 | 28·570 | 122 | dominated |
| ALL – 1 dose plus HIVE - Extended | 1·8 | 1·3 | 2·9 | 69·152 | 28·577 | 111 | 613 |
| ALL - Extended | 1·8 | 0·8 | 2·5 | 69·233 | 28·604 | 134 | 882*·† |
| **Zimbabwe [CET: ICER ≤ $243/YLS (20% GDP per capita), ICER ≤ $607/YLS (50% GDP per capita)]** | | | | | | | |
| ***Low infant oral prophylaxis efficacy (intrapartum efficacy: 40%; postpartum efficacy: 58%)*** | | | | | | | |
| Standard-of-care | 4·5 | 5·8 | 9·8 | 68·303 | 28·027 | 74 | Reference |
| HR-HIVE – 1 dose | 3·9 | 5·4 | 8·8 | 68·354 | 28·046 | 72 | dominated |
| HR-HIVE – 2 doses | 3·9 | 5·3 | 8·7 | 68·361 | 28·048 | 71 | dominated |
| HIVE – 1 dose | 3·8 | 5·2 | 8·6 | 68·369 | 28·051 | 68 | dominated |
| HR-HIVE – Extended | 3·9 | 5·0 | 8·4 | 68·375 | 28·053 | 70 | dominated |
| ALL – 1 dose | 3·6 | 4·9 | 8·1 | 68·389 | 28·058 | 81 | dominated |
| HIVE – 2 doses | 3·8 | 4·8 | 8·2 | 68·391 | 28·059 | 67 | dominated |
| ALL – 2 doses | 3·6 | 4·4 | 7·6 | 68·427 | 28·070 | 94 | dominated |
| HIVE – Extended | 3·8 | 3·7 | 7·1 | 68·446 | 28·076 | 63 | cost-saving*·† |
| ALL – 1 dose plus HIVE - Extended | 3·6 | 3·4 | 6·7 | 68·467 | 28·084 | 75 | 1715 |
| ALL - Extended | 3·6 | 2·6 | 5·9 | 68·506 | 28·097 | 135 | 4520 |
| ***High infant oral prophylaxis efficacy (intrapartum efficacy: 90%; postpartum efficacy: 80%)*** | | | | | | | |
| Standard-of-care | 4·2 | 5·5 | 9·2 | 68·334 | 28·038 | 70 | Reference |
| HR-HIVE – 1 dose | 3·6 | 5·3 | 8·5 | 68·374 | 28·053 | 70 | dominated |
| HR-HIVE – 2 doses | 3·6 | 5·1 | 8·3 | 68·382 | 28·056 | 69 | dominated |
| HIVE – 1 dose | 3·6 | 5·1 | 8·3 | 68·388 | 28·058 | 66 | dominated |
| HR-HIVE – Extended | 3·6 | 4·8 | 8·1 | 68·396 | 28·060 | 68 | dominated |
| ALL – 1 dose | 3·3 | 4·8 | 7·8 | 68·408 | 28·065 | 78 | dominated |
| HIVE – 2 doses | 3·6 | 4·7 | 7·9 | 68·410 | 28·066 | 65 | dominated |
| ALL – 2 doses | 3·3 | 4·3 | 7·3 | 68·446 | 28·077 | 92 | dominated |
| HIVE – Extended | 3·6 | 3·6 | 6·8 | 68·466 | 28·083 | 60 | cost-saving*·† |
| ALL – 1 dose plus HIVE - Extended | 3·3 | 3·3 | 6·3 | 68·486 | 28·091 | 73 | 1738 |
| ALL - Extended | 3·3 | 2·5 | 5·5 | 68·525 | 28·104 | 133 | 4516 |

*bNAb: broadly neutralizing antibody; IU/IP: intrauterine/intrapartum; yr: year; ICER: incremental cost-effectiveness ratio; YLS: years of life saved; CET: cost-effectiveness threshold; HR-HIVE: high-risk HIV-exposed infants; HIVE: all HIV-exposed infants; ALL: all live infants at birth.*

Pediatric HIV incidence is rounded to the nearest tenth of a percent. IU/IP HIV incidence is calculated based on the number of infants exposed to HIV at birth. Postnatal and total HIV incidence is calculated based on the number of infants ever exposed to HIV through 36 months of life. Undiscounted and discounted life expectancies are rounded to the nearest ten thousandth. Costs are rounded to the nearest dollar and are presented in 2020 USD. Discounted values are discounted at 3% per year. ICERs are rounded to the nearest dollar and are calculated using unrounded discounted life expectancy and discounted costs. The cost-effective bNAb strategy was the strategy that offered the greatest increase in overall population life expectancy while still having an ICER less than the cost-effectiveness threshold when compared to the next best performing, non-dominated strategy. * Indicates the cost-effective strategy using a cost-effectiveness threshold of 20% GDP per capita. † Indicates the cost-effective strategy using a cost-effectiveness threshold of 50% GDP per capita.

**Supplementary Table 19.** One-way sensitivity analysis: additional cost of identifying high-risk infants

| Country/strategy | Clinical outcomes | | | Lifetime efficacy and costs | | | |
|---|---|---|---|---|---|---|---|
| | IU/IP cumulative HIV incidence (%) | Postnatal cumulative HIV incidence (%) | Total cumulative HIV incidence (%) | Undiscounted life expectancy (yrs) | Discounted life expectancy (yrs) | Discounted costs ($) | ICER ($/YLS) |
| **Côte d'Ivoire [CET: ICER ≤ $465/YLS (20% GDP per capita), ICER ≤ $1163/YLS (50% GDP per capita)]** | | | | | | | |
| ***Cost of identifying high-risk infants: $0*** | | | | | | | |
| Standard-of-care | 3·6 | 5·0 | 8·4 | 60·129 | 26·373 | 15 | Reference |
| HR-HIVE – 1 dose | 3·3 | 4·8 | 8·0 | 60·133 | 26·375 | 14 | dominated |
| HR-HIVE – 2 doses | 3·3 | 4·7 | 7·8 | 60·134 | 26·375 | 14 | dominated |
| HIVE – 1 dose | 3·3 | 4·7 | 7·8 | 60·134 | 26·375 | 14 | dominated |
| HR-HIVE – Extended | 3·3 | 4·3 | 7·5 | 60·136 | 26·376 | 14 | dominated |
| HIVE – 2 doses | 3·3 | 4·3 | 7·4 | 60·138 | 26·377 | 14 | dominated |
| ALL – 1 dose | 3·0 | 4·6 | 7·5 | 60·139 | 26·377 | 27 | dominated |
| ALL – 2 doses | 3·0 | 4·2 | 7·0 | 60·140 | 26·378 | 43 | dominated |
| HIVE – Extended | 3·3 | 3·2 | 6·3 | 60·146 | 26·379 | 13 | cost-saving*·† |
| ALL – 1 dose plus HIVE - Extended | 3·0 | 3·1 | 6·0 | 60·150 | 26·382 | 26 | 6242 |
| ALL - Extended | 3·0 | 2·7 | 5·5 | 60·156 | 26·383 | 91 | 57 860 |
| ***Cost of identifying high-risk infants: $32*** | | | | | | | |
| Standard-of-care | 3·6 | 5·0 | 8·4 | 60·129 | 26·373 | 15 | Reference |
| HR-HIVE – 1 dose | 3·3 | 4·8 | 8·0 | 60·133 | 26·375 | 15 | dominated |
| HR-HIVE – 2 doses | 3·3 | 4·7 | 7·8 | 60·134 | 26·375 | 15 | dominated |
| HIVE – 1 dose | 3·3 | 4·7 | 7·8 | 60·134 | 26·375 | 14 | dominated |
| HR-HIVE – Extended | 3·3 | 4·3 | 7·5 | 60·136 | 26·376 | 14 | dominated |
| HIVE – 2 doses | 3·3 | 4·3 | 7·4 | 60·138 | 26·377 | 14 | dominated |
| ALL – 1 dose | 3·0 | 4·6 | 7·5 | 60·139 | 26·377 | 27 | dominated |
| ALL – 2 doses | 3·0 | 4·2 | 7·0 | 60·140 | 26·378 | 43 | dominated |
| HIVE – Extended | 3·3 | 3·2 | 6·3 | 60·146 | 26·379 | 13 | cost-saving*·† |
| ALL – 1 dose plus HIVE - Extended | 3·0 | 3·1 | 6·0 | 60·150 | 26·382 | 26 | 6242 |
| ALL - Extended | 3·0 | 2·7 | 5·5 | 60·156 | 26·383 | 91 | 57 860 |
| **South Africa [CET: ICER ≤ $1131/YLS (20% GDP per capita), ICER ≤ $2828/YLS (50% GDP per capita)]** | | | | | | | |
| ***Cost of identifying high-risk infants: $0*** | | | | | | | |
| Standard-of-care | 2·3 | 2·2 | 4·4 | 68·924 | 28·499 | 112 | Reference |
| HR-HIVE – 1 dose | 2·1 | 2·1 | 4·0 | 68·979 | 28·518 | 105 | dominated |
| HR-HIVE – 2 doses | 2·1 | 2·1 | 4·0 | 68·986 | 28·521 | 105 | dominated |
| HR-HIVE – Extended | 2·1 | 2·0 | 3·9 | 68·995 | 28·523 | 104 | dominated |
| HIVE – 1 dose | 2·0 | 2·0 | 3·8 | 69·011 | 28·529 | 105 | dominated |
| HIVE – 2 doses | 2·0 | 1·8 | 3·6 | 69·045 | 28·541 | 104 | dominated |
| ALL – 1 dose | 1·8 | 1·8 | 3·5 | 69·063 | 28·548 | 116 | dominated |
| HIVE – Extended | 2·0 | 1·5 | 3·3 | 69·095 | 28·557 | 100 | cost-saving |
| ALL – 2 doses | 1·8 | 1·5 | 3·2 | 69·122 | 28·568 | 122 | dominated |
| ALL – 1 dose plus HIVE - Extended | 1·8 | 1·3 | 3·0 | 69·147 | 28·575 | 111 | 606 |
| ALL - Extended | 1·8 | 0·8 | 2·5 | 69·228 | 28·602 | 135 | 882*·† |

**Supplementary Table 19.** One-way sensitivity analysis: additional cost of identifying high-risk infants (cont.)

| | Clinical outcomes | | | Lifetime efficacy and costs | | | |
|---|---|---|---|---|---|---|---|
| Country/strategy | IU/IP cumulative HIV incidence (%) | Postnatal cumulative HIV incidence (%) | Total cumulative HIV incidence (%) | Undiscounted life expectancy (yrs) | Discounted life expectancy (yrs) | Discounted costs ($) | ICER ($/YLS) |
| **South Africa [CET: ICER ≤ $1131/YLS (20% GDP per capita), ICER ≤ $2828/YLS (50% GDP per capita)]** | | | | | | | |
| ***Cost of identifying high-risk infants: $32*** | | | | | | | |
| Standard-of-care | 2·3 | 2·2 | 4·4 | 68·924 | 28·499 | 112 | Reference |
| HR-HIVE – 1 dose | 2·1 | 2·1 | 4·0 | 68·979 | 28·518 | 115 | dominated |
| HR-HIVE – 2 doses | 2·1 | 2·1 | 4·0 | 68·986 | 28·521 | 115 | dominated |
| HR-HIVE – Extended | 2·1 | 2·0 | 3·9 | 68·995 | 28·523 | 114 | dominated |
| HIVE – 1 dose | 2·0 | 2·0 | 3·8 | 69·011 | 28·529 | 105 | dominated |
| HIVE – 2 doses | 2·0 | 1·8 | 3·6 | 69·045 | 28·541 | 104 | dominated |
| ALL – 1 dose | 1·8 | 1·8 | 3·5 | 69·063 | 28·548 | 116 | dominated |
| HIVE – Extended | 2·0 | 1·5 | 3·3 | 69·095 | 28·557 | 100 | cost-saving |
| ALL – 2 doses | 1·8 | 1·5 | 3·2 | 69·122 | 28·568 | 122 | dominated |
| ALL – 1 dose plus HIVE - Extended | 1·8 | 1·3 | 3·0 | 69·147 | 28·575 | 111 | 606 |
| ALL - Extended | 1·8 | 0·8 | 2·5 | 69·228 | 28·602 | 135 | 882*·† |
| **Zimbabwe [CET: ICER ≤ $243/YLS (20% GDP per capita), ICER ≤ $607/YLS (50% GDP per capita)]** | | | | | | | |
| ***Cost of identifying high-risk infants: $0*** | | | | | | | |
| Standard-of-care | 4·3 | 5·6 | 9·5 | 68·321 | 28·034 | 72 | Reference |
| HR-HIVE – 1 dose | 3·7 | 5·3 | 8·6 | 68·366 | 28·050 | 67 | dominated |
| HR-HIVE – 2 doses | 3·7 | 5·2 | 8·5 | 68·373 | 28·053 | 66 | dominated |
| HIVE – 1 dose | 3·7 | 5·2 | 8·4 | 68·380 | 28·055 | 67 | dominated |
| HR-HIVE – Extended | 3·7 | 4·9 | 8·2 | 68·387 | 28·057 | 65 | dominated |
| ALL – 1 dose | 3·4 | 4·9 | 7·9 | 68·400 | 28·062 | 79 | dominated |
| HIVE – 2 doses | 3·7 | 4·8 | 8·0 | 68·402 | 28·063 | 66 | dominated |
| ALL – 2 doses | 3·4 | 4·3 | 7·4 | 68·438 | 28·074 | 93 | dominated |
| HIVE – Extended | 3·7 | 3·6 | 6·9 | 68·458 | 28·081 | 61 | cost-saving*·† |
| ALL – 1 dose plus HIVE - Extended | 3·4 | 3·4 | 6·5 | 68·478 | 28·088 | 74 | 1731 |
| ALL - Extended | 3·4 | 2·6 | 5·7 | 68·517 | 28·101 | 134 | 4516 |
| ***Cost of identifying high-risk infants: $32*** | | | | | | | |
| Standard-of-care | 4·3 | 5·6 | 9·5 | 68·321 | 28·034 | 72 | Reference |
| HR-HIVE – 1 dose | 3·7 | 5·3 | 8·6 | 68·366 | 28·050 | 71 | dominated |
| HR-HIVE – 2 doses | 3·7 | 5·2 | 8·5 | 68·373 | 28·053 | 70 | dominated |
| HIVE – 1 dose | 3·7 | 5·2 | 8·4 | 68·380 | 28·055 | 67 | dominated |
| HR-HIVE – Extended | 3·7 | 4·9 | 8·2 | 68·387 | 28·057 | 69 | dominated |
| ALL – 1 dose | 3·4 | 4·9 | 7·9 | 68·400 | 28·062 | 79 | dominated |
| HIVE – 2 doses | 3·7 | 4·8 | 8·0 | 68·402 | 28·063 | 66 | dominated |
| ALL – 2 doses | 3·4 | 4·3 | 7·4 | 68·438 | 28·074 | 93 | dominated |
| HIVE – Extended | 3·7 | 3·6 | 6·9 | 68·458 | 28·081 | 61 | cost-saving*·† |
| ALL – 1 dose plus HIVE - Extended | 3·4 | 3·4 | 6·5 | 68·478 | 28·088 | 74 | 1731 |
| ALL - Extended | 3·4 | 2·6 | 5·7 | 68·517 | 28·101 | 134 | 4516 |

*bNAb: broadly neutralizing antibody; IU/IP: intrauterine/intrapartum; yr: year; ICER: incremental cost-effectiveness ratio; YLS: years of life saved; CET: cost-effectiveness threshold; HR-HIVE: high-risk HIV-exposed infants; HIVE: all HIV-exposed infants; ALL: all live infants at birth.*

The cost of ascertaining an infant's HIV exposure risk status includes a maternal viral load test ($24·05), result return ($3·48), and personnel/overhead costs ($4·13).[26,27,149,151] Pediatric HIV incidence is rounded to the nearest tenth of a percent. IU/IP HIV incidence is calculated based on the number of infants exposed to HIV at birth. Postnatal and total HIV incidence is calculated based on the number of infants ever exposed to HIV through 36 months of life. Undiscounted and discounted life expectancies are rounded to the nearest ten thousandth. Costs are rounded to the nearest dollar and are presented in 2020 USD. Discounted values are discounted at 3% per year. ICERs are rounded to the nearest dollar and are calculated using unrounded discounted life expectancy and discounted costs. The cost-effective bNAb strategy was the strategy that offered the greatest increase in overall population life expectancy while still having an ICER less than the cost-effectiveness threshold when compared to the next best performing, non-dominated strategy. * Indicates the cost-effective strategy using a cost-effectiveness threshold of 20% GDP per capita. † Indicates the cost-effective strategy using a cost-effectiveness threshold of 50% GDP per capita.

**Supplementary Table 20.** One-way sensitivity analysis: maternal knowledge of acute HIV infection

| Country/strategy | Clinical outcomes | | | Lifetime efficacy and costs | | | |
|---|---|---|---|---|---|---|---|
| | IU/IP cumulative HIV incidence (%) | Postnatal cumulative HIV incidence (%) | Total cumulative HIV incidence (%) | Undiscounted life expectancy (yrs) | Discounted life expectancy (yrs) | Discounted costs ($) | ICER ($/YLS) |
| **Côte d'Ivoire [CET: ICER ≤ $465/YLS (20% GDP per capita), ICER ≤ $1163/YLS (50% GDP per capita)]** | | | | | | | |
| ***Low maternal knowledge of acute HIV infection (in pregnancy: 25%; retesting postpartum: 0%/month)*** | | | | | | | |
| Standard-of-care | 3·7 | 5·0 | 8·4 | 60·128 | 26·373 | 15 | Reference |
| HR-HIVE – 1 dose | 3·4 | 4·9 | 8·1 | 60·132 | 26·374 | 15 | dominated |
| HR-HIVE – 2 doses | 3·4 | 4·8 | 7·9 | 60·133 | 26·375 | 15 | dominated |
| HIVE – 1 dose | 3·3 | 4·8 | 7·9 | 60·133 | 26·375 | 14 | dominated |
| HR-HIVE – Extended | 3·4 | 4·4 | 7·6 | 60·135 | 26·375 | 14 | dominated |
| HIVE – 2 doses | 3·3 | 4·4 | 7·5 | 60·136 | 26·376 | 14 | dominated |
| ALL – 1 dose | 3·0 | 4·7 | 7·6 | 60·138 | 26·377 | 27 | dominated |
| ALL – 2 doses | 3·0 | 4·2 | 7·1 | 60·139 | 26·378 | 44 | dominated |
| HIVE – Extended | 3·3 | 3·2 | 6·4 | 60·144 | 26·379 | 13 | cost-saving*·† |
| ALL – 1 dose plus HIVE - Extended | 3·0 | 3·2 | 6·1 | 60·149 | 26·381 | 26 | 6012 |
| ALL - Extended | 3·0 | 2·7 | 5·6 | 60·155 | 26·382 | 91 | 55 092 |
| ***High maternal knowledge of acute HIV infection (in pregnancy: 95%; retesting postpartum: 15%/month)*** | | | | | | | |
| Standard-of-care | 3·6 | 4·6 | 8·0 | 60·133 | 26·375 | 14 | Reference |
| HR-HIVE – 1 dose | 3·3 | 4·5 | 7·6 | 60·136 | 26·376 | 14 | dominated |
| HR-HIVE – 2 doses | 3·3 | 4·4 | 7·5 | 60·137 | 26·377 | 14 | dominated |
| HIVE – 1 dose | 3·2 | 4·4 | 7·4 | 60·138 | 26·377 | 14 | dominated |
| HR-HIVE – Extended | 3·3 | 4·0 | 7·1 | 60·140 | 26·377 | 14 | dominated |
| HIVE – 2 doses | 3·2 | 4·0 | 7·0 | 60·141 | 26·378 | 13 | dominated |
| ALL – 1 dose | 3·0 | 4·3 | 7·1 | 60·143 | 26·379 | 27 | dominated |
| ALL – 2 doses | 3·0 | 3·8 | 6·7 | 60·143 | 26·379 | 42 | dominated |
| HIVE – Extended | 3·2 | 2·8 | 5·9 | 60·149 | 26·381 | 12 | cost-saving*·† |
| ALL – 1 dose plus HIVE - Extended | 3·0 | 2·8 | 5·6 | 60·154 | 26·383 | 26 | 6779 |
| ALL - Extended | 3·0 | 2·4 | 5·3 | 60·159 | 26·384 | 91 | 79 100 |
| **South Africa [CET: ICER ≤ $1131/YLS (20% GDP per capita), ICER ≤ $2828/YLS (50% GDP per capita)]** | | | | | | | |
| ***Low maternal knowledge of acute HIV infection (in pregnancy: 25%; retesting postpartum: 0%/month)*** | | | | | | | |
| Standard-of-care | 2·7 | 2·2 | 4·7 | 68·851 | 28·472 | 118 | Reference |
| HR-HIVE – 1 dose | 2·4 | 2·1 | 4·3 | 68·905 | 28·491 | 121 | dominated |
| HR-HIVE – 2 doses | 2·4 | 2·1 | 4·3 | 68·911 | 28·493 | 121 | dominated |
| HR-HIVE – Extended | 2·4 | 2·0 | 4·3 | 68·919 | 28·495 | 120 | dominated |
| HIVE – 1 dose | 2·3 | 2·0 | 4·1 | 68·937 | 28·502 | 111 | dominated |
| HIVE – 2 doses | 2·3 | 1·8 | 4·0 | 68·970 | 28·513 | 110 | dominated |
| ALL – 1 dose | 2·1 | 1·8 | 3·7 | 69·000 | 28·524 | 120 | dominated |
| HIVE – Extended | 2·3 | 1·5 | 3·7 | 69·019 | 28·529 | 107 | cost-saving |
| ALL – 2 doses | 2·1 | 1·5 | 3·4 | 69·060 | 28·545 | 127 | dominated |
| ALL – 1 dose plus HIVE - Extended | 2·1 | 1·3 | 3·2 | 69·082 | 28·551 | 116 | 405 |
| ALL - Extended | 2·1 | 0·8 | 2·8 | 69·164 | 28·578 | 139 | 875*·† |

**Supplementary Table 20.** One-way sensitivity analysis: HIV knowledge of acute infection (cont.)

| | Clinical outcomes | | | Lifetime efficacy and costs | | | |
|---|---|---|---|---|---|---|---|
| Country/strategy | IU/IP cumulative HIV incidence (%) | Postnatal cumulative HIV incidence (%) | Total cumulative HIV incidence (%) | Undiscounted life expectancy (yrs) | Discounted life expectancy (yrs) | Discounted costs ($) | ICER ($/YLS) |
| **South Africa [CET: ICER ≤ $1131/YLS (20% GDP per capita), ICER ≤ $2828/YLS (50% GDP per capita)]** | | | | | | | |
| ***High maternal knowledge of acute HIV infection (in pregnancy: 95%; retesting postpartum: 15%/month)*** | | | | | | | |
| Standard-of-care | 1·9 | 2·0 | 3·8 | 69·051 | 28·546 | 101 | Reference |
| HR-HIVE – 1 dose | 1·6 | 1·9 | 3·4 | 69·109 | 28·566 | 104 | dominated |
| HR-HIVE – 2 doses | 1·6 | 1·8 | 3·4 | 69·117 | 28·568 | 104 | dominated |
| HR-HIVE – Extended | 1·6 | 1·8 | 3·3 | 69·127 | 28·571 | 104 | dominated |
| HIVE – 1 dose | 1·5 | 1·8 | 3·2 | 69·140 | 28·577 | 94 | dominated |
| ALL – 1 dose | 1·5 | 1·7 | 3·1 | 69·161 | 28·584 | 109 | dominated |
| HIVE – 2 doses | 1·5 | 1·6 | 3·0 | 69·175 | 28·589 | 93 | dominated |
| ALL – 2 doses | 1·5 | 1·4 | 2·8 | 69·217 | 28·603 | 115 | dominated |
| HIVE – Extended | 1·5 | 1·3 | 2·7 | 69·226 | 28·605 | 90 | cost-saving* |
| ALL – 1 dose plus HIVE - Extended | 1·5 | 1·1 | 2·6 | 69·247 | 28·612 | 104 | dominated |
| ALL - Extended | 1·5 | 0·7 | 2·1 | 69·320 | 28·636 | 128 | 1221† |
| **Zimbabwe [CET: ICER ≤ $243/YLS (20% GDP per capita), ICER ≤ $607/YLS (50% GDP per capita)]** | | | | | | | |
| ***Low maternal knowledge of acute HIV infection (in pregnancy: 25%; retesting postpartum: 0%/month)*** | | | | | | | |
| Standard-of-care | 4·9 | 6·6 | 10·9 | 68·213 | 27·994 | 79 | Reference |
| HR-HIVE – 1 dose | 4·5 | 6·4 | 10·3 | 68·244 | 28·005 | 79 | dominated |
| HR-HIVE – 2 doses | 4·5 | 6·3 | 10·2 | 68·249 | 28·007 | 79 | dominated |
| HIVE – 1 dose | 4·4 | 6·3 | 10·1 | 68·258 | 28·010 | 76 | dominated |
| HR-HIVE – Extended | 4·5 | 6·1 | 10·0 | 68·260 | 28·010 | 78 | dominated |
| HIVE – 2 doses | 4·4 | 5·9 | 9·7 | 68·278 | 28·017 | 75 | dominated |
| ALL – 1 dose | 3·9 | 5·6 | 9·1 | 68·310 | 28·029 | 85 | dominated |
| HIVE – Extended | 4·4 | 4·8 | 8·7 | 68·330 | 28·034 | 70 | cost-saving* |
| ALL – 2 doses | 3·9 | 5·0 | 8·5 | 68·351 | 28·042 | 99 | dominated |
| ALL – 1 dose plus HIVE - Extended | 3·9 | 4·2 | 7·7 | 68·383 | 28·053 | 80 | 520† |
| ALL - Extended | 3·9 | 3·1 | 6·6 | 68·438 | 28·072 | 138 | 3081 |
| ***High maternal knowledge of acute HIV infection (in pregnancy: 95%; retesting postpartum: 15%/month)*** | | | | | | | |
| Standard-of-care | 4·0 | 5·0 | 8·6 | 68·383 | 28·056 | 67 | Reference |
| HR-HIVE – 1 dose | 3·4 | 4·6 | 7·6 | 68·436 | 28·075 | 65 | dominated |
| HR-HIVE – 2 doses | 3·4 | 4·5 | 7·5 | 68·444 | 28·078 | 64 | dominated |
| HIVE – 1 dose | 3·3 | 4·5 | 7·4 | 68·449 | 28·080 | 61 | dominated |
| ALL – 1 dose | 3·2 | 4·4 | 7·2 | 68·454 | 28·082 | 75 | dominated |
| HR-HIVE – Extended | 3·4 | 4·2 | 7·2 | 68·460 | 28·083 | 63 | dominated |
| HIVE – 2 doses | 3·3 | 4·1 | 7·0 | 68·473 | 28·088 | 60 | dominated |
| ALL – 2 doses | 3·2 | 3·9 | 6·7 | 68·490 | 28·093 | 89 | dominated |
| HIVE – Extended | 3·3 | 2·9 | 5·9 | 68·530 | 28·107 | 55 | cost-saving*·† |
| ALL – 1 dose plus HIVE - Extended | 3·2 | 2·8 | 5·7 | 68·534 | 28·108 | 70 | dominated |
| ALL - Extended | 3·2 | 2·3 | 5·2 | 68·561 | 28·117 | 130 | 7013 |

*bNAb: broadly neutralizing antibody; IU/IP: intrauterine/intrapartum; yr: year; ICER: incremental cost-effectiveness ratio; YLS: years of life saved; CET: cost-effectiveness threshold; HR-HIVE: high-risk HIV-exposed infants; HIVE: all HIV-exposed infants; ALL: all live infants at birth.*

Pediatric HIV incidence is rounded to the nearest tenth of a percent. IU/IP HIV incidence is calculated based on the number of infants exposed to HIV at birth. Postnatal and total HIV incidence is calculated based on the number of infants ever exposed to HIV through 36 months of life. Undiscounted and discounted life expectancies are rounded to the nearest ten thousandth. Costs are rounded to the nearest dollar and are presented in 2020 USD. Discounted values are discounted at 3% per year. ICERs are rounded to the nearest dollar and are calculated using unrounded discounted life expectancy and discounted costs. The cost-effective bNAb strategy was the strategy that offered the greatest increase in overall population life expectancy while still having an ICER less than the cost-effectiveness threshold when compared to the next best performing, non-dominated strategy. * Indicates the cost-effective strategy using a cost-effectiveness threshold of 20% GDP per capita. † Indicates the cost-effective strategy using a cost-effectiveness threshold of 50% GDP per capita.

**Supplementary Table 21.** One-way sensitivity analysis: maternal knowledge of chronic HIV infection

| | Clinical outcomes | | | Lifetime efficacy and costs | | | |
|---|---|---|---|---|---|---|---|
| **Country/strategy** | **IU/IP cumulative HIV incidence (%)** | **Postnatal cumulative HIV incidence (%)** | **Total cumulative HIV incidence (%)** | **Undiscounted life expectancy (yrs)** | **Discounted life expectancy (yrs)** | **Discounted costs ($)** | **ICER ($/YLS)** |
| **Côte d'Ivoire [CET: ICER ≤ $465/YLS (20% GDP per capita), ICER ≤ $1163/YLS (50% GDP per capita)]** | | | | | | | |
| ***Low maternal knowledge of chronic HIV infection: 56%*** | | | | | | | |
| Standard-of-care | 11·0 | 8·1 | 18·9 | 60·020 | 26·329 | 27 | Reference |
| HR-HIVE – 1 dose | 10·8 | 8·1 | 18·6 | 60·022 | 26·329 | 27 | dominated |
| HR-HIVE – 2 doses | 10·8 | 8·0 | 18·5 | 60·023 | 26·330 | 27 | dominated |
| HIVE – 1 dose | 10·7 | 8·0 | 18·5 | 60·023 | 26·330 | 27 | dominated |
| HR-HIVE – Extended | 10·8 | 7·7 | 18·3 | 60·025 | 26·330 | 27 | dominated |
| HIVE – 2 doses | 10·7 | 7·7 | 18·3 | 60·025 | 26·331 | 27 | dominated |
| HIVE – Extended | 10·7 | 7·0 | 17·5 | 60·030 | 26·332 | 26 | cost-saving*·† |
| ALL – 1 dose | 9·0 | 7·6 | 16·5 | 60·041 | 26·337 | 39 | dominated |
| ALL – 2 doses | 9·0 | 6·9 | 15·7 | 60·044 | 26·339 | 54 | dominated |
| ALL – 1 dose plus HIVE - Extended | 9·0 | 6·7 | 15·5 | 60·048 | 26·340 | 38 | 1587 |
| ALL - Extended | 9·0 | 4·5 | 13·4 | 60·065 | 26·345 | 100 | 11 429 |
| ***High maternal knowledge of chronic HIV infection: 100%*** | | | | | | | |
| Standard-of-care | 2·4 | 4·2 | 6·6 | 60·149 | 26·381 | 12 | Reference |
| HR-HIVE – 1 dose | 2·1 | 4·1 | 6·2 | 60·153 | 26·383 | 13 | dominated |
| HR-HIVE – 2 doses | 2·1 | 4·0 | 6·0 | 60·154 | 26·383 | 13 | dominated |
| HIVE – 1 dose | 2·0 | 4·0 | 6·0 | 60·155 | 26·384 | 12 | dominated |
| HR-HIVE – Extended | 2·1 | 3·6 | 5·7 | 60·157 | 26·384 | 12 | dominated |
| ALL – 1 dose | 2·0 | 4·0 | 5·9 | 60·157 | 26·385 | 25 | dominated |
| HIVE – 2 doses | 2·0 | 3·6 | 5·6 | 60·158 | 26·385 | 12 | dominated |
| ALL – 2 doses | 2·0 | 3·6 | 5·5 | 60·158 | 26·385 | 41 | dominated |
| HIVE – Extended | 2·0 | 2·4 | 4·4 | 60·166 | 26·388 | 10 | cost-saving*·† |
| ALL – 1 dose plus HIVE - Extended | 2·0 | 2·4 | 4·3 | 60·169 | 26·389 | 24 | 11 617 |
| ALL - Extended | 2·0 | 2·2 | 4·2 | 60·173 | 26·390 | 89 | 197 506 |
| **South Africa [CET: ICER ≤ $1131/YLS (20% GDP per capita), ICER ≤ $2828/YLS (50% GDP per capita)]** | | | | | | | |
| ***Low maternal knowledge of chronic HIV infection: 75%*** | | | | | | | |
| Standard-of-care | 6·9 | 3·3 | 9·8 | 67·760 | 28·071 | 207 | Reference |
| HR-HIVE – 1 dose | 6·7 | 3·2 | 9·5 | 67·807 | 28·087 | 209 | dominated |
| HR-HIVE – 2 doses | 6·7 | 3·1 | 9·5 | 67·812 | 28·089 | 209 | dominated |
| HR-HIVE – Extended | 6·7 | 3·1 | 9·4 | 67·819 | 28·091 | 208 | dominated |
| HIVE – 1 dose | 6·6 | 3·1 | 9·3 | 67·831 | 28·095 | 201 | dominated |
| HIVE – 2 doses | 6·6 | 2·9 | 9·2 | 67·857 | 28·104 | 200 | dominated |
| HIVE – Extended | 6·6 | 2·7 | 8·9 | 67·895 | 28·117 | 198 | dominated |
| ALL – 1 dose | 5·3 | 2·7 | 7·7 | 68·064 | 28·178 | 193 | dominated |
| ALL – 1 dose plus HIVE - Extended | 5·3 | 2·3 | 7·3 | 68·128 | 28·199 | 189 | cost-saving |
| ALL – 2 doses | 5·3 | 2·2 | 7·2 | 68·149 | 28·207 | 196 | dominated |
| ALL - Extended | 5·3 | 1·2 | 6·2 | 68·302 | 28·257 | 203 | 239*·† |

**Supplementary Table 21.** One-way sensitivity analysis: maternal knowledge of chronic HIV infection (cont.)

| | Clinical outcomes | | | Lifetime efficacy and costs | | | |
|---|---|---|---|---|---|---|---|
| **Country/strategy** | **IU/IP cumulative HIV incidence (%)** | **Postnatal cumulative HIV incidence (%)** | **Total cumulative HIV incidence (%)** | **Undiscounted life expectancy (yrs)** | **Discounted life expectancy (yrs)** | **Discounted costs ($)** | **ICER ($/YLS)** |
| **South Africa [CET: ICER ≤ $1131/YLS (20% GDP per capita), ICER ≤ $2828/YLS (50% GDP per capita)]** | | | | | | | |
| ***High maternal knowledge of chronic HIV infection: 100%*** | | | | | | | |
| Standard-of-care | 2·3 | 2·1 | 4·3 | 68·946 | 28·507 | 110 | Reference |
| HR-HIVE – 1 dose | 2·0 | 2·0 | 3·9 | 69·000 | 28·526 | 114 | dominated |
| HR-HIVE – 2 doses | 2·0 | 2·0 | 3·9 | 69·007 | 28·528 | 113 | dominated |
| HR-HIVE – Extended | 2·0 | 1·9 | 3·8 | 69·016 | 28·531 | 113 | dominated |
| HIVE – 1 dose | 1·9 | 1·9 | 3·7 | 69·034 | 28·538 | 103 | dominated |
| HIVE – 2 doses | 1·9 | 1·7 | 3·5 | 69·068 | 28·549 | 102 | dominated |
| ALL – 1 dose | 1·7 | 1·7 | 3·4 | 69·082 | 28·555 | 115 | dominated |
| HIVE – Extended | 1·9 | 1·4 | 3·2 | 69·118 | 28·566 | 99 | cost-saving |
| ALL – 2 doses | 1·7 | 1·4 | 3·1 | 69·140 | 28·575 | 121 | dominated |
| ALL – 1 dose plus HIVE - Extended | 1·7 | 1·2 | 2·9 | 69·166 | 28·583 | 110 | 674 |
| ALL - Extended | 1·7 | 0·8 | 2·4 | 69·245 | 28·608 | 134 | 909*·† |
| **Zimbabwe [CET: ICER ≤ $243/YLS (20% GDP per capita), ICER ≤ $607/YLS (50% GDP per capita)]** | | | | | | | |
| ***Low maternal knowledge of chronic HIV infection: 80%*** | | | | | | | |
| Standard-of-care | 7·3 | 6·6 | 13·4 | 68·047 | 27·933 | 94 | Reference |
| HR-HIVE – 1 dose | 6·8 | 6·3 | 12·6 | 68·088 | 27·947 | 93 | dominated |
| HR-HIVE – 2 doses | 6·8 | 6·2 | 12·5 | 68·094 | 27·950 | 93 | dominated |
| HIVE – 1 dose | 6·7 | 6·2 | 12·5 | 68·099 | 27·951 | 90 | dominated |
| HR-HIVE – Extended | 6·8 | 6·0 | 12·3 | 68·106 | 27·954 | 92 | dominated |
| HIVE – 2 doses | 6·7 | 5·9 | 12·1 | 68·117 | 27·958 | 89 | dominated |
| HIVE – Extended | 6·7 | 4·9 | 11·2 | 68·164 | 27·973 | 85 | cost-saving* |
| ALL – 1 dose | 5·8 | 5·8 | 11·2 | 68·159 | 27·973 | 99 | dominated |
| ALL – 2 doses | 5·8 | 5·1 | 10·5 | 68·203 | 27·987 | 112 | dominated |
| ALL – 1 dose plus HIVE - Extended | 5·8 | 4·6 | 9·9 | 68·224 | 27·994 | 94 | 410† |
| ALL - Extended | 5·8 | 3·1 | 8·5 | 68·298 | 28·019 | 150 | 2238 |
| ***High maternal knowledge of chronic HIV infection: 100%*** | | | | | | | |
| Standard-of-care | 4·0 | 5·3 | 9·1 | 68·348 | 28·044 | 69 | Reference |
| HR-HIVE – 1 dose | 3·4 | 5·0 | 8·2 | 68·397 | 28·062 | 68 | dominated |
| HR-HIVE – 2 doses | 3·4 | 4·9 | 8·1 | 68·404 | 28·064 | 68 | dominated |
| HIVE – 1 dose | 3·4 | 4·9 | 8·0 | 68·408 | 28·065 | 64 | dominated |
| HR-HIVE – Extended | 3·4 | 4·6 | 7·8 | 68·418 | 28·069 | 67 | dominated |
| ALL – 1 dose | 3·2 | 4·6 | 7·6 | 68·424 | 28·071 | 77 | dominated |
| HIVE – 2 doses | 3·4 | 4·5 | 7·6 | 68·430 | 28·073 | 64 | dominated |
| ALL – 2 doses | 3·2 | 4·1 | 7·1 | 68·462 | 28·083 | 91 | dominated |
| HIVE – Extended | 3·4 | 3·4 | 6·5 | 68·487 | 28·091 | 59 | cost-saving*·† |
| ALL – 1 dose plus HIVE - Extended | 3·2 | 3·1 | 6·1 | 68·503 | 28·097 | 72 | 2215 |
| ALL - Extended | 3·2 | 2·4 | 5·4 | 68·539 | 28·109 | 132 | 4077 |

*bNAb: broadly neutralizing antibody; IU/IP: intrauterine/intrapartum; yr: year; ICER: incremental cost-effectiveness ratio; YLS: years of life saved; CET: cost-effectiveness threshold; HR-HIVE: high-risk HIV-exposed infants; HIVE: all HIV-exposed infants; ALL: all live infants at birth.*

Pediatric HIV incidence is rounded to the nearest tenth of a percent. IU/IP HIV incidence is calculated based on the number of infants exposed to HIV at birth. Postnatal and total HIV incidence is calculated based on the number of infants ever exposed to HIV through 36 months of life. Undiscounted and discounted life expectancies are rounded to the nearest ten thousandth. Costs are rounded to the nearest dollar and are presented in 2020 USD. Discounted values are discounted at 3% per year. ICERs are rounded to the nearest dollar and are calculated using unrounded discounted life expectancy and discounted costs. The cost-effective bNAb strategy was the strategy that offered the greatest increase in overall population life expectancy while still having an ICER less than the cost-effectiveness threshold when compared to the next best performing, non-dominated strategy. *Indicates the cost-effective strategy using a cost-effectiveness threshold of 20% GDP per capita. †Indicates the cost-effective strategy using a cost-effectiveness threshold of 50% GDP per capita.

**Supplementary Table 22.** One-way sensitivity analysis: postpartum maternal retention in care

| Country/strategy | Clinical outcomes | | | Lifetime efficacy and costs | | | |
|---|---|---|---|---|---|---|---|
| | IU/IP cumulative HIV incidence (%) | Postnatal cumulative HIV incidence (%) | Total cumulative HIV incidence (%) | Undiscounted life expectancy (yrs) | Discounted life expectancy (yrs) | Discounted costs ($) | ICER ($/YLS) |
| **Côte d'Ivoire [CET: ICER ≤ $465/YLS (20% GDP per capita), ICER ≤ $1163/YLS (50% GDP per capita)]** | | | | | | | |
| ***Low postpartum maternal retention in care (50% of mothers with known HIV status in care at 6+ months)*** | | | | | | | |
| Standard-of-care | 3·6 | 7·1 | 10·4 | 60·114 | 26·368 | 18 | Reference |
| HR-HIVE – 1 dose | 3·3 | 7·0 | 10·0 | 60·118 | 26·369 | 19 | dominated |
| HR-HIVE – 2 doses | 3·3 | 6·8 | 9·8 | 60·119 | 26·370 | 18 | dominated |
| HIVE – 1 dose | 3·3 | 6·8 | 9·8 | 60·120 | 26·370 | 18 | dominated |
| HR-HIVE – Extended | 3·3 | 6·2 | 9·2 | 60·123 | 26·371 | 18 | dominated |
| HIVE – 2 doses | 3·3 | 6·2 | 9·2 | 60·125 | 26·372 | 17 | dominated |
| ALL – 1 dose | 3·0 | 6·8 | 9·5 | 60·125 | 26·372 | 31 | dominated |
| ALL – 2 doses | 3·0 | 6·0 | 8·8 | 60·127 | 26·373 | 47 | dominated |
| HIVE – Extended | 3·3 | 4·3 | 7·4 | 60·138 | 26·377 | 15 | cost-saving*·† |
| ALL – 1 dose plus HIVE - Extended | 3·0 | 4·2 | 7·1 | 60·143 | 26·379 | 28 | 6245 |
| ALL - Extended | 3·0 | 3·8 | 6·7 | 60·148 | 26·380 | 93 | 57 575 |
| ***High postpartum maternal retention in care (100% of mothers with known HIV status in care at 6+ months)*** | | | | | | | |
| Standard-of-care | 3·6 | 4·1 | 7·5 | 60·135 | 26·375 | 13 | Reference |
| HR-HIVE – 1 dose | 3·3 | 4·0 | 7·1 | 60·139 | 26·377 | 13 | dominated |
| HR-HIVE – 2 doses | 3·3 | 3·9 | 7·0 | 60·140 | 26·377 | 13 | dominated |
| HIVE – 1 dose | 3·3 | 3·9 | 6·9 | 60·140 | 26·378 | 13 | dominated |
| HR-HIVE – Extended | 3·3 | 3·6 | 6·7 | 60·142 | 26·378 | 13 | dominated |
| HIVE – 2 doses | 3·3 | 3·5 | 6·6 | 60·143 | 26·379 | 12 | dominated |
| ALL – 1 dose | 3·0 | 3·8 | 6·6 | 60·145 | 26·380 | 26 | dominated |
| ALL – 2 doses | 3·0 | 3·4 | 6·3 | 60·145 | 26·380 | 41 | dominated |
| HIVE – Extended | 3·3 | 2·7 | 5·8 | 60·149 | 26·381 | 12 | cost-saving*·† |
| ALL – 1 dose plus HIVE - Extended | 3·0 | 2·6 | 5·5 | 60·154 | 26·383 | 25 | 6264 |
| ALL - Extended | 3·0 | 2·2 | 5·1 | 60·160 | 26·384 | 90 | 57 976 |
| **South Africa [CET: ICER ≤ $1131/YLS (20% GDP per capita), ICER ≤ $2828/YLS (50% GDP per capita)]** | | | | | | | |
| ***Low postpartum maternal retention in care (50% of mothers with known HIV status in care at 6+ months)*** | | | | | | | |
| Standard-of-care | 2·3 | 2·8 | 5·0 | 68·827 | 28·467 | 125 | Reference |
| HR-HIVE – 1 dose | 2·1 | 2·7 | 4·6 | 68·883 | 28·486 | 128 | dominated |
| HR-HIVE – 2 doses | 2·1 | 2·6 | 4·5 | 68·893 | 28·490 | 127 | dominated |
| HR-HIVE – Extended | 2·1 | 2·5 | 4·4 | 68·908 | 28·494 | 126 | dominated |
| HIVE – 1 dose | 2·0 | 2·6 | 4·4 | 68·919 | 28·499 | 117 | dominated |
| ALL – 1 dose | 1·8 | 2·3 | 4·0 | 68·970 | 28·517 | 128 | dominated |
| HIVE – 2 doses | 2·0 | 2·2 | 4·1 | 68·973 | 28·517 | 114 | dominated |
| HIVE – Extended | 1·8 | 1·9 | 3·6 | 69·050 | 28·544 | 132 | dominated |
| ALL – 2 doses | 2·0 | 1·7 | 3·5 | 69·057 | 28·545 | 106 | cost-saving |
| ALL – 1 dose plus HIVE - Extended | 1·8 | 1·5 | 3·2 | 69·109 | 28·563 | 116 | 575 |
| ALL - Extended | 1·8 | 1·0 | 2·7 | 69·192 | 28·590 | 139 | 847*·† |

**Supplementary Table 22.** One-way sensitivity analysis: postpartum maternal retention in care (cont.)

| Country/strategy | Clinical outcomes: IU/IP cumulative HIV incidence (%) | Postnatal cumulative HIV incidence (%) | Total cumulative HIV incidence (%) | Lifetime efficacy and costs: Undiscounted life expectancy (yrs) | Discounted life expectancy (yrs) | Discounted costs ($) | ICER ($/YLS) |
|---|---|---|---|---|---|---|---|
| **South Africa [CET: ICER ≤ $1131/YLS (20% GDP per capita), ICER ≤ $2828/YLS (50% GDP per capita)]** | | | | | | | |
| ***High postpartum maternal retention in care (100% of mothers with known HIV status in care at 6+ months)*** | | | | | | | |
| Standard-of-care | 2·3 | 2·0 | 4·2 | 68·956 | 28·510 | 108 | Reference |
| HR-HIVE – 1 dose | 2·1 | 1·9 | 3·8 | 69·011 | 28·529 | 111 | dominated |
| HR-HIVE – 2 doses | 2·1 | 1·9 | 3·8 | 69·017 | 28·531 | 111 | dominated |
| HR-HIVE – Extended | 2·1 | 1·8 | 3·7 | 69·024 | 28·533 | 110 | dominated |
| HIVE – 1 dose | 2·0 | 1·8 | 3·6 | 69·043 | 28·540 | 101 | dominated |
| ALL – 1 dose | 2·0 | 1·6 | 3·5 | 69·070 | 28·549 | 100 | dominated |
| HIVE – 2 doses | 1·8 | 1·6 | 3·3 | 69·094 | 28·558 | 111 | dominated |
| ALL – 2 doses | 2·0 | 1·4 | 3·2 | 69·108 | 28·561 | 99 | cost-saving |
| HIVE – Extended | 1·8 | 1·3 | 3·0 | 69·147 | 28·576 | 118 | dominated |
| ALL – 1 dose plus HIVE - Extended | 1·8 | 1·2 | 2·9 | 69·160 | 28·580 | 109 | 569 |
| ALL - Extended | 1·8 | 0·7 | 2·4 | 69·240 | 28·606 | 133 | 921*·† |
| **Zimbabwe [CET: ICER ≤ $243/YLS (20% GDP per capita), ICER ≤ $607/YLS (50% GDP per capita)]** | | | | | | | |
| ***Low postpartum maternal retention in care (50% of mothers with known HIV status in care at 6+ months)*** | | | | | | | |
| Standard-of-care | 4·3 | 7·5 | 11·3 | 68·227 | 28·003 | 86 | Reference |
| HR-HIVE – 1 dose | 3·7 | 7·2 | 10·4 | 68·271 | 28·019 | 85 | dominated |
| HR-HIVE – 2 doses | 3·7 | 7·0 | 10·2 | 68·282 | 28·022 | 84 | dominated |
| HIVE – 1 dose | 3·7 | 7·0 | 10·2 | 68·288 | 28·025 | 81 | dominated |
| HR-HIVE – Extended | 3·7 | 6·5 | 9·8 | 68·305 | 28·030 | 81 | dominated |
| ALL – 1 dose | 3·4 | 6·7 | 9·7 | 68·308 | 28·032 | 93 | dominated |
| HIVE – 2 doses | 3·7 | 6·4 | 9·6 | 68·322 | 28·037 | 78 | dominated |
| ALL – 2 doses | 3·4 | 5·9 | 9·0 | 68·358 | 28·048 | 106 | dominated |
| HIVE – Extended | 3·7 | 4·5 | 7·8 | 68·415 | 28·066 | 68 | cost-saving*·† |
| ALL – 1 dose plus HIVE - Extended | 3·4 | 4·2 | 7·3 | 68·435 | 28·074 | 81 | 1738 |
| ALL - Extended | 3·4 | 3·4 | 6·5 | 68·474 | 28·087 | 140 | 4438 |
| ***High postpartum maternal retention in care (100% of mothers with known HIV status in care at 6+ months)*** | | | | | | | |
| Standard-of-care | 4·3 | 4·9 | 8·8 | 68·356 | 28·045 | 66 | Reference |
| HR-HIVE – 1 dose | 3·7 | 4·6 | 7·9 | 68·401 | 28·062 | 65 | dominated |
| HR-HIVE – 2 doses | 3·7 | 4·5 | 7·8 | 68·407 | 28·064 | 65 | dominated |
| HIVE – 1 dose | 3·7 | 4·5 | 7·7 | 68·415 | 28·066 | 61 | dominated |
| HR-HIVE – Extended | 3·7 | 4·3 | 7·6 | 68·418 | 28·067 | 64 | dominated |
| HIVE – 2 doses | 3·7 | 4·1 | 7·4 | 68·433 | 28·073 | 61 | dominated |
| ALL – 1 dose | 3·4 | 4·2 | 7·3 | 68·435 | 28·074 | 74 | dominated |
| ALL – 2 doses | 3·4 | 3·7 | 6·8 | 68·469 | 28·084 | 88 | dominated |
| HIVE – Extended | 3·7 | 3·3 | 6·6 | 68·475 | 28·086 | 59 | cost-saving*·† |
| ALL – 1 dose plus HIVE - Extended | 3·4 | 3·0 | 6·1 | 68·495 | 28·093 | 71 | 1724 |
| ALL - Extended | 3·4 | 2·2 | 5·4 | 68·534 | 28·107 | 131 | 4539 |

*bNAb: broadly neutralizing antibody; IU/IP: intrauterine/intrapartum; yr: year; ICER: incremental cost-effectiveness ratio; YLS: years of life saved; CET: cost-effectiveness threshold; HR-HIVE: high-risk HIV-exposed infants; HIVE: all HIV-exposed infants; ALL: all live infants at birth.*

Pediatric HIV incidence is rounded to the nearest tenth of a percent. IU/IP HIV incidence is calculated based on the number of infants exposed to HIV at birth. Postnatal and total HIV incidence is calculated based on the number of infants ever exposed to HIV through 36 months of life. Undiscounted and discounted life expectancies are rounded to the nearest ten thousandth. Costs are rounded to the nearest dollar and are presented in 2020 USD. Discounted values are discounted at 3% per year. ICERs are rounded to the nearest dollar and are calculated using unrounded discounted life expectancy and discounted costs. The cost-effective bNAb strategy was the strategy that offered the greatest increase in overall population life expectancy while still having an ICER less than the cost-effectiveness threshold when compared to the next best performing, non-dominated strategy. *Indicates the cost-effective strategy using a cost-effectiveness threshold of 20% GDP per capita. †Indicates the cost-effective strategy using a cost-effectiveness threshold of 50% GDP per capita.
